# Sexual health matters. Sexual Problems of Patients with Subacute Spinal Cord Injury and Their Relationships to dimensions of Sexual Quality of Life

**DOI:** 10.1101/2025.08.19.25334038

**Authors:** Alicja Widuch-Spodyniuk, Beata Tarnacka, Bogumił Korczyński, Aleksandra Borkowska

## Abstract

**Background:** Spinal cord injuries (SCI) are associated with a range of sexual dysfunctions that impede both sexual fulfillment and reproductive capability. This study analyzes the prevalence and severity of sexual problems following SCI-classified as primary, secondary, and tertiary dysfunctions-and examines their relationships with three key components of sexual quality of life: sexual activity, sexual interest, and sexual satisfaction in both male and female patients in the subacute phase of SCI (i.e., within two years post-injury).

**Methods:** A total of 110 patients with SCI were enrolled in the study. Analyses of primary sexual dysfunctions included 91 participants, while 81 were included in subsequent analyses. Data collection tools comprised: the *International Spinal Cord Injury Male Sexual Function Basic Data Set* (Version 2.0), the *International Spinal Cord Injury Female Sexual Function Basic Data Set*, the Ashworth Scale, SCIM III, the Numeric Rating Scale for Pain (NRSP), the Depression Assessment Questionnaire (KPD), the *State-Trait Anxiety Inventory* (STAI X-1), and numerical and verbal scales measuring subjective physical sexual attractiveness and the three domains of sexual quality of life. Additional data were gathered on bladder sphincter function, cardiac status, autonomic dysreflexia, and sleep apnea.

**Results:** Among participants, 87.65% of men and 83.35% of women reported at least one primary sexual dysfunction. In men with AIS A classification, a negative correlation was observed between ejaculatory dysfunction and sexual activity (rho = -0.50, p = 0.028). In men with AIS B, C, or D, orgasmic dysfunction negatively correlated with both sexual satisfaction (rho = -0.41, p = 0.024) and sexual activity (rho = -0.45, p = 0.012), while reflex erection was positively correlated with sexual interest (rho = 0.45, p = 0.007). Functional independence post-SCI (SCIM III) among men with AIS B, C, or D was associated with higher levels of sexual activity (rho = 0.31, p = 0.047). In AIS A men, a significant positive correlation was found between walking independence (WISCI II) and sexual interest (rho = -0.43, p = 0.031). Among AIS A women, spasticity severity negatively correlated with both sexual satisfaction (rho = -0.74, p = 0.036) and sexual activity (rho = -0.80, p = 0.017).

At the tertiary level, anxiety in women negatively correlated with sexual activity (rho = -0.79, p = 0.021 and rho = -0.84, p = 0.036), and with sexual satisfaction in AIS A women (p = 0.081). In AIS B, C, or D men, depression was associated with reduced sexual satisfaction (rho = -0.37, p = 0.017), and both anxiety and depression correlated negatively with sexual interest (rho = -0.38, p = 0.012 and rho = -0.47, p = 0.002, respectively). A strong positive correlation was identified between subjective sexual attractiveness and all three domains of sexual quality of life: interest (rho = 0.75), activity (rho = 0.82), and satisfaction (rho = 0.48) (p ≤ 0.001 for all domains).

Moreover, increased age and longer time since injury among women were associated with greater reflex genital arousal dysfunction. In men, advancing age correlated with increased reflex erectile dysfunction. Interestingly, in AIS B, C, or D men, a longer duration post-injury was linked to fewer psychogenic erectile dysfunctions.

**Conclusion:** The majority of participants exhibited at least one primary sexual dysfunction. Across all three domains of sexual quality of life, individuals with SCI scored significantly lower than the general population. Statistically significant associations were found between these domains and primary sexual dysfunctions, as well as secondary and tertiary SCI-related symptoms affecting sexual health.

## 1. Introduction

Sexual functioning affects the overall quality of life in patients with spinal cord injury (SCI), and its recovery is considered one of the key priorities in rehabilitation [1–5].

Sexual problems manifest in various forms and are influenced by somatic, psychological, relational, cultural, and social factors [6–8].

F.W. Foley [9] identified three levels of factors influencing sexual problems (SP) in individuals with multiple sclerosis, which may be applicable to the sexual lives of individuals with SCI: primary (directly related to changes in the central nervous system), secondary (complications resulting from SCI that are not directly related to neurological damage), and tertiary (psychosocial factors following SCI that may negatively affect sexuality) [9–10].

### 1.1. Primary and Secondary Levels of Sexual Dysfunction Associated with Spinal Cord Injury

#### 1.1.1 The primary level

The primary level of sexual dysfunction following SCI is directly related to spinal cord trauma (i.e., alterations or loss of sensation and function in the genital area, anorgasmia, and biologically induced decreases in libido). One of the most vulnerable domains after SCI is the ability to experience sexual arousal. The activation of reflex sexual arousal in both sexes is primarily mediated by the parasympathetic nervous system located in the sacral spinal cord segments S2–S4 [11–13]. In contrast, psychogenic arousal involves the sympathetic nervous system at the T11– L2 spinal segments.

Erectile dysfunction of varying severity, as well as problems with ejaculation, affect the majority of men following SCI. Within two years post-injury, the ability to achieve an erection returns in approximately 86% of men; however, it is often insufficient for penetrative intercourse [14].

Ejaculation is a coordinated neuromuscular event. The spinal cord segments responsible for ejaculation are located at the L3–L4 levels. The process of ejaculation primarily involves the sympathetic nerve structures situated in the T10–L2 segments, in conjunction with the coordinated activity of the parasympathetic nervous system located at S2–S4 and the activation of the somatic nervous system via the pudendal nerve at S2–S5 [11,13,15]. Ejaculatory dysfunction affects between 89.5% and nearly 100% of men [16–19], significantly impacting their fertility. A normal ejaculatory response is most likely to occur in individuals with incomplete injury to the conus medullaris and the cauda equina structures [20].

The majority of men with spinal cord injury either lack ejaculation altogether or experience altered ejaculatory patterns, with semen often exhibiting reduced sperm motility and viability.

Difficulty achieving orgasm is among the most frequently reported dysfunctions following SCI [19, 21–22]. There is no single neurophysiological definition of orgasm; therefore, most research on this phenomenon relies on self-reported data, physiological measures (such as heart rate and blood pressure), or documentation of phenomenological experiences [23].

Analyses concerning orgasm retention in women and men with SCI remain inconclusive: it is estimated that between 26.6% and 50% of women [24–25], and approximately 40% to 45% of men [26–27], retain the ability to experience orgasm. Studies involving both sexes indicate that 50% of individuals with SCI remain sexually active [27]. Differences in the ability to achieve full sexual satisfaction are primarily related to the level and extent of spinal cord injury. Women and men with damage to the lower motor neuron fibers located in the spinal segments S2 to S5 experience significantly greater difficulty achieving orgasm compared to patients with injuries at other levels and degrees of severity [13,28]. In cases of complete injury to this area, only 17% of women were able to reach orgasm [24], and the time required to achieve orgasm was prolonged compared to control groups-both during intercourse and masturbation [24]. Although orgasm is primarily a genital experience, it can also be triggered through stimulation of non-genital areas (e.g., nipples, ears, neck, anus) as well as through erotic fantasies [26].

Spinal cord injury may lead to menstrual cycle disturbances. However, many women with tetraplegia or paraplegia retain both menstruation and ovulation; approximately 50% resume menstruation within 3 to 8 months post-injury, and cycle regularity is typically maintained [29–31]. Spinal cord injury does not affect female fertility and is not a contraindication to becoming pregnant.

#### 1.1.2 Secondary symptoms

Secondary symptoms following SCI emerge immediately after the injury, and their consequences are typically permanent, depending on the timing, severity, and level of the injury [32].

Spinal cord injury disrupts or severs the control and bidirectional communication between centrally located neural centers and peripheral nerves innervating internal organs, leading to dysfunction of the autonomic nervous system (ANS) below the level of injury. SCI has serious consequences, including dysregulation of the cardiovascular, respiratory, endocrine, excretory, immune, and metabolic systems, as well as spasticity, neuropathic pain, and dysesthesia [32–34,16], and may even impair cognitive functioning [35]. Moreover, pharmacological treatments introduced to manage these complications can significantly affect sexual activity [36–37].

Cardiovascular conditions may contribute to sexual dysfunctions [38]. Difficulties in blood pressure regulation can vary depending on the type of injury-whether it involves upper or lower motor neuron damage [39]. Research by Foa et al. demonstrated that sexual dysfunctions following SCI affected 42.1% of women with hypertension, compared to only 19.4% in women with normal blood pressure [40]. Hypertension is also associated with erectile dysfunction in men [40–42,38]. The most severe form of blood pressure dysregulation is autonomic dysreflexia (AD). This condition, also referred to as hyperreflexia, occurs in patients with high-level spinal cord injuries (above T6) and is characterized by a sudden increase in blood pressure of 20–40 mmHg above baseline-where resting blood pressure in individuals with tetraplegia averages around 90 mmHg. Symptoms of AD include headache, myocardial infarction, and stroke. Autonomic dysreflexia is triggered by stimuli below the level of injury, most commonly due to bladder wall distension, bowel impaction, labor, skin injuries, bone fractures, and also during sexual activity - particularly ejaculation and genital stimulation [43–44]. In a study by K.D. Anderson et al., AD was reported in 39% of women and 29% of men; 28% of female and 16% of male participants indicated that AD had an impact on their sexual functioning [45].

Other biological factors contributing to the deterioration of sexual life include bladder dysfunction and loss of sphincter control. Bladder dysfunction, bowel impairment, and sexual dysfunction share common neurological pathways [1]. Additional challenges include an increased risk of bacterial infections associated with catheterization, frequent urinary tract infections, and the possibility of involuntary bowel movements during sexual activity. Discomfort caused by urinary or fecal incontinence significantly increases the risk of low sexual satisfaction among patients [46]. Moreover, the presence of a catheter-particularly an indwelling catheter used in cases of severe neurogenic bladder dysfunction-acts as a physical barrier to intercourse and causes considerable discomfort due to odor, visual appearance, and restricted movement [46–48].

Metabolic disorders - including diabetes, dyslipidemia, and osteoporosis-also contribute to sexual problems (SP). Diabetes is one of the most common comorbidities among individuals with SCI and has a negative impact on sexual functioning [49]. SCI may impair the activation of the PI3K signaling pathway in the hypothalamus, which can contribute to peripheral inflammation and the development of diabetes in patients [50–51].

Endocrine disorders indirectly associated with SCI contribute to sexual dysfunction. Correlational studies have shown that testosterone levels in individuals with SCI, both in the acute and chronic phases, are lower compared to age-matched controls. Low testosterone levels have been observed in 25% of men with SCI aged 18 to 45, compared to 6.7% in the control group [52]. Insufficient testosterone negatively affects erectile function and is correlated with reduced libido [53,48]. Muscle spasticity is one of the most common complications following SCI, particularly in patients with subacute injury and incomplete spinal cord damage, as well as in the chronic stage regardless of injury severity [54]. Muscle spasticity limits patients’ daily functioning and adversely affects sexuality by impairing positioning, mobility, muscle activation, and strength [55–56].

Approximately 80% of patients with SCI experience pain of varying intensity and type, with a significant portion-around 40%-classified as neuropathic pain [57–58].

The complexity of pain-related issues in patients may be associated with treatment resistance and a lack of satisfactory outcomes from implemented therapies [59].

Many medications used by individuals with SCI have an indirect impact on sexual dysfunction. Antihypertensive drugs such as thiazide diuretics, beta-blockers, and centrally acting agents inhibit the sympathetic nervous system [60]. Antidepressants from the selective serotonin reuptake inhibitor (SSRI) class may significantly affect libido and contribute to other forms of sexual dysfunction [61–62]. Antispasmodic and muscle relaxant agents can affect erection quality, while opioid analgesics may disrupt testosterone levels [63–64, 27].

### 1.2 Tertiary (psychosocial) level of sexual dysfunction following spinal cord injury

Tertiary symptoms of sexual dysfunction include mood disorders (such as depression and dysthymia), emotional disturbances, reduced self-efficacy, and diminished self-esteem related to changes in body image or the perceived ability to fulfill social roles. These factors may influence one’s perception of sexual attractiveness and alter interpersonal relationships.

Emotional and psychological problems have a significant impact on the frequency and quality of sexual activity. Low mood, depression, and anxiety disorders are among the most common secondary symptoms following spinal cord injury. The prevalence of anxiety and depression in women with SCI is higher compared to able-bodied women, reaching 51% and 53%, respectively, versus 26% and 29% in the control group [65].

In addition to the strong association between depression and reduced sexual drive identified in clinical studies, epidemiological research confirms the negative impact of depression on orgasmic experience-particularly among patients treated with SSRIs and SNRIs [66–67, 1]. The precise mechanism underlying sexual dysfunction in the context of low mood and depression is neither clear-cut nor unidirectional. Sadness, anhedonia, and psychomotor retardation may lead to difficulties with sexual arousal and diminish the ability to derive pleasure from intercourse or autoerotic activity. Some analyses also suggest a reverse direction of causality-namely, that sexual and relational difficulties may trigger depressive symptoms or contribute to a low mood. Individuals suffering from depression, dysthymia, or chronic low mood may have a parallel predisposition to sexual dysfunction [68]. Anxiety disorders likewise affect intimate life and may be associated with fears of involuntary bowel movements, inability to satisfy a partner, or negative self-perception in terms of sexual attractiveness [69]. People with anxiety often exhibit heightened sensitivity to symptoms associated with the emotional state itself-anxiety is triggered by the body’s own anxiety response. Physical sensations caused by sympathetic arousal-such as shortness of breath, elevated body temperature, muscle tension, and palpitations-are frequently misinterpreted as signs of anxiety, which may interfere with concentration and diminish the pleasure derived from physical sexual arousal. This results in the association of sexual arousal with anxiety symptoms, leading to the transference of anxiety onto the sexual act itself [70].

Individuals experiencing chronic anxiety may have difficulty achieving physiological arousal, including vaginal lubrication; during intercourse, they may experience significant discomfort or pain. These painful stimuli can become associated with sexual activity, potentially leading to avoidance of sex in the long term [71].

An important factor is the reduced sense of sexual attractiveness, defined as the subjective perception of diminished ability to elicit desire and interest from a partner, along with the anticipation of decreased sexual performance [45,72–73]. Among women with SCI, 74.7% reported that their self-image as a sexual being changed significantly after the injury [45]. SCI is often associated with muscle mass reduction, changes in body composition, extensive and aesthetically displeasing scarring, and skin conditions-all of which significantly affect self-esteem and perceived attractiveness in both men and women. Sexual attractiveness is also influenced by concerns related to involuntary bowel movements and unpleasant body odor. Changes in body image and dissatisfaction with one’s appearance are correlated with increased levels of depression and decreased sexual activity, including avoidance of sexual contact altogether [1,47]. Lowered self-worth may result from altered body image, viewing oneself as a less attractive sexual partner, or from previously internalized stereotypes regarding disability and gender roles. Physical disability and the resulting loss of independence negatively impact an individual’s perceived ability to fulfill culturally defined gender roles [74]. Tertiary aspects of spinal cord injury are broad and may serve as significant predictors of overall sexual quality of life in individuals with SCI. Some studies suggest that sexual health is more strongly associated with mental health than with physical functioning, stress levels, or age [68].

Stereotypes, ableism, and prejudice are frequently directed toward individuals with SCI, leading to social, educational, and occupational stigmatization, as well as self-stigmatization through the internalization of messages and meanings related to disability [7].

Cultural assumptions, biases, and stereotypes regarding gender roles and the sexuality of individuals with disabilities contribute to the loss of gender identity [75]. Social stereotypes portraying people with disabilities as asexual negatively affect women’s self-esteem in the context of their femininity. For men, the stereotype of constant sexual readiness and vitality exerts a detrimental influence on their sexual self-concept. Men who internalize this stereotype are more likely to experience frustration due to the gap between cultural expectations and their current capabilities, leading to reduced psychological well-being and diminished adaptability to intimacy-related changes following SCI [76]. Stereotypes surrounding male independence and physical strength are also of considerable importance [77–78].

Romantic relationships and their quality play a vital role in maintaining psychophysical well-being. Some studies indicate that women tend to prioritize emotional fulfillment and intimacy over physical sexual acts [79]. Having a steady partner positively influences both sexual activity and satisfaction [78,80–81]. In a study by K.D. Anderson et al. (2007), the primary motivation for engaging in sexual activity was the need for intimacy, followed by the desire to maintain a relationship [45]. For individuals of both sexes, the perception of a partner’s satisfaction was a significant factor in their own sexual fulfillment. The quality of the relationship is crucial to overall well-being-individuals in satisfying partnerships report feeling safe, accepted, and free in their sexual expression. Those in high-quality emotional relationships, characterized by deep connection, mutual acceptance, and a sense of security, tend to achieve better sexual adjustment. Such relationships foster creativity, openness to sexual experimentation, and a shared commitment to achieving mutual satisfaction while reducing anxiety surrounding sexual activity [73,78,82–83].

SCI often necessitates the modification of roles within the family or romantic relationship, as the roles of sexual partners may be overshadowed by caregiving responsibilities related to hygiene and self-care. Relationships between women and men following SCI require reconstruction and redefinition of roles, duties, and the ways in which both individual and partner needs are addressed [76,84]. It is essential to distinguish and balance the roles of man, woman, sexual partner, and caregiver or care recipient [69,85–86].

Due to the complexity of factors affecting sexuality in individuals with SCI, it is considered best practice to provide multidisciplinary, individualized care. Such care should take into account the patient’s age, time since injury, severity, extent, and level of the lesion, comorbid conditions, psychological state, relationship dynamics and support network, as well as current medications [7,15,86–87].

### 1.3. Factors related to overall quality of sexual Life

Three components of overall sexual quality of life have been identified: sexual interest, sexual activity, and sexual satisfaction, all of which are influenced by somatic and psychosocial factors (Fig. 1) [88–89].

**Figure 1.**
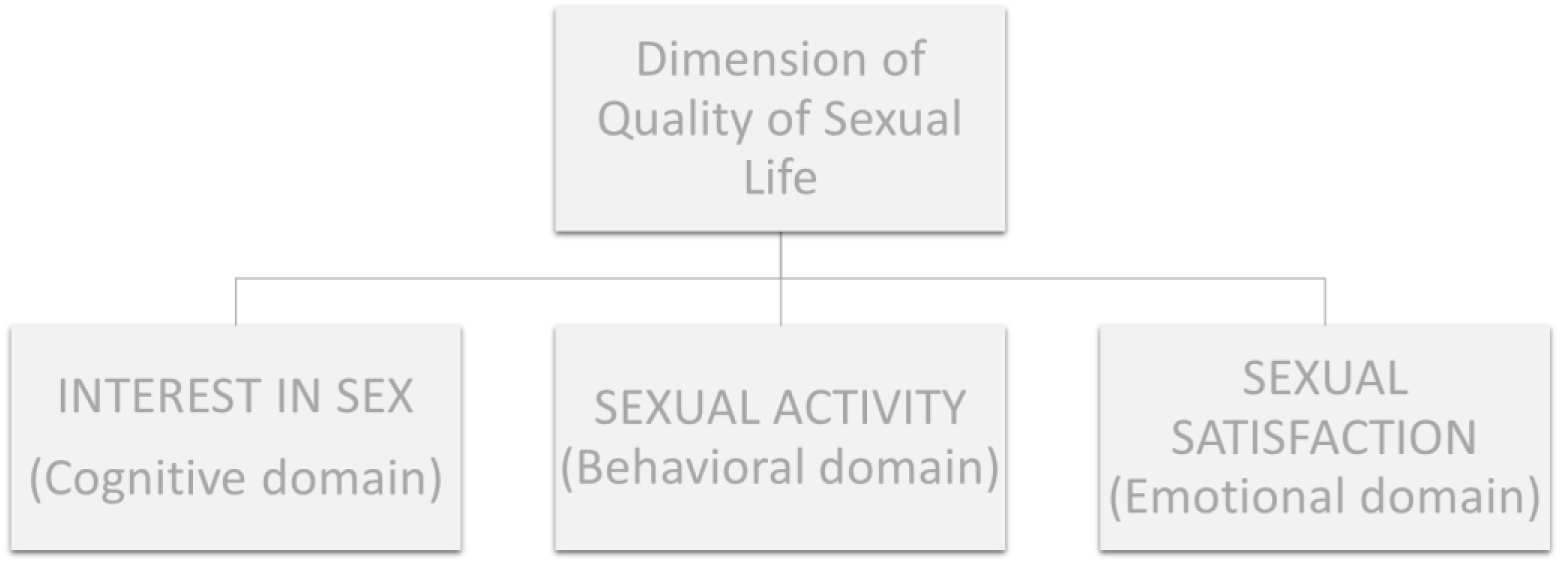
Domains of overall sexual quality of life after SCI – own elaboration.

The domains of overall sexual quality of life correlate with somatic health and psychosocial factors; however, the nature of these correlations is not always straightforward [88–89]. In a study by Dosch A. et al., a positive relationship was observed between sexual activity, sexual drive, and satisfaction [89]. The connection between activity and interest or desire can be complex-individuals may remain sexually active without experiencing desire, and conversely, they may feel sexual longing without fulfilling it due to factors related to partnership, physical health, or psychosocial circumstances [89].

Sexual interest is understood as a spontaneous cognitive phenomenon linked to an individual’s reaction to a specific situation, while desire is considered a more personalized experience encompassing both cognitive and physical domains [90]. The terms interest, desire, and arousal are often defined similarly; however, arousal is typically characterized as a physical phenomenon, non-specific to the individual, and regarded as a response to physical stimulation [90]. Sexual interest can be defined as a cognitive phenomenon involving thoughts and fantasies about engaging in sexual or erotic activities. It may also be associated with the emotional-motivational component of the sexual response, which corresponds to the initial stage of sexual response in Bancroft’s model [91].

Sexual activity encompasses all individual behaviors of a sexual or erotic nature-whether performed alone or with others, involving genital or non-genital stimulation-and includes behavioral aspects that contribute to sexual quality of life [92,88,73]. Sexual activity significantly decreases in both men and women following SCI [93]. Ferreiro-Velasco et al. observed a notable decline in the frequency of sexual intercourse after spinal cord injury: the average decreased from 9.9 times per month before injury to 4.2 times per month afterward [72]. This reduction was not related to the level of injury, neurological status as measured by the ASIA scale, time since injury, degree of independence, or control over sphincters. Research indicates a negative correlation between the severity of neurological impairment and sexual activity in men [94]. Predictors of lower sexual activity include bladder and bowel sphincter dysfunction, as well as the risk of autonomic dysreflexia [93,95].

Longitudinal studies have shown that six months after injury and discharge from institutional treatment, sexual activity tends to increase, although it does not return to pre-injury levels [73,29]. Despite reduced sexual activity, both men and women with SCI report that sexuality remains an important aspect of their lives [96]. In a study by Reitz A., 53% of participants described their current level of sexual desire as very high, high, or higher compared to before the injury, while 44% reported a similar level [97].

Sexual satisfaction is defined as the subjective level of contentment with one’s sexual activity, based on a comparison between one’s current experience and personal norms and standards [98]. As an emotional component of overall sexual quality of life, sexual satisfaction is influenced by both somatic and psychosocial factors. Sexual satisfaction is strongly correlated with overall life satisfaction and, for many patients, represents a high priority in the rehabilitation process following spinal cord injury [48]. Reduced sexual satisfaction is one of the most vulnerable factors following SCI [78].

A multivariate analysis by Sale et al. showed that 31% of women and men with SCI were satisfied with their sexual lives, and the level of injury did not correlate with sexual satisfaction. However, there appears to be a relationship between neurological status and sexual quality of life [83]. Time since injury may play an important role. Research by Biering-Sørensen found that 54% of individuals reported sexual satisfaction at least 10 years after spinal cord injury [99]. Preserved erectile function has been identified as a predictor of higher sexual satisfaction in men [80]. Interestingly, men with high-level spinal cord injuries report greater satisfaction with their sexual lives. Individuals with upper motor neuron lesions more frequently retain erectile function compared to those with lower motor neuron injuries [46]. Factors negatively correlated with sexual satisfaction include gender, age, occupation, type of injury, relationship status, bladder function, level of independence, and overall life satisfaction [100–101,83].

Younger age at the time of injury is a predictor of higher sexual life satisfaction; however, no correlation has been observed between satisfaction and the amount of time since the injury occurred [82]. Physiological factors tend to have a greater impact on sexual satisfaction in men, whereas women are more sensitive to psychological factors. Women’s preferences and expectations are more often centered on aspects of touch and intimate connection-such as kissing, cuddling, and caressing-rather than on intercourse itself [83]. [83].

## 2. Case Presentation

This study is part of a prospective clinical trial approved by an ethics committee and conducted in accordance with the Declaration of Helsinki. The project received approval from the Ethics Board of the Regional Medical Chamber in Szczecin, Poland (Approval No. OIL-Sz/MF/KB/452/05/07/2018; No. OIL-SZ/MF/KB/450/UKP/10/2018).

Patients were recruited through self-selection from the general population of people with SCI in Poland. (Patient recruitment lasted from 19 April 2018 to 13 December 2021).

The objectives of the study and specific research questions are as follows:

1. Analysis of sexual problems in individuals following spinal cord injury.
  a. What are the main sexual problems at the primary, secondary, and tertiary levels in patients in the early stage post-SCI (i.e., type, number, and scope)?
  b. Do the extent and severity of injury (AIS A vs. AIS B, C, D), time since SCI, and age differentiate patients in terms of the type, number, and severity of primary, secondary, and tertiary sexual problems after SCI?
2. Analysis of the impact of sexual problems on the domains of sexual quality of life.
  a. What is the dynamic of the three domains of sexual quality of life in individuals with SCI?
    i. Is there a relationship between the three domains of sexual quality of life: sexual interest, sexual activity, and sexual satisfaction?
    ii. Does the extent and severity of spinal cord injury differentiate SCI patients in terms of the domains of sexual quality of life?
    iii. Are age and time since injury associated with the domains of sexual quality of life in patients with SCI?
  b. Is there a relationship between sexual problems at the primary, secondary, and tertiary levels and the domains of sexual quality of life in individuals with SCI?
  c. Do individuals with spinal cord injury differ from the general population in terms of the domains of sexual quality of life?

### 2.1. Study Inclusion Criteria

The study participants were self-recruited from the general population of individuals with SCI in Poland, primarily through the website of the Research Institute for Innovative Rehabilitation Methods for People with Spinal Cord Injury: https://uzdrowisko-kamienpomorski.pl/instytut-badawczy/. Prior to enrollment, all participants underwent medical and psychological evaluations conducted by a team of specialists, including a neurologist, a physical medicine and rehabilitation physician, physiotherapists, and a psychologist. The medical inclusion criteria were as follows: time since injury between 3 months and 2 years; general condition defined as conscious, cooperative with the physiotherapist, and able to maintain an upright position; complete or incomplete SCI (at the cervical, thoracic, or lumbar level) with spinal stabilization in the final stage of bone fusion; and absence of contraindications to rehabilitation (e.g., venous thrombosis, pulmonary embolism, orthostatic hypotension, epilepsy, infections).

The exclusion criteria included: high and complete tetraplegia, very low lumbar spinal cord injury, incomplete bone fusion following spinal surgery and stabilization, respiratory failure, cardiovascular insufficiency of class III or IV according to the New York Heart Association (NYHA) classification, past or present neurological disorders (e.g., traumatic spinal stroke, multiple sclerosis, cerebral palsy), menopause or postmenopausal status, and pre-existing sexual dysfunctions prior to spinal cord injury.

Additional psychological exclusion criteria were: reduced intellectual functioning that prevented the administration of a structured interview and questionnaire-based assessments, age under 18, and lack of prior sexual experience.

All participants signed an informed consent form for participation in the study.

### 2.2. Participants

A total of 158 individuals with SCI enrolled in the studies conducted as part of the seven-week rehabilitation program at the Research Institute for Innovative Rehabilitation Methods. All patients consented to participate and signed an informed consent form prior to the start of the study. Forty-eight patients did not meet the medical and psychological eligibility criteria.

The study included a group of 110 adult participants (N = 110), comprising 89 men and 21 women with neurological impairments following SCI.

A total of 91 individuals were included in the analyses related to primary sexual problems, while 81 participants were included in the remaining analyses due to missing data.

Patients who qualified for the study were randomly assigned by a blinded researcher to one of two study groups using coin-flip randomization. Participants in the S0 group (control group) underwent conventional rehabilitation along with 30-minute sessions using a dynamic parapodium trainer (DPT). The S1 group (experimental group), in addition to conventional therapy, received rehabilitation using Robotic-Assisted Gait Training (RAGT)-with either an exoskeleton or the Lokomat-also in 30-minute sessions. The rehabilitation program lasted for seven weeks, with a one-week break after the third week.

From the study participants, three subgroups were identified based on the following criterion:

– rehabilitation modality (S0 = 31; S1 = 79),
– extent and completeness of injury, assessed using the ASIA (American Spinal Injury Association) Impairment Scale: AIS A (n = 44; 35 men, 9 women) and AIS B, C, D (n = 66; 54 men, 12 women),
– level of injury: 24 patients (20 men and 4 women) had cervical spinal cord injuries (Cervical Vertebrae C1–C7); 57 patients (46 men and 11 women) had thoracic injuries (Thoracic Vertebrae Th1–Th12); and 29 patients (23 men and 6 women) had lumbar injuries (Lumbar Vertebrae L1–L5).

Using the Mann-Whitney U test (for time since injury and age) and the chi-square test of independence along with Fisher’s exact test, it was assessed whether the male and female groups differed in terms of their group characteristics. The results of these analyses revealed no significant differences between the groups in terms of sociodemographic variables or injury-related factors (Table 1).

**Table 1.** Characteristics of the study group.

| Group | Male ($N = 89$; 80.9%) | Female ($N = 21$; 19.1%) | $p$ |
| --- | --- | --- | --- |
| **Age** |  |  |  |
| Median (IQR) | 36 (24) | 41 (28) | 0.816 |
| **Education** |  |  |  |
| In the course of education | 4 (4.5%) | 2 (9.5%) | 0.056<sup>a</sup> |
| Elementary | 2 (2.2%) | 0 (0.0%) |  |
| Vocational | 12 (13.5%) | 1 (4.8%) |  |
| Secondary | 24 (27.0%) | 1 (4.8%) |  |
| Higher | 47 (52.8%) | 17 (81.0%) |  |
| **Accomodation** |  |  |  |
| Countryside | 24 (27.3%) | 6 (30.0%) | 0.192 |
| Small town | 21 (23.9%) | 6 (30.0%) |  |
| Medium-size town | 17 (19.3%) | 0 (0.0%) |  |
| Big city | 26 (29.5%) | 8 (40.0%) |  |
| **Marital status** |  |  |  |
| Single | 21 (23.6%) | 6 (28.6%) | 0.865 |
| Informal relationship | 16 (18.0%) | 4 (19.0%) |  |
| Formal relationship | 52 (58.4%) | 11 (52.4%) |  |
| **Sex orientation** |  |  |  |
| Heterosexual | 71 (97.3%) | 18 (100.0%) | 1.000<sup>a</sup> |
| Homosexual | 2 (2.7%) | 0 (0.0%) |  |
| Bisexual | 0 (0.0%) | 0 (0.0%) |  |
| **Time from accident (months)** |  |  |  |
| Median (IQR) | 12 (13) | 12 (11) | 0.715 |
| **Cause of injury** |  |  |  |
| Vehicle accident | 30 (34.1%) | 6 (28.6%) | 0.179<sup>a</sup> |
| Fall < 1 m | 6 (6.8%) | 1 (4.8%) |  |
| Fall > 1 m | 31 (35.2%) | 6 (28.6%) |  |
| Dive | 4 (4.5%) | 0 (0.0%) |  |
| Violence-related trauma | 0 (0.0%) | 1 (4.8%) |  |
| Body crushing | 5 (5.7%) | 0 (0.0%) |  |
| Others | 12 (13.6%) | 7 (33.3%) |  |
| **Level of neurological impairment** |  |  |  |
| Cervical | 20 (22.5%) | 4 (19.0%) | 0.932 |
| Thoracic | 46 (51.7%) | 11 (52.4%) |  |
| Lumbar | 23 (25.8%) | 6 (28.6%) |  |
| **ASIA** |  |  |  |
| AIS A | 35 (39.3%) | 9 (42.9%) | 0.766 |
| AIS B/C/D | 54 (60.7%) | 12 (57.1%) |  |
| **Rehabilitation Modality** |  |  |  |
| S0 (DPT) | 27(30,3%) | 4(19,0%) |  |
| S1 (RAGT) | 62(69,7%) | 17(81,0%) |  |

Note. <sup>a</sup> – exact Fisher's test; Rehabilitation modality: S0 – conventional rehabilitation using a dynamic parapodium (DPT), S1 – robotic rehabilitation.

### 2.2.Physiotherapy and psychological impact

Conventional rehabilitation was carried out using a dynamic parapodium (DPT). RAGT rehabilitation was conducted using either an exoskeleton (EKSO 1 model, manufactured by Ekso Bionics, 2014) or the Lokomat system (Locomat Pro, model LO218, manufactured by Hocoma AG, 2014). Each training session was conducted and supervised by a trained and qualified physiotherapist. All rehabilitation interventions were delivered in the form of 30-minute sessions.

Robotic-assisted gait training (RAGT) is a safe and well-tolerated rehabilitation modality that utilizes advanced devices such as Lokomats and exoskeletons. The effectiveness of RAGT in the rehabilitation of patients with spinal cord injury (SCI) remains inconclusive [102–103]. However, many studies indicate a beneficial impact of robotic rehabilitation on improving patient mobility, gait function, muscle strength, and reducing spasticity and pain [104–108]. Due to the potential for increasing exercise intensity when using robotic devices, it is assumed that they positively influence the cardiorespiratory fitness of patients with SCI [109]. The best outcomes are observed in individuals with incomplete spinal cord injury [102–103].

Moreover, studies suggest that training with the use of robotic rehabilitation devices has a positive impact on self-esteem, mood, and increases hope among individuals after SCI [110–111].

All study participants received psychological support in the form of 50-minute individual or couple sessions, as well as 90-minute group therapy sessions. Each patient attended a total of 7 individual and 6 group sessions. Individual therapy was tailored to each patient’s specific difficulties and current needs, based on their personal and environmental resources, as well as their current capabilities. Group therapy focused on psychoeducation, sharing experiences, peer support, and the development of social skills. Two group sessions were devoted to issues of sexuality (sexual dysfunction after SCI, its etiology, prevalence, and treatment methods, improving partner relationships, self-worth, and sexual attractiveness in relational and sexual contexts). The sessions were conducted using evidence-based methods and techniques, such as Cognitive-Behavioral Therapy (CBT), with elements of Rational Behavior Therapy, Motivational Interviewing, and Mindfulness-Based Cognitive Therapy [112–114].

## 3. Methods

### 3.1. Study Protocol

The study employed research tools with proven validity and reliability. The research procedure followed the sequence outlined below:

1. A medical physical and neurological examination, including a medical interview and questionnaire assessments using the following instruments: the Spinal Cord Independence Measure, Version III (SCIM-III), the Walking Index for Spinal Cord Injury, Version II (WISCI-II), the American Spinal Cord Injury Association (ASIA) Impairment Scale (AIS), and the Ashworth Scale;
2. Psychological assessment: a structured interview covering demographic data, the Numeric Rating Scale for Pain (NRS Pain), the Sexual Activity Rating Scale, the Scale of Overall Satisfaction with Sex Life, and the Sex Interest Rating Scale (Annex 1), as well as standardized questionnaires including: the International Spinal Cord Injury Male Sexual Function Basic Data Set (Version 2.0), the International Spinal Cord Injury Female Sexual Function Basic Data Set, the Depression Assessment Questionnaire (KPD), and the State-Trait Anxiety Inventory (STAI, Form X-1).

To ensure consistency of the research procedure, the psychologist read the questionnaire items aloud to the study participants and recorded their responses. This approach was adopted due to motor difficulties experienced by some respondents, which made it challenging for them to mark their answers or turn the pages of the questionnaires.

It should be emphasized that the present study was not conducted in accordance with the initial research plan, which included a follow-up measurement of the studied variables after one year. During the subsequent assessment-after a one-year latency period-the majority of patients from the control group (S0) reported that, in the meantime, they had undergone robotic rehabilitation at private medical facilities. This prevented the analysis of potential changes in factors related to sexual problems and components of overall sexual quality of life.

### 3.2. Outcome measures

#### 3.2.1. Primary outcome measures

To assess the primary symptoms of sexual dysfunction following SCI, the International Spinal Cord Injury Male Sexual Function Basic Data Set (Version 2.0) and the International Spinal Cord Injury Female Sexual Function Basic Data Set were used. These tools were developed by the International Spinal Cord Society (ISCoS), the American Spinal Injury Association (ASIA), and a representative of the ASIA International Standards and Data Sets Executive Committee [115].

The self-report-based questionnaires on male and female sexual function enable the assessment of: the patient’s interest in discussing sexual functioning after SCI (SEXDISCU), sexual orientation (SEXORIEN), the presence of sexual dysfunction prior to the injury or unrelated sexual issues (SEXPRBUN), sexual dysfunction related to spinal cord injury (SEXDYSCL), as well as the extent of difficulties with psychogenic and reflex genital arousal, ejaculation, ability to reach orgasm, and menstruation [115–116]. For the purpose of this study, this tool was validated (see Appendix 1).

To measure secondary sexual problems after spinal cord injury (SCI), the following tools were used:

Numerical Rating Scale for Pain (NRS Pain). This tool includes an 11-point pain intensity scale ranging from 0 to 10, where 0 means “no pain” and 10 means “pain as bad as you can imagine” [117]. Following Forchheimer et al., we adopted the following cut-off points: 1–3 – no or mild pain; 4–6 – moderate pain; 7–10 – severe pain [118].

A modified version of the Ashworth Scale, commonly used to assess increased muscle tone (spasticity), was applied. This is a five-point scale, where: 0 indicates no increase in muscle tone, and 4 indicates rigid immobility of the assessed limb in flexion or extension [119].

The Walking Index for Spinal Cord Injury II (WISCI-II) – this tool enables the assessment of improvements in walking function in patients after SCI. It is the most sensitive measure for detecting changes in walking ability compared to other leading assessment scales. The index provides a graded evaluation of walking function: Level 0 indicates an inability to stand or participate in walking, while Level 21 indicates the ability to walk without assistance, orthoses, or support from others [120–121].

The Spinal Cord Independence Measure III (SCIM III) is used to assess functional independence after SCI. It is a reliable and valid scale for evaluating the daily functioning skills of patients with spinal cord injury. Version III consists of 19 items grouped into three areas: Self-care, Respiration and sphincter management, Mobility. The total score ranges from 0 (indicating complete dependence) to 100 (indicating complete independence) [122–124].

To assess the severity of tertiary (psychosocial) symptoms related to depression, the Depression Assessment Questionnaire (KPD) was used. This questionnaire measures the intensity of depression and its individual symptoms. It consists of five subscales: Cognitive deficits and loss of energy (DPUE), Thoughts of death, pessimism, and alienation (MSPA), Feelings of guilt and anxiety tension (PWNL), Psychosomatic symptoms and loss of interest (OPSZ), and Self-regulation (SR). The total score (WO) is calculated as the sum of the first four subscales [125].

To measure state anxiety, the X-1 subscale of the State-Trait Anxiety Inventory (STAI) was used. State anxiety is understood as tension related to the current situation [126]. The study also employed the Subjective Sexual Attractiveness Scale (ASS). Patients performed a self-assessment using a five-point scale, where: 1 indicated the lowest perceived level of attractiveness, and 5 indicated a high level of perceived sexual attractiveness (Appendix 2).

To measure the three domains of sexual quality of life, numerical subjective scales were used (developed by the authors): Sexual Interest Rating Scale (SIRS), Sexual Activity Rating Scale (SxARS), and Scale of Overall Satisfaction with Sex (SSWS). A five-point scale was applied to assess these variables: Sexual activity was rated as: 1 – very low, 2 – rather low, 3 – moderate, 4 – rather high, 5 – very high; Satisfaction with sexual life was rated as: 1 – completely unsatisfactory, 2 – rather unsatisfactory, 3 – moderately satisfactory, 4 – rather satisfactory, 5 – completely satisfactory; Sexual interest was rated as: 1 – very low, 2 – rather low, 3 – moderate, 4 – rather high, 5 – very high. (See Appendix 3.).

#### 3.2.2. Secondary outcome measures

To assess neurological and functional status after SCI, the American Spinal Injury Association Impairment Scale (AIS) was used. This scale evaluates motor, reflex, and sensory functions and allows for the classification of spinal cord injuries as A, B, C, D, or E: AIS A indicates a complete SCI, AIS B, C, and D represent varying degrees of incomplete SCI, AIS E indicates no neurological impairment and preserved motor function [127–129]. In this study, all participants exhibited neurological symptoms. For the purposes of AIS classification, participants were divided into two groups: AIS A and a combined group including those with spinal cord injuries classified as AIS B, C, or D.

### 3.3. Statistical analyses

In the present analysis, continuous data were presented as mean values (M) and standard deviations (SD), while categorical data were reported as percentages. Initially, data distributions were examined, and basic descriptive statistics for quantitative variables were calculated. The Shapiro–Wilk test was used to assess the normality of distributions, which showed that most of the variables did not follow a normal distribution (p < 0.05). Additionally, skewness and kurtosis values were considered, many of which exceeded the conventional threshold of |2| [130] - see Table 1. Due to the violation of the normality assumption and the presence of numerous outliers (values exceeding the third standard deviation), further analyses were conducted using non-parametric tests.

In the analyses concerning differences in the levels of the three domains of sexual quality of life between individuals with and without SCI, and across gender groups, the skewness and kurtosis values for all sexual quality of life variables remained within the |2| range, despite the Shapiro–Wilk test yielding significant results. Therefore, it was assumed that the distributions of sexual quality of life variables by gender and disability status approximate a normal distribution [130]. As a result, a parametric test-specifically, a two-way MANOVA-was employed to examine the effects of gender and health status on sexual quality of life.

Comparisons of nominal and ordinal variables between groups were conducted using the Mann–Whitney U test, the Chi-square test of independence, and Fisher’s exact test. In addition, Spearman’s rho correlation analyses were performed to determine the relationships between variables. To assess the effect size and the strength of the relationship between independent and dependent variables, the following significance thresholds were applied: p < 0.001, p < 0.01, and p < 0.05.

Effect sizes were estimated and interpreted based on the following correlation values: 0.00–0.19 – very weak, 0.20–0.39 – weak, 0.40–0.59 – moderate, 0.60–0.79 – strong, 0.80–1.00 – very strong [131].

The quantitative analysis of group characteristics and normality tests was conducted on a sample of 110 participants. However, calculations and analyses concerning primary problems following spinal cord injury (SCI) were carried out on a subgroup of 91 individuals, and subsequent comparative analyses involving other variables were performed on a group of 81 participants. The variation in sample size was due to missing data.

Statistical analyses were conducted in the following steps:

1. Comparison of variables describing group characteristics of men and women was performed using the Mann– Whitney U test (for time since injury and age) and the chi-square test of independence with Fisher’s exact test (see Table 1).
2. Distributions of quantitative variables were examined by calculating basic descriptive statistics and conducting the Shapiro–Wilk test to assess normality.
3. Arithmetic means were calculated for the number and types of primary sexual problems after SCI. These calculations were performed based on gender, as well as the extent and severity of the injury (AIS A vs. AIS B, C, D).
4. It was examined whether the extent and severity of spinal cord injury influenced changes in sexual activity, interest, and satisfaction. These differences were analyzed separately for men and women using the Mann– Whitney U test.
5. Spearman’s rho correlation analysis was conducted to examine the relationship between:

– age and time since injury and the severity of primary sexual dysfunction, depending on gender and the level and severity of spinal cord injury;
– primary, secondary, and tertiary symptoms of sexual dysfunction and the levels of the three domains of sexual quality of life after SCI-sexual activity (SxARS), sexual interest (SIRS), and sexual satisfaction (SSWS)-were analyzed using Spearman’s rho correlation. The analysis was conducted by gender, type of dysfunction, and AIS scale results (AIS A vs. AIS B, C, D). The correlation between menstrual disruption (as a primary symptom of sexual dysfunction after SCI) and sexual activity, interest, and satisfaction was excluded from the analysis due to the low prevalence of this issue among the studied women (83.3% did not experience this symptom). Due to the large discrepancy in group sizes (Table 1), it was not possible to perform a comparative analysis of the mean scores between men and women-whether single, in informal relationships, or in formal partnerships-with regard to sexual interest, activity, and satisfaction;
– sex, age, and time since injury and the three domains of sexual quality of life after SCI;
– the three domains of sexual quality of life, broken down by sex, type and severity of injury (AIS A vs. AIS B, C, D).

6. The distributions of variables were re-examined by performing an analysis of basic descriptive statistics, along with the Shapiro–Wilk test for normality, for age, time since injury, and the domains of sexual quality of life, based on sex and the extent and severity of injury (AIS A vs. AIS B) among men. The division of the female group by ASIA scale was omitted due to the small sample size.
7. A two-way MANOVA was conducted to analyze the differences in the three domains of sexual quality of life between individuals with spinal cord injury and able-bodied individuals, taking gender into account.

The analyses were performed using IBM SPSS Statistics, version 29.

## 4. Results

### 4.1. Analysis of sexual problems in individuals with Spinal cord injury

#### 4.1.1. Primary level of sexual problems in individuals after spinal cord injury

To answer the first research question, the frequency of primary-level sexual problems in patients after spinal cord injury (SCI) was calculated. These data were collected using the Female and Male Sexual and Reproductive Function Data Set and are presented in Tables 2-10.

**Table 2.**
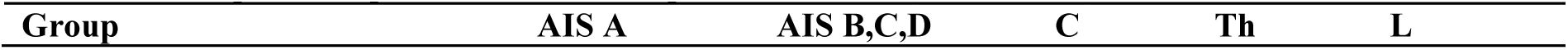
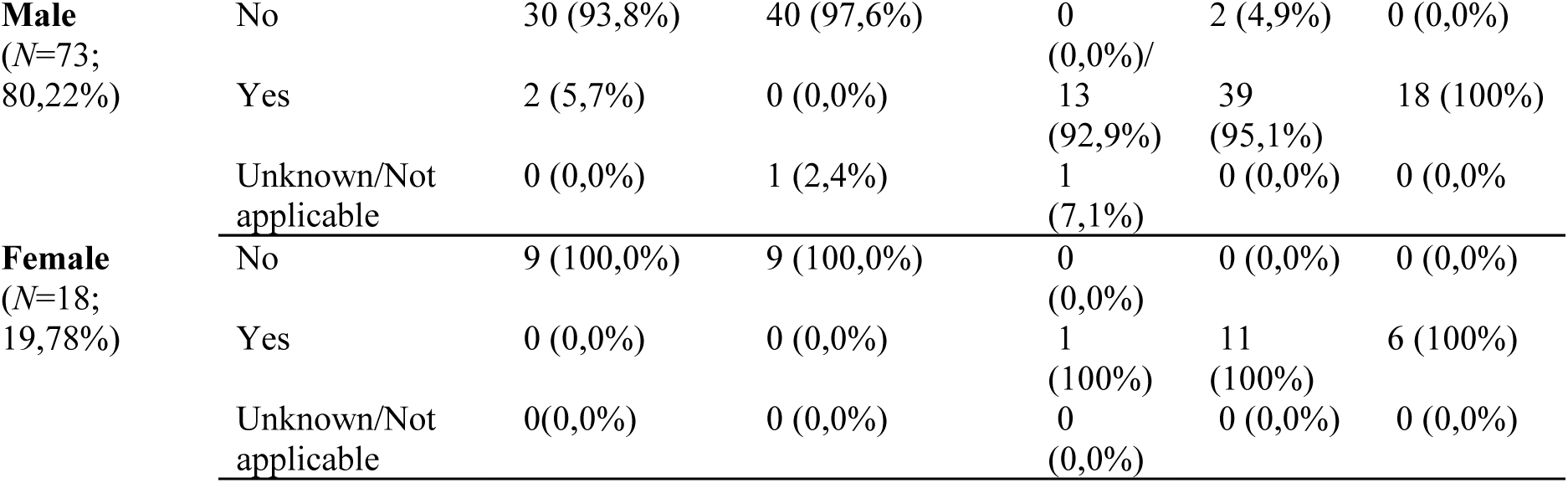
Sexual problems prior or unrelated to spinal cord lesion (SEXPRBSP).

| Group | AIS A | AIS B,C,D | C | Th | L |
| --- | --- | --- | --- | --- | --- |

|  |  |  |  |  |  |  |
| --- | --- | --- | --- | --- | --- | --- |
| **Male**<br>(N=73;<br>80,22%) | No | 30 (93,8%) | 40 (97,6%) | 0<br>(0,0%)/ | 2 (4,9%) | 0 (0,0%) |
|  | Yes | 2 (5,7%) | 0 (0,0%) | 13<br>(92,9%) | 39<br>(95,1%) | 18 (100%) |
|  | Unknown/Not applicable | 0 (0,0%) | 1 (2,4%) | 1<br>(7,1%) | 0 (0,0%) | 0 (0,0%) |
| **Female**<br>(N=18;<br>19,78%) | No | 9 (100,0%) | 9 (100,0%) | 0<br>(0,0%) | 0 (0,0%) | 0 (0,0%) |
|  | Yes | 0 (0,0%) | 0 (0,0%) | 1<br>(100%) | 11<br>(100%) | 6 (100%) |
|  | Unknown/Not applicable | 0(0,0%) | 0 (0,0%) | 0<br>(0,0%) | 0 (0,0%) | 0 (0,0%) |

**Table 3.** Sexual dysfunction related to the spinal cord lesion (SEXDYSCL).

| **Group** |  | **AIS A** | **AIS B,C,D** | **C** | **Th** | **L** |
| --- | --- | --- | --- | --- | --- | --- |
| **Male**<br>(N=73;<br>80,22%) | Yes | 28 (87,5%) | 36 (87,8%) | 11<br>(78,6%) | 36<br>(87,8%) | 17<br>(94,4%) |
|  | No | 1 (3,1%) | 2 (4,9%) | 1 (7,1%) | 2 (4,9%) | 0 (0,0%) |
|  | Unknown/Not applicable | 3 (9,4%) | 3 (7,3%) | 2 (14,4%) | 3 (7,3%) | 1 (4,3%) |
| **Female**<br>(N=18;<br>19,78%) | Yes | 7 (77,8%) | 8 (88,9%) | 0 (0,0%) | 9<br>(81,8%) | 6 (100%) |
|  | No | 1 (11,1%) | 0 (0,0%) | 0 (0,0%) | 1 (9,1%) | 0 (0,0%) |
|  | Unknown/Not applicable | 1(11,1%) | 1 (11,1%) | 1 (100%) | 1 (9,1) | 0 (0,0%) |

**Table 4.** Orgasmic function (ORGSMFXN).

| **Group** |  | **AIS A** | **AIS B,C,D** | **C** | **Th** | **L** |
| --- | --- | --- | --- | --- | --- | --- |
| **Male**<br>(N=73;<br>80,22%) | Normal | 3 (9,4%) | 4 (9,8%) | 7(53,8%) | 5(15,2%) | 2(11,8%) |
|  | Reduced/altered | 6 (18,8%) | 16 (39,0%) | 0(0%) | 8(24,2%) | 7(41,2%) |
|  | Absent | 18 (56,3%) | 16 (39,0%) | 6(46,2%) | 20(60,6%) | 8(47,1%) |
|  | Unknown | 5 (15,6%) | 5 (12,2%) | 0(0%) | 0(0%) | 0(0%) |
| **Female**<br>(N=18;<br>19,78%) | Normal | 1 (11,1%) | 1 (11,1%) | 0(0%) | 1(10%) | 1(20%) |
|  | Reduced/altered | 4 (44,4%) | 1 (11,1%) | 0(0%) | 4(40%) | 1(20%) |
|  | Absent | 3 (33,3%) | 5 (55,6%) | 0(0%) | 5(50%) | 3(60%) |
|  | Unknown | 1(11,1%) | 2(22,2%) | 1(100%) | 0(0%) | 0(0%) |

**Table 5.** Female psychogenic genital arousal (PSYCAROU).

| **Group** |  | **AIS A** | **AIS B,C,D** | **C** | **Th** | **L** |
| --- | --- | --- | --- | --- | --- | --- |
| **Female**<br>(N=18;<br>19,78%) | Normal | 6 (66,7%) | 2 (22,2%) | 0(0%) | 7(63,6%) | 1(16,7%) |
|  | Reduced/altered | 3 (33,3%) | 7 (77,8%) | 1(100%) | 4(36,4%) | 5(83,3%) |
|  | Absent | 0 (0,0%) | 0 (0,0%) | 0(0%) | 0(%) | 0(%) |
|  | Unknown | 0 (0,0%) | 0 (0,0%) | 0(0%) | 0(%) | 0(%) |

**Table 6.** Female Reflex genital arousal (REFLAROU).

| **Group** |  | **AIS A** | **AIS B,C,D** | **C** | **Th** | **L** |
| --- | --- | --- | --- | --- | --- | --- |
| **Female**<br>(N=18;<br>19,78%) | Normal | 4 (44,4%) | 1 (11,1%) | 0(0%) | 4(36,4%) | 1(16,67%) |
|  | Reduced/altered | 3 (33,3%) | 4 (44,4%) | 0(0%) | 5(45,5%) | 2(33,33%) |
|  | Absent | 2 (22,2%) | 2 (22,2%) | 0(0%) | 2(18,2%) | 2(33,33%) |
|  | Unknown | 0(0,0%) | 2 (22,2%) | 1(100%) | 0(0%) | 1(16,67%) |

**Table 7.** Menstruation (MENSTRUA).

| **Group** |  | **AIS A** | **AIS B,C,D** | **C** | **Th** | **L** |
| --- | --- | --- | --- | --- | --- | --- |
| **Female**<br>(N=18;<br>19,78%) | Normal | 7 (77,8%) | 7 (77,8%) | 1(100%) | 9(81,8%) | 4(66,7%) |
|  | Reduced/altered | 0 (0,0%) | 0 (0,0%) | 0(0%) | 0(0%) | 0(0%) |
|  | Absent | 0 (0,0%) | 1 (11,1%) | 0(0%) | 0(0%) | 1(16,7%) |
|  | Unknown | 2(22,2%) | 1 (11,1%) | 0(0%) | 2(18,2%) | 1(16,7%) |

**Table 8.**
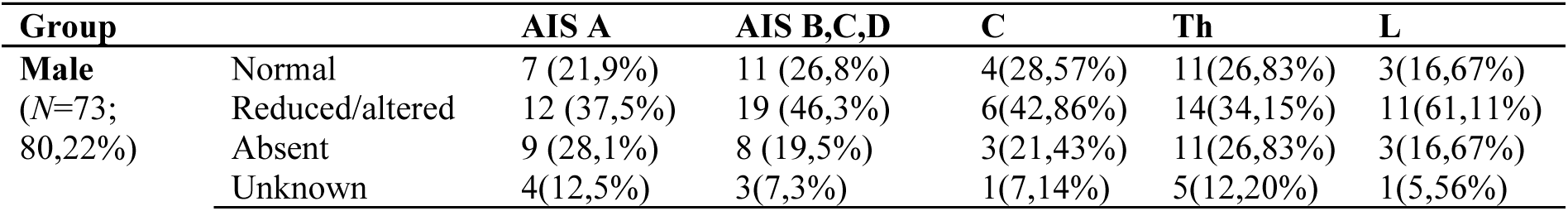
Psychogenic erection (PSYCEREC).

**Table 9.**
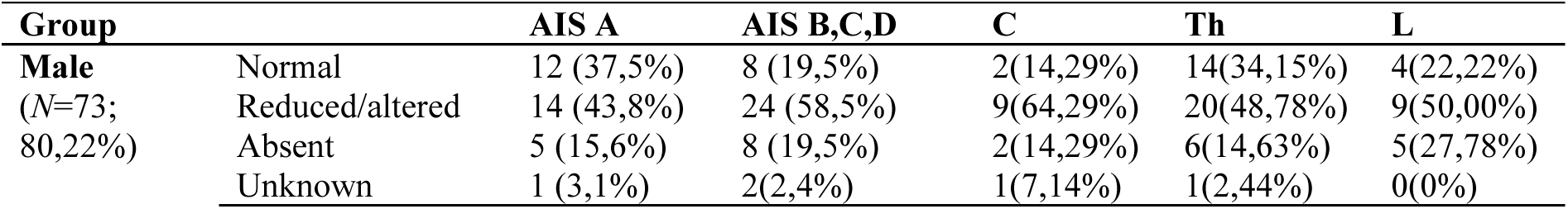
Reflex erection (REFLEREC).

**Table 10.**
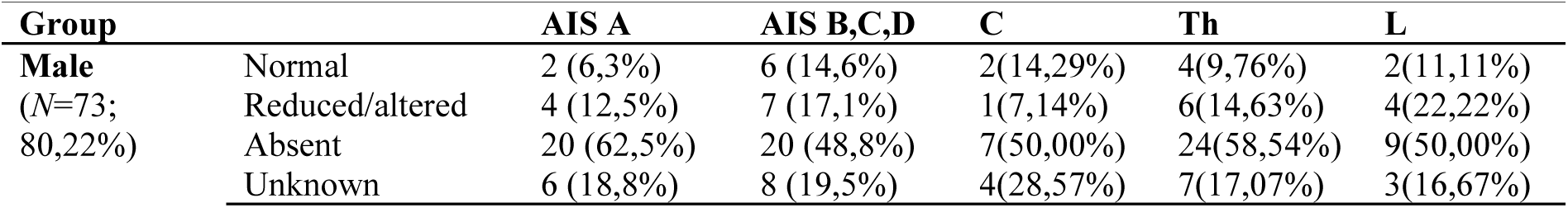
Ejaculation (EJACULAT).

The collected data indicate that the majority of patients reported sexual problems related to spinal cord injury, such as impaired or absent reflex and psychogenic erections and ejaculation in men, as well as impaired or absent psychogenic and reflex genital arousal and menstruation in women, along with reduced or absent orgasm in both sexes. Among men, this percentage was 87.65% (87.5% in the AIS A group and 87.8% in the AIS B, C, D group), and among women, it was 83.35% (77.8% in the AIS A group and 88.9% in the AIS B, C, D group). None of the women and the majority of men (N = 73) reported having sexual problems prior to the spinal cord injury (Table 3).

#### 4.1.2. Extent and severity of injury, age, and time since SCI in relation to the type, frequency, and severity of primary sexual problems following spinal cord injury

To determine whether age and time since injury are associated with the severity of sex-specific sexual dysfunctions in relation to the degree and completeness of spinal cord injury, the first step involved analyzing basic descriptive statistics along with the Shapiro-Wilk test results for time since injury, age, and primary sexual symptoms after SCI. The analysis was conducted by sex and according to AIS classification (AIS A vs. AIS B, C, D). The results are presented in Tables 11 and 12.

**Table 11.**
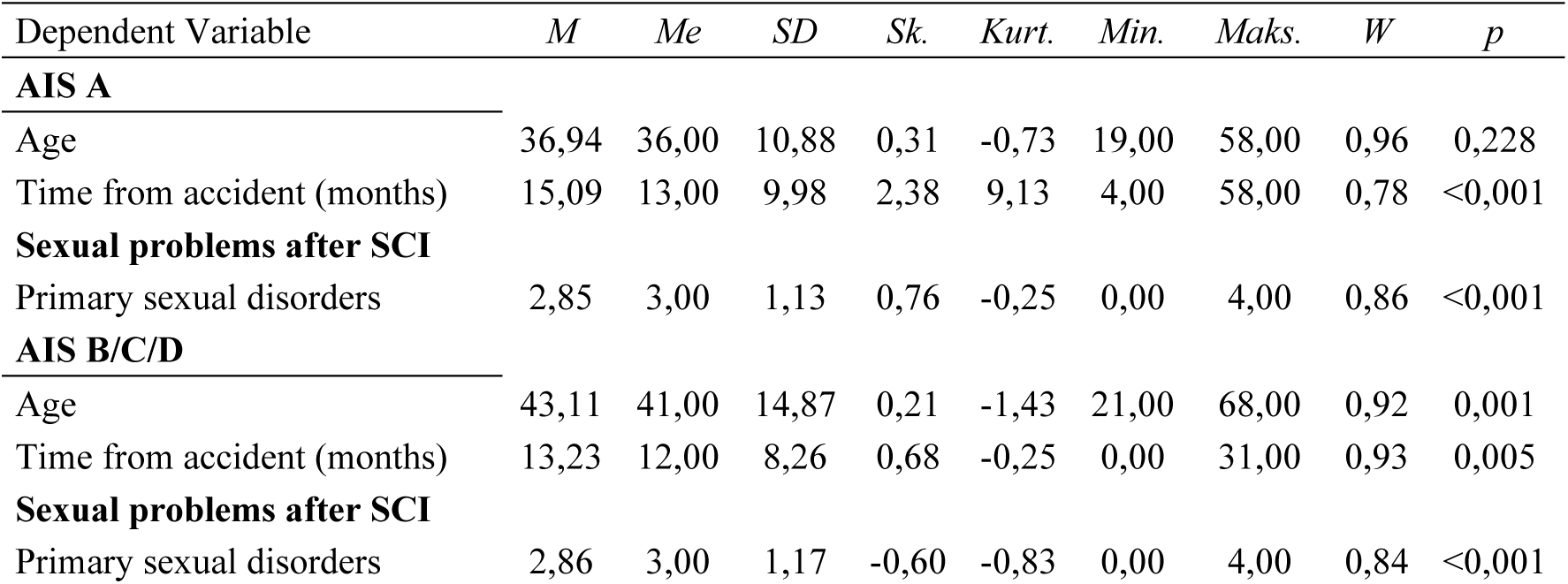
Basic descriptive statistics with the result of Shapiro-Wilk test in the group of men.

**Table 12.**
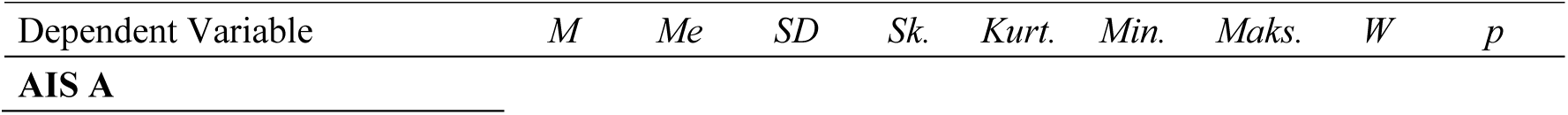
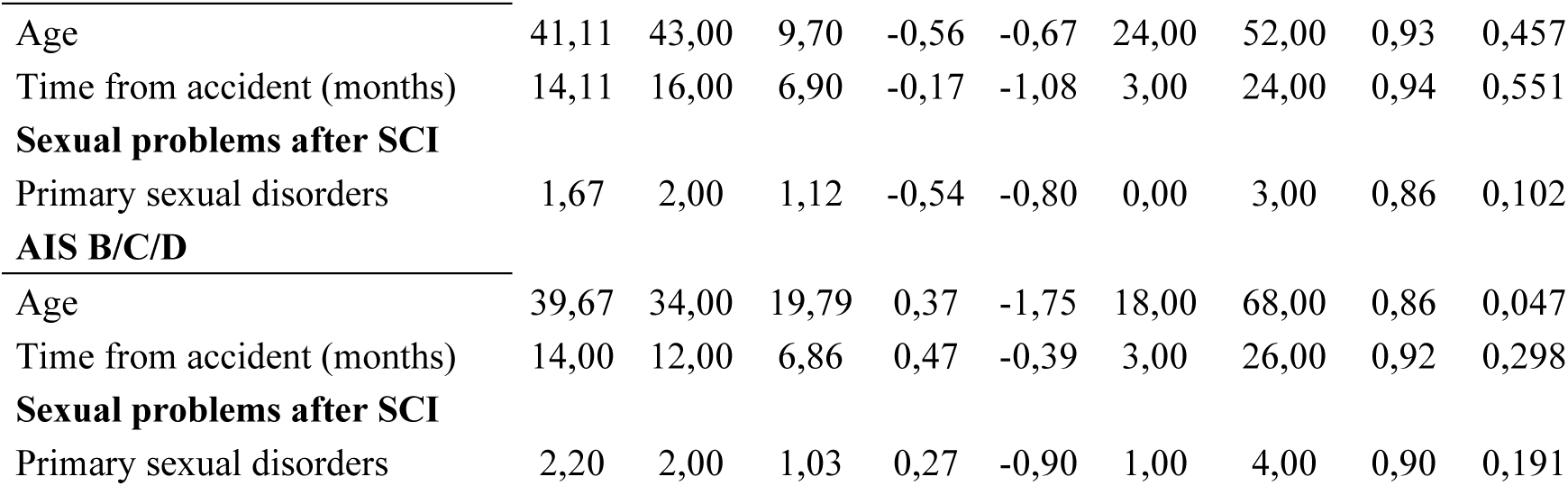
Basic descriptive statistics with the result of Shapiro-Wilk test in the group of women. Dependent Variable *M Me SD Sk. Kurt. Min. Maks. W p* AIS A.

|  |  |  |  |  |  |  |  |  |  |
| --- | --- | --- | --- | --- | --- | --- | --- | --- | --- |
| Age | 41,11 | 43,00 | 9,70 | -0,56 | -0,67 | 24,00 | 52,00 | 0,93 | 0,457 |
| Time from accident (months) | 14,11 | 16,00 | 6,90 | -0,17 | -1,08 | 3,00 | 24,00 | 0,94 | 0,551 |
| **Sexual problems after SCI** |  |  |  |  |  |  |  |  |  |
| Primary sexual disorders | 1,67 | 2,00 | 1,12 | -0,54 | -0,80 | 0,00 | 3,00 | 0,86 | 0,102 |
| **AIS B/C/D** |  |  |  |  |  |  |  |  |  |
| Age | 39,67 | 34,00 | 19,79 | 0,37 | -1,75 | 18,00 | 68,00 | 0,86 | 0,047 |
| Time from accident (months) | 14,00 | 12,00 | 6,86 | 0,47 | -0,39 | 3,00 | 26,00 | 0,92 | 0,298 |
| **Sexual problems after SCI** |  |  |  |  |  |  |  |  |  |
| Primary sexual disorders | 2,20 | 2,00 | 1,03 | 0,27 | -0,90 | 1,00 | 4,00 | 0,90 | 0,191 |

In the next step, a correlation analysis using Spearman’s rho coefficient was conducted. The results of this analysis are presented in Table 13.

**Table 13.**
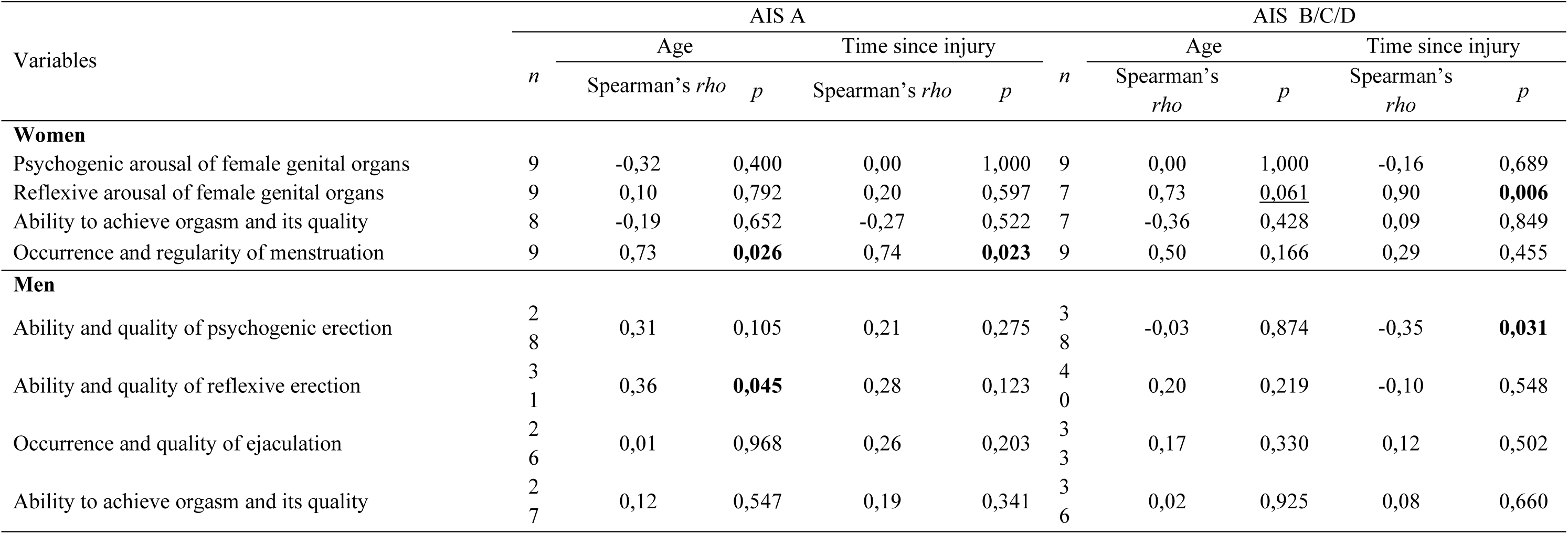
Results of Spearman’s rho correlation analysis between age and time since injury and gender-specific sexual dysfunctions, divided by degree of impairment.

| Variables | AIS A |  |  |  |  | AIS B/C/D |  |  |  |  |
| --- | --- | --- | --- | --- | --- | --- | --- | --- | --- | --- |
|  | *n* | Age |  | Time since injury |  | *n* | Age |  | Time since injury |  |
|  |  | Spearman's *rho* | *p* | Spearman's *rho* | *p* |  | Spearman's *rho* | *p* | Spearman's *rho* | *p* |
| **Women** |  |  |  |  |  |  |  |  |  |  |
| Psychogenic arousal of female genital organs | 9 | -0,32 | 0,400 | 0,00 | 1,000 | 9 | 0,00 | 1,000 | -0,16 | 0,689 |
| Reflexive arousal of female genital organs | 9 | 0,10 | 0,792 | 0,20 | 0,597 | 7 | 0,73 | <u>0,061</u> | 0,90 | **0,006** |
| Ability to achieve orgasm and its quality | 8 | -0,19 | 0,652 | -0,27 | 0,522 | 7 | -0,36 | 0,428 | 0,09 | 0,849 |
| Occurrence and regularity of menstruation | 9 | 0,73 | **0,026** | 0,74 | **0,023** | 9 | 0,50 | 0,166 | 0,29 | 0,455 |
| **Men** |  |  |  |  |  |  |  |  |  |  |
| Ability and quality of psychogenic erection | 2<br>8 | 0,31 | 0,105 | 0,21 | 0,275 | 3<br>8 | -0,03 | 0,874 | -0,35 | **0,031** |
| Ability and quality of reflexive erection | 3<br>1 | 0,36 | **0,045** | 0,28 | 0,123 | 4<br>0 | 0,20 | 0,219 | -0,10 | 0,548 |
| Occurrence and quality of ejaculation | 2<br>6 | 0,01 | 0,968 | 0,26 | 0,203 | 3<br>3 | 0,17 | 0,330 | 0,12 | 0,502 |
| Ability to achieve orgasm and its quality | 2<br>7 | 0,12 | 0,547 | 0,19 | 0,341 | 3<br>6 | 0,02 | 0,925 | 0,08 | 0,660 |

The conducted analyses showed that among women with SCI at AIS A level, the occurrence and regularity of menstruation were positively associated with age (*rho* = 0.73, *p* = 0.026) and time since injury (*rho* = 0.74, *p* = 0.023). These correlations were very strong; however, it is worth noting the small group size (*N* = 18). In the group of women with spinal cord injury at AIS B/C/D level, significant, positive, and very strong correlations were observed between reflexive sexual arousal of the genital organs and the following variables: time since injury (*rho* = 0.90, *p* = 0.006) and age (*rho* = 0.73, *p* = 0.061).

In the group of men with spinal cord injury at the AIS A level, a single positive correlation was found between age and the ability and quality of achieving reflexive erection (moderate correlation, *rho* = 0.36, *p* = 0.045). Among men with SCI at the AIS B/C/D level, significant negative correlations were observed between time since injury and the ability and quality of psychogenic erection (*rho* = -0.35, *p* = 0.031). These correlations were of moderate strength.

#### 4.1.3. Secondary level of sexual dysfunction symptoms after spinal cord injury

To determine the scope and level of secondary symptoms potentially related to sexual problems after SCI, an analysis of basic descriptive statistics was conducted using the Shapiro-Wilk test. **Tables 14 and 15** present the results of the assessment of secondary symptoms after SCI that may affect sexuality, i.e.: evaluations of walking aid performance (*Walking Index for Spinal Cord Injury – WISCI*), assessments of the level of functional independence of patients after SCI (*Spinal Cord Independence Measure – SCIM*), pain level (*Numeric Rating Scale for Pain – NRS Pain*), and spasticity level (*Ashworth Scale*). The results are presented separately for women and men, broken down by injury level and its scope and severity according to the AIS classification.

**Table 14.** Basic descriptive statistics with the result of Shapiro-Wilk test in the group of men.

| Dependent variable | *M* | *Me* | *SD* | *Sk.* | *Kurt.* | *Min.* | *Maks.* | *W* | *p* |
| --- | --- | --- | --- | --- | --- | --- | --- | --- | --- |
| **AIS A** |  |  |  |  |  |  |  |  |  |
| **Sexual problems after SCI** |  |  |  |  |  |  |  |  |  |
| Primary sexual disorders | 2,85 | 3,00 | 1,13 | 0,76 | -0,25 | 0,00 | 4,00 | 0,86 | <0,001 |
| WISCI | 1,36 | 0,00 | 3,00 | 2,54 | 6,58 | 0,00 | 13,00 | 0,53 | <0,001 |
| SCIM | 59,40 | 63,00 | 9,91 | -0,88 | 0,23 | 38,00 | 76,00 | 0,91 | 0,006 |
| NRS Pain | 3,49 | 2,00 | 2,86 | 0,79 | -0,85 | 1,00 | 9,00 | 0,81 | <0,001 |
| **AIS B/C/D** |  |  |  |  |  |  |  |  |  |
| **Sexual problems after SCI** |  |  |  |  |  |  |  |  |  |
| Primary sexual disorders | 2,86 | 3,00 | 1,17 | -0,60 | -0,83 | 0,00 | 4,00 | 0,84 | <0,001 |
| WISCI | 7,42 | 6,00 | 6,65 | 0,36 | -1,23 | 0,00 | 20,00 | 0,89 | <0,001 |
| SCIM | 66,66 | 69,00 | 20,35 | -0,50 | -0,42 | 20,00 | 105,00 | 0,96 | 0,113 |
| NRS Pain | 3,02 | 1,00 | 2,82 | 1,09 | -0,15 | 1,00 | 10,00 | 0,73 | <0,001 |

**Table 15.**
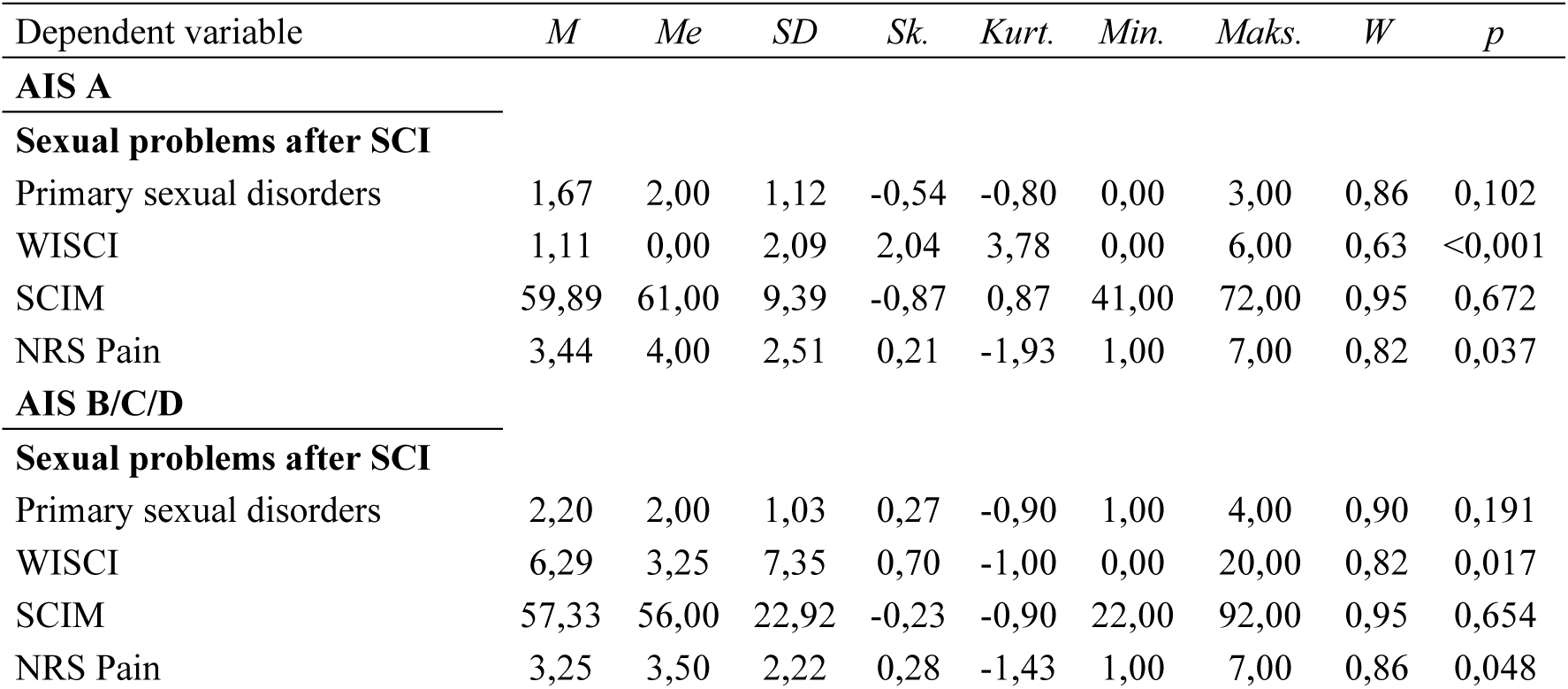
Basic descriptive statistics with the result of Shapiro-Wilk test in the group of women.

In terms of functional independence after SCI, the mean score in the group of men with complete SCI was M = 59.40, and in the group with incomplete SCI M = 66.66; in the group of women, the mean scores were M = 59.89 and M = 57.33, respectively, with the total score range spanning from 0 to 100.

In terms of walking function and the value of walking assistance, men and women with spinal cord injury classified as AIS B/C/D received a mean score of M = 7.42 and M = 6.29, respectively, while patients with complete injury (AIS A) scored M = 1.36 for men and M = 1.11 for women, with the total score range spanning from 0 to 20.

The average level of pain intensity in the group of women was 3.35 (i.e. 3.44 in the AIS A group and 3.25 in the AIS B, C, D group), and M = 3.25 in the group of men (i.e. M = 3.49 in the AIS A group and 3.02 in the AIS B, C, D group), with a scale range from 0 to 10. Based on this, it can be concluded that the average pain intensity in the studied groups was low.

#### 4.1.4. Tertiary level of symptoms of sexual problems after spinal cord injury

To examine the scope of selected tertiary symptoms after spinal cord injury (SCI) that may be related to sexuality, an analysis of basic descriptive statistics was conducted using the Shapiro–Wilk test. Tables 16 and 17 present the results for tertiary symptoms after SCI that may affect sexuality, including: level of state anxiety, depression, subjective sexual attractiveness scale, and domains of sexual quality of life. The results are presented separately for women and men, divided by the level, extent, and severity of spinal cord injury according to AIS classification.

**Table 16.**
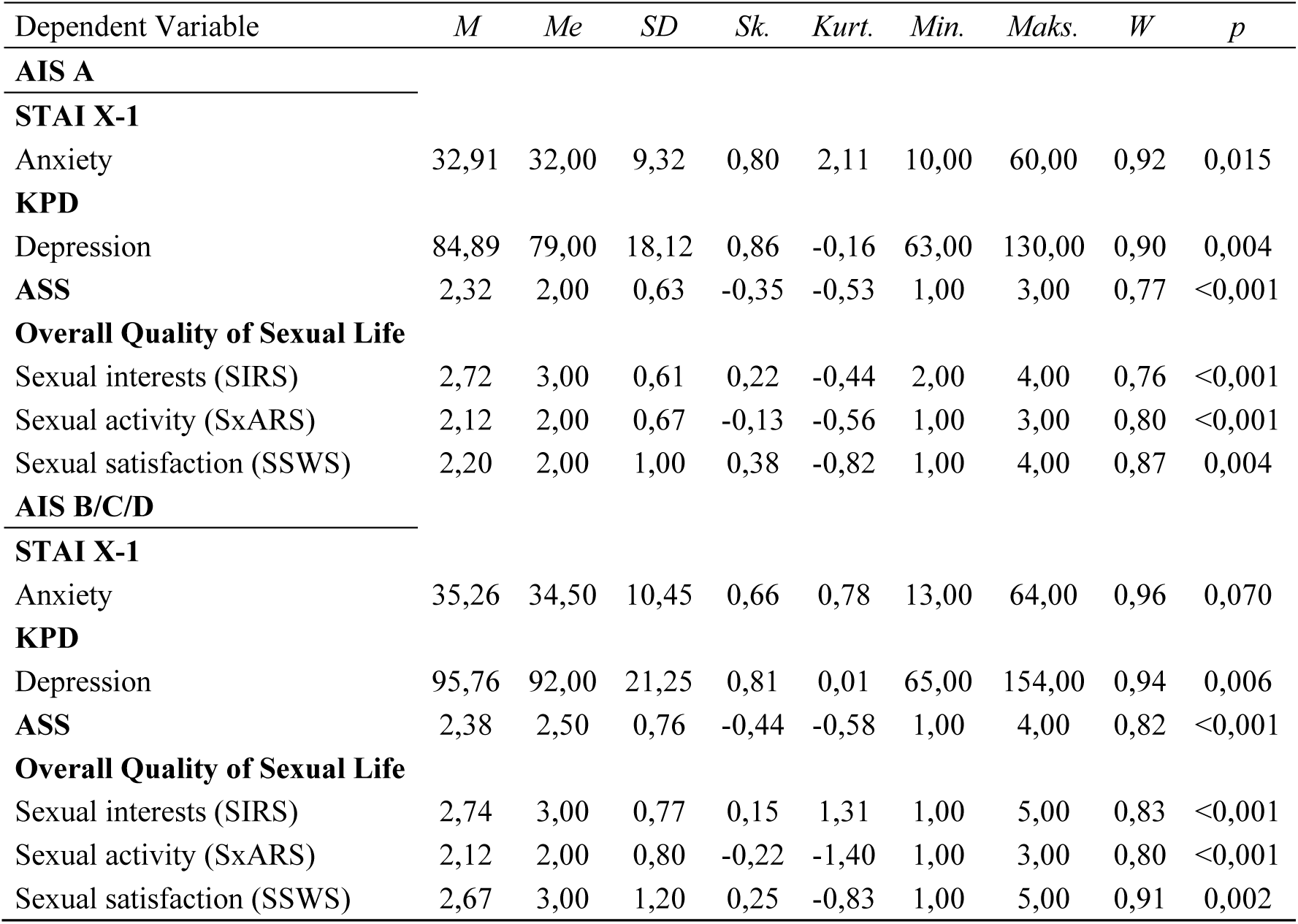
Basic descriptive statistics with the result of Shapiro-Wilk test in the group of men.

**Table 17.**
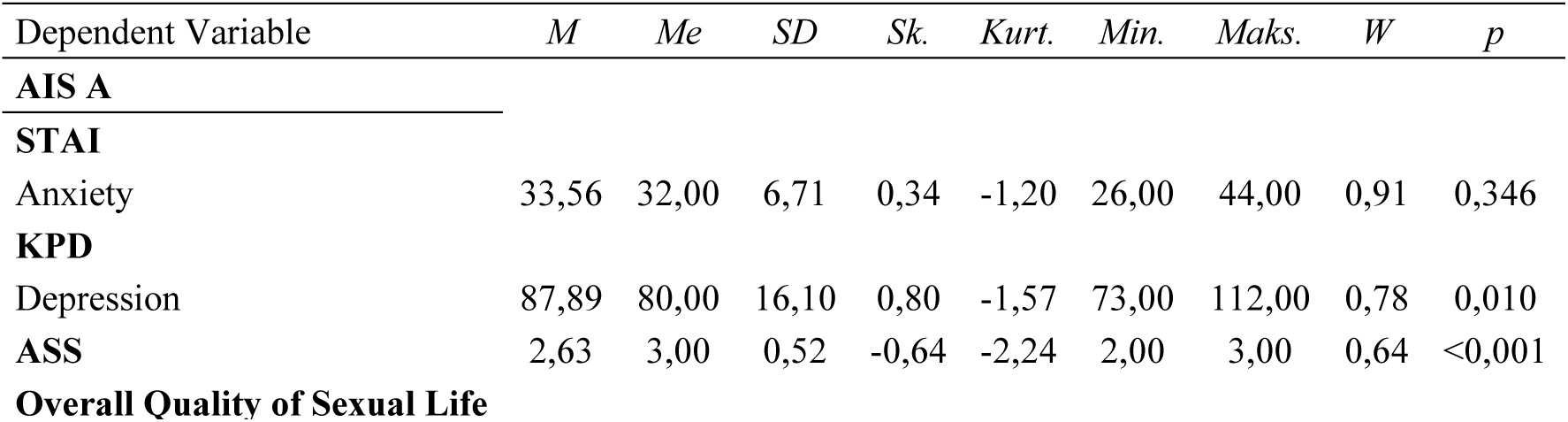
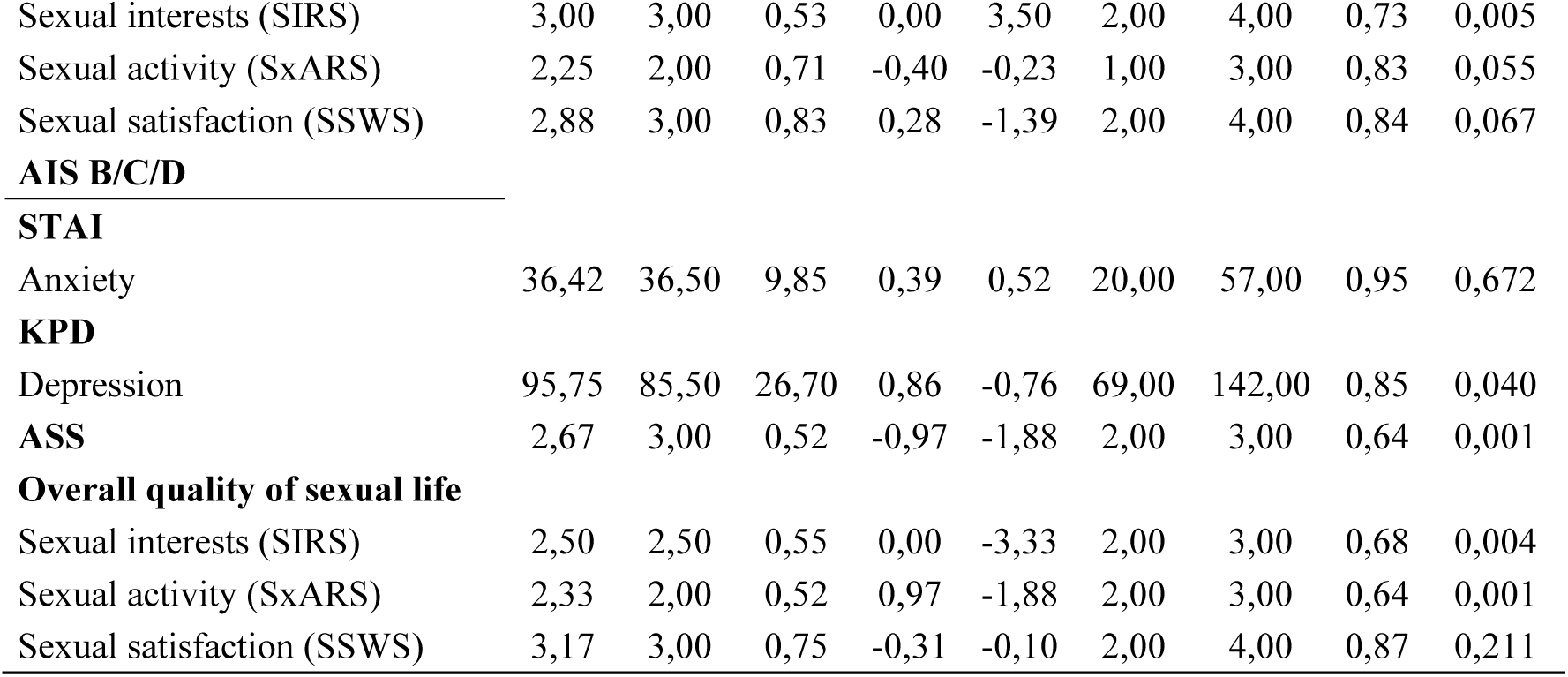
Basic descriptive statistics with the result of Shapiro-Wilk test in the group of women.

The data presented in Tables 16 and 17 indicate that both women and men, regardless of the extent and severity of injury (AIS A and AIS B/C/D), obtained average scores for symptoms of depression. However, it is worth noting that in the group of individuals with incomplete spinal cord injury (AIS B/C/D), there were participants whose depression symptom levels exceeded the clinical cut-off point for diagnosing depression, i.e., 130 points.

In terms of perceived sexual attractiveness, the average scores in the female group were: F AIS A: M = 2.32; F AIS B, C, D: M = 2.38, and in the male group: M AIS A: M = 2.63; M AIS B, C, D: M = 2.67, on a scale ranging from 1 to 5. For the domains of sexual quality of life, the highest levels of sexual interest were reported by both women and men with spinal cord injuries classified as AIS A (F: M = 2.72, M: M = 3.00, on a 5-point scale). In terms of sexual activity, most participants (both female and male, regardless of AIS classification) scored below the average value of the 5-point scale (range: 1–5). Regarding sexual satisfaction, the highest average was reported by men classified as AIS B, C, D (M = 3.17), and the lowest average by women with AIS A classification (M = 2.20). The score range for the scale was 1 to 5 points.

#### 4.1.5. The extent and severity of injury, age, and time since SCI in relation to the type, quantity, and intensity of secondary and tertiary problems potentially related to sexual dysfunction in individuals after spinal cord injury

To examine whether the extent and severity of injury, age, and time since SCI are related to the intensity of secondary and tertiary sexual problems after spinal cord injury, the first step was to assess the distributions of quantitative or ordinal variables measured on a Likert scale. Descriptive statistics and Shapiro-Wilk test results, broken down by gender as well as by the extent and severity of the injury, are presented in Table 18.

**Table 18.**
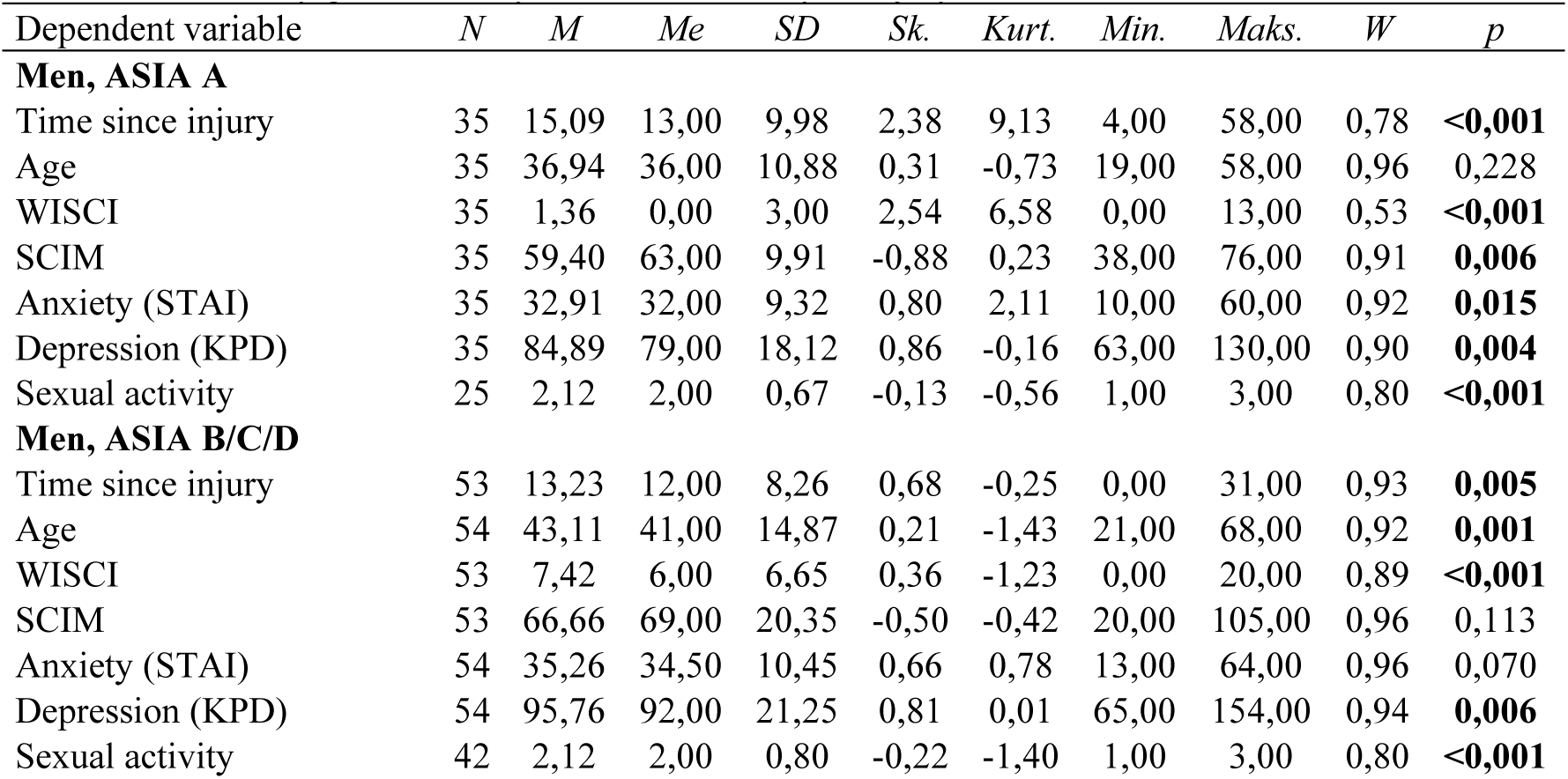
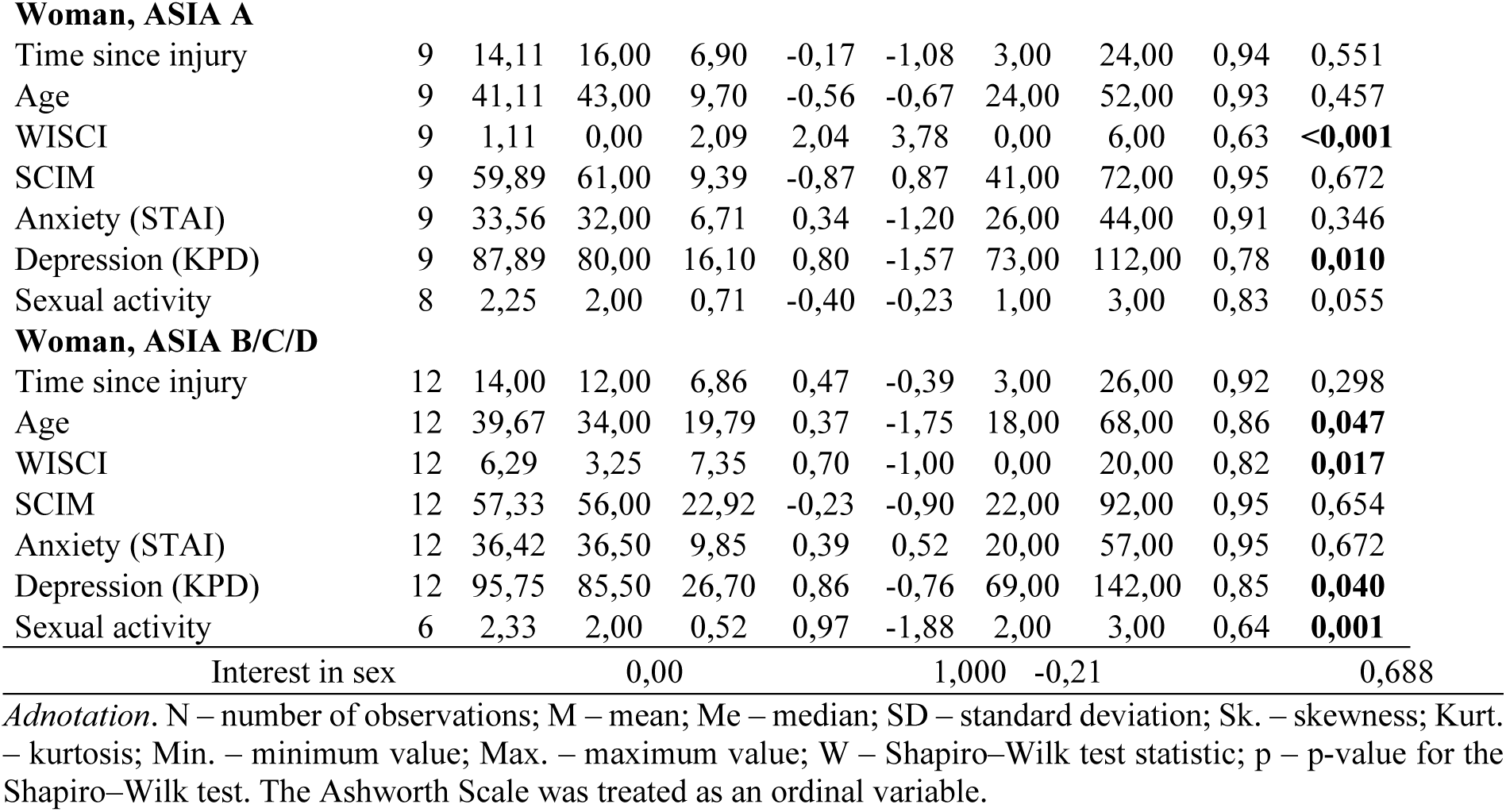
Descriptive statistics for time since injury and age, as well as secondary and tertiary symptoms after SCI, broken down by gender and by extent and severity of injury.

| Dependent variable | *N* | *M* | *Me* | *SD* | *Sk.* | *Kurt.* | *Min.* | *Maks.* | *W* | *p* |
| --- | --- | --- | --- | --- | --- | --- | --- | --- | --- | --- |
| **Men, ASIA A** |  |  |  |  |  |  |  |  |  |  |
| Time since injury | 35 | 15,09 | 13,00 | 9,98 | 2,38 | 9,13 | 4,00 | 58,00 | 0,78 | **<0,001** |
| Age | 35 | 36,94 | 36,00 | 10,88 | 0,31 | -0,73 | 19,00 | 58,00 | 0,96 | 0,228 |
| WISCI | 35 | 1,36 | 0,00 | 3,00 | 2,54 | 6,58 | 0,00 | 13,00 | 0,53 | **<0,001** |
| SCIM | 35 | 59,40 | 63,00 | 9,91 | -0,88 | 0,23 | 38,00 | 76,00 | 0,91 | **0,006** |
| Anxiety (STAI) | 35 | 32,91 | 32,00 | 9,32 | 0,80 | 2,11 | 10,00 | 60,00 | 0,92 | **0,015** |
| Depression (KPD) | 35 | 84,89 | 79,00 | 18,12 | 0,86 | -0,16 | 63,00 | 130,00 | 0,90 | **0,004** |
| Sexual activity | 25 | 2,12 | 2,00 | 0,67 | -0,13 | -0,56 | 1,00 | 3,00 | 0,80 | **<0,001** |
| **Men, ASIA B/C/D** |  |  |  |  |  |  |  |  |  |  |
| Time since injury | 53 | 13,23 | 12,00 | 8,26 | 0,68 | -0,25 | 0,00 | 31,00 | 0,93 | **0,005** |
| Age | 54 | 43,11 | 41,00 | 14,87 | 0,21 | -1,43 | 21,00 | 68,00 | 0,92 | **0,001** |
| WISCI | 53 | 7,42 | 6,00 | 6,65 | 0,36 | -1,23 | 0,00 | 20,00 | 0,89 | **<0,001** |
| SCIM | 53 | 66,66 | 69,00 | 20,35 | -0,50 | -0,42 | 20,00 | 105,00 | 0,96 | 0,113 |
| Anxiety (STAI) | 54 | 35,26 | 34,50 | 10,45 | 0,66 | 0,78 | 13,00 | 64,00 | 0,96 | 0,070 |
| Depression (KPD) | 54 | 95,76 | 92,00 | 21,25 | 0,81 | 0,01 | 65,00 | 154,00 | 0,94 | **0,006** |
| Sexual activity | 42 | 2,12 | 2,00 | 0,80 | -0,22 | -1,40 | 1,00 | 3,00 | 0,80 | **<0,001** |
| **Woman, ASIA A** |  |  |  |  |  |  |  |  |  |  |
| Time since injury | 9 | 14,11 | 16,00 | 6,90 | -0,17 | -1,08 | 3,00 | 24,00 | 0,94 | 0,551 |
| Age | 9 | 41,11 | 43,00 | 9,70 | -0,56 | -0,67 | 24,00 | 52,00 | 0,93 | 0,457 |
| WISCI | 9 | 1,11 | 0,00 | 2,09 | 2,04 | 3,78 | 0,00 | 6,00 | 0,63 | **<0,001** |
| SCIM | 9 | 59,89 | 61,00 | 9,39 | -0,87 | 0,87 | 41,00 | 72,00 | 0,95 | 0,672 |
| Anxiety (STAI) | 9 | 33,56 | 32,00 | 6,71 | 0,34 | -1,20 | 26,00 | 44,00 | 0,91 | 0,346 |
| Depression (KPD) | 9 | 87,89 | 80,00 | 16,10 | 0,80 | -1,57 | 73,00 | 112,00 | 0,78 | **0,010** |
| Sexual activity | 8 | 2,25 | 2,00 | 0,71 | -0,40 | -0,23 | 1,00 | 3,00 | 0,83 | 0,055 |
| **Woman, ASIA B/C/D** |  |  |  |  |  |  |  |  |  |  |
| Time since injury | 12 | 14,00 | 12,00 | 6,86 | 0,47 | -0,39 | 3,00 | 26,00 | 0,92 | 0,298 |
| Age | 12 | 39,67 | 34,00 | 19,79 | 0,37 | -1,75 | 18,00 | 68,00 | 0,86 | **0,047** |
| WISCI | 12 | 6,29 | 3,25 | 7,35 | 0,70 | -1,00 | 0,00 | 20,00 | 0,82 | **0,017** |
| SCIM | 12 | 57,33 | 56,00 | 22,92 | -0,23 | -0,90 | 22,00 | 92,00 | 0,95 | 0,654 |
| Anxiety (STAI) | 12 | 36,42 | 36,50 | 9,85 | 0,39 | 0,52 | 20,00 | 57,00 | 0,95 | 0,672 |
| Depression (KPD) | 12 | 95,75 | 85,50 | 26,70 | 0,86 | -0,76 | 69,00 | 142,00 | 0,85 | **0,040** |
| Sexual activity | 6 | 2,33 | 2,00 | 0,52 | 0,97 | -1,88 | 2,00 | 3,00 | 0,64 | **0,001** |

Interest in sex 0,00 1,000 -0,21 0,688
*Adnotation.* N – number of observations; M – mean; Me – median; SD – standard deviation; Sk. – skewness; Kurt. – kurtosis; Min. – minimum value; Max. – maximum value; W – Shapiro–Wilk test statistic; p – p-value for the Shapiro–Wilk test. The Ashworth Scale was treated as an ordinal variable.

The results of the Shapiro–Wilk test indicate distributions that are both similar (p > 0.05) and dissimilar (p < 0.05) to the normal distribution. Nevertheless, some variables showed very high skewness and kurtosis values (> |2|). Therefore, dependency analyses were based on non-parametric tests – specifically, Spearman’s rho correlation analysis.

The relationship between age and time since injury and the severity of secondary symptoms after SCI

To determine whether age and time since SCI are associated with the severity of secondary symptoms following SCI, a Spearman’s rho correlation analysis was conducted. The analysis was performed by dividing the sample by gender (women vs. men) and by the extent and severity of the injury (AIS A vs. AIS B/C/D). The results are reported in Table 19.

**Table 19.**
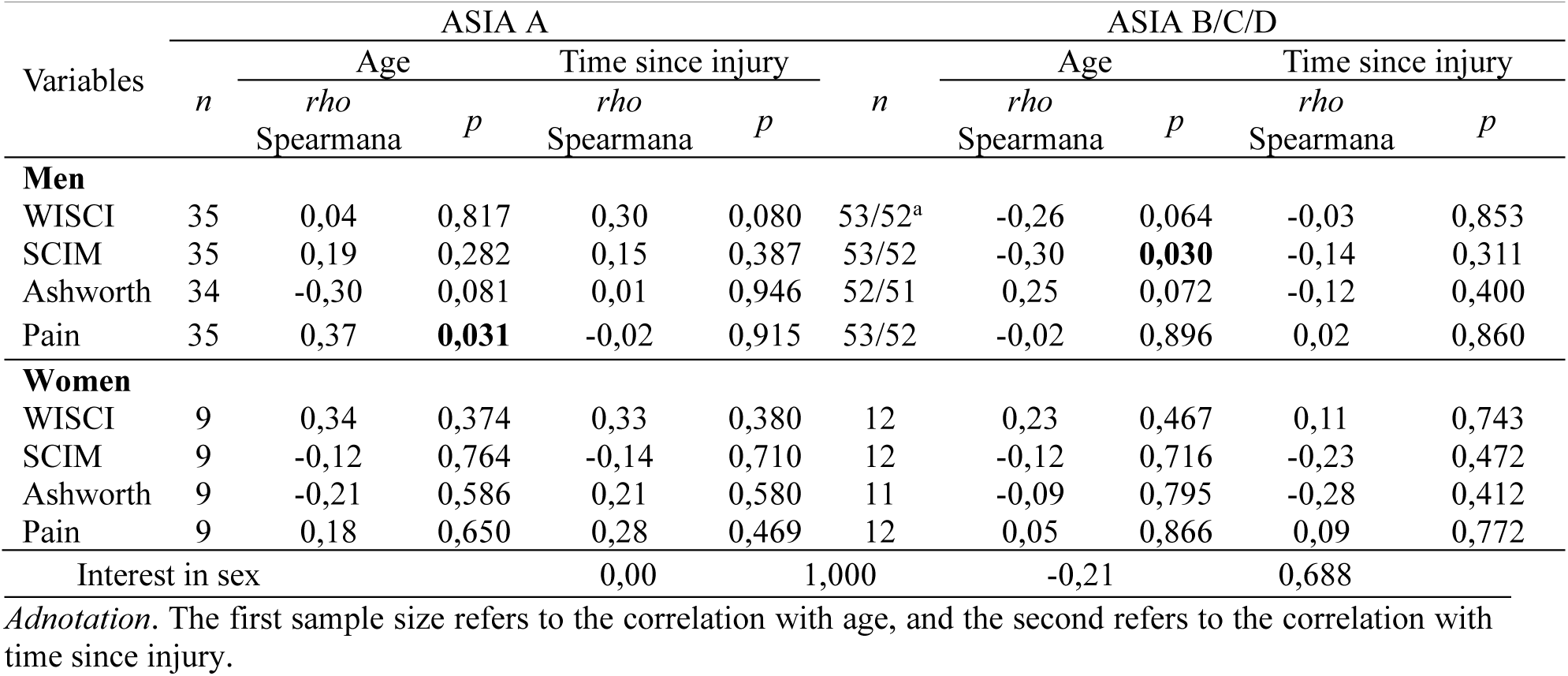
Results of Spearman’s rho correlation analysis between age and time since injury and the severity of secondary symptoms after SCI.

The analysis showed that in the group of men with spinal cord injury classified as AIS A, age was associated with experienced pain. This relationship was positive and of moderate strength (p = 0.031, Rho = -0.02). Other results were not statistically significant, although some correlations reached the level of statistical tendency. In this group, age was moderately and negatively correlated with the Ashworth Scale (Rho = -0.30, p = 0.081). A similar moderate positive relationship was observed between time since injury and the WISCI scale (Rho = 0.30, p = 0.080).

In the group of men classified as AIS B/C/D, a statistically significant correlation was found between age and the SCIM scale (a moderate negative relationship, Rho = -0.30, p = 0.030). Two additional correlations reached the level of statistical tendency: between age and the WISCI scale (a weak negative relationship, Rho = -0.26, p = 0.064) and between age and the Ashworth scale (a weak positive relationship, Rho = 0.25, p = 0.072). Other correlations in this group were not statistically significant.

In the groups of women classified as AIS A and AIS B/C/D, none of the relationships between age or time since injury and the severity of secondary symptoms after SCI were statistically significant. Moreover, no results reached the level of statistical tendency.

Relationship between age and time since injury and the severity of tertiary symptoms after SCI.

In the next step, the relationship between age and time since SCI and the level of tertiary symptoms after SCI was examined. The analysis was again conducted by dividing participants by gender and by the extent and completeness of the injury. The results of the Spearman’s rho correlation analysis are presented in Table 20.

**Table 20.**
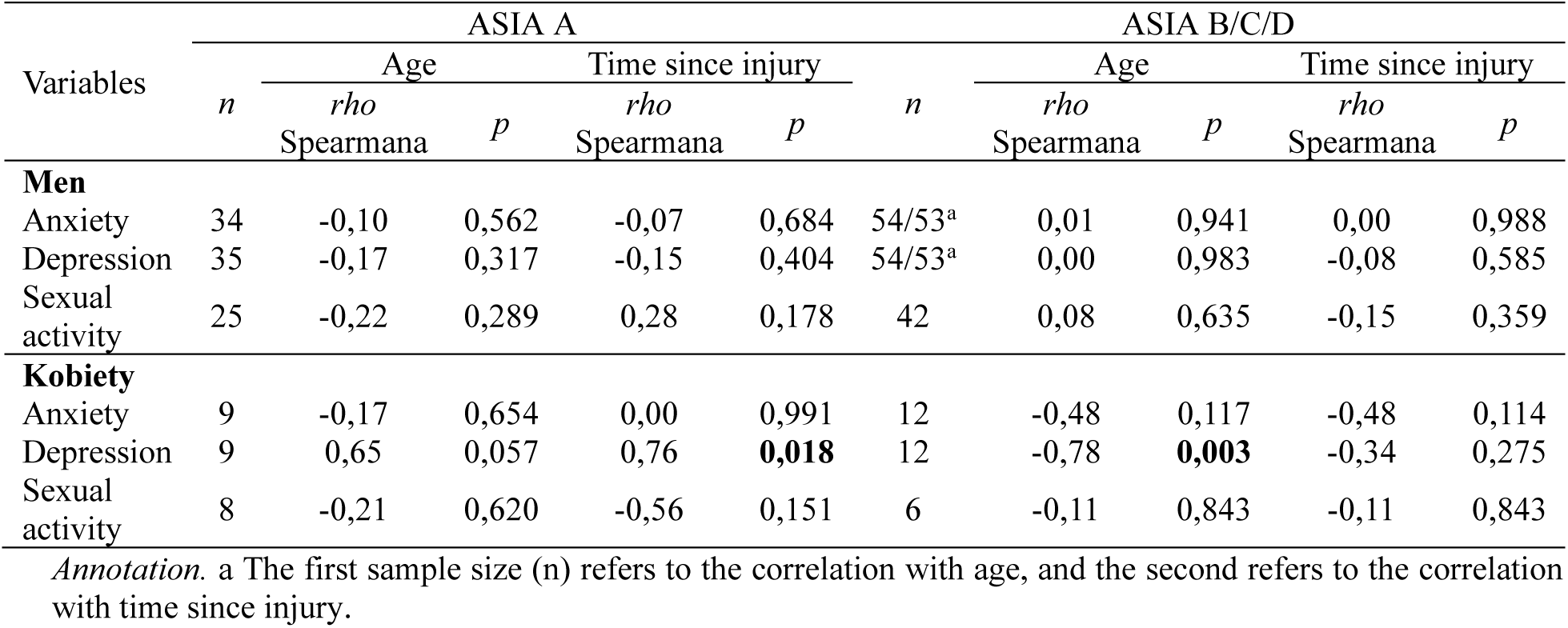
Results of Spearman’s rho correlation analysis between age and time since injury and tertiary symptoms after SCI.

| after SCI. |  |  |  |  |  |  |  |  |  |  |
| --- | --- | --- | --- | --- | --- | --- | --- | --- | --- | --- |
| Variables | ASIA A |  |  |  |  | ASIA B/C/D |  |  |  |  |
|  | *n* | Age |  | Time since injury |  | *n* | Age |  | Time since injury |  |
|  |  | *rho* |  | *rho* |  |  | *rho* |  | *rho* |  |
|  |  | Spearmana | *p* | Spearmana | *p* |  | Spearmana | *p* | Spearmana | *p* |
| **Men** |  |  |  |  |  |  |  |  |  |  |
| Anxiety | 34 | -0,10 | 0,562 | -0,07 | 0,684 | 54/53<sup>a</sup> | 0,01 | 0,941 | 0,00 | 0,988 |
| Depression | 35 | -0,17 | 0,317 | -0,15 | 0,404 | 54/53<sup>a</sup> | 0,00 | 0,983 | -0,08 | 0,585 |
| Sexual activity | 25 | -0,22 | 0,289 | 0,28 | 0,178 | 42 | 0,08 | 0,635 | -0,15 | 0,359 |
| **Kobiety** |  |  |  |  |  |  |  |  |  |  |
| Anxiety | 9 | -0,17 | 0,654 | 0,00 | 0,991 | 12 | -0,48 | 0,117 | -0,48 | 0,114 |
| Depression | 9 | 0,65 | 0,057 | 0,76 | **0,018** | 12 | -0,78 | **0,003** | -0,34 | 0,275 |
| Sexual activity | 8 | -0,21 | 0,620 | -0,56 | 0,151 | 6 | -0,11 | 0,843 | -0,11 | 0,843 |

*Annotation.* a The first sample size (*n*) refers to the correlation with age, and the second refers to the correlation with time since injury.

In the groups of men classified as AIS A and AIS B/C/D, the results were not statistically significant. This indicates that age and time since SCI were not associated with tertiary symptoms following SCI in the male group.

In the AIS A female group, a significant relationship was found between time since SCI and depression. This association was positive and very strong (Rho = 0.76, p = 0.018). A significant relationship was also observed between depression and age, with a strong positive correlation (Rho = 0.65, p = 0.057). Other correlations in this group were not statistically significant.

In the AIS B/C/D female group, the relationship between age and depression was statistically significant but showed an opposite trend compared to the AIS A female group. The strength of this correlation was very strong (Rho = -0.78, p = 0.003), indicating that depressive symptoms decrease with age.

### 4.2. The relationship between sexual problems after SCI and the domains of general sexual quality of life

#### 4.2.1 Dynamics of sexual Life domains in individuals after spinal cord injury (SCI)

##### 4.2.1.1 Relationships between domains of Sexual Quality of Life in Individuals After Spinal Cord Injury (SCI)

To answer the research question regarding the relationship between the three identified domains of sexual quality of life in individuals after spinal cord injury (SCI), a correlation analysis was conducted among these domains. Spearman’s rho test was used for this purpose. The results are presented in Table 21.

**Table 21.**
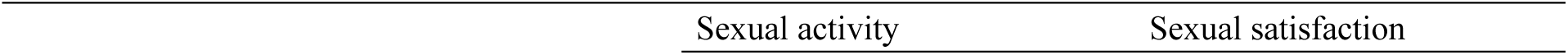
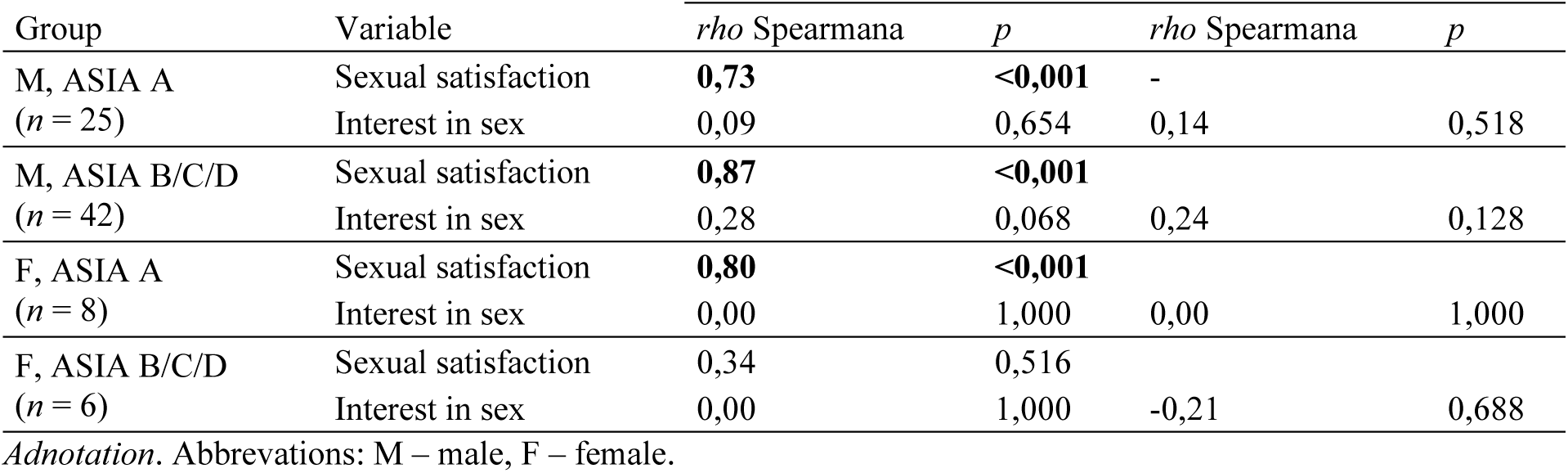
Results of Spearman’s rho correlation analisis between sexual activity, sexual satisfaction, and interest in sex.

| Group | Variable | *rho* Spearmana | *p* | *rho* Spearmana | *p* |
| --- | --- | --- | --- | --- | --- |
| M, ASIA A<br>(*n* = 25) | Sexual satisfaction | **0,73** | **<0,001** | - |  |
|  | Interest in sex | 0,09 | 0,654 | 0,14 | 0,518 |
| M, ASIA B/C/D<br>(*n* = 42) | Sexual satisfaction | **0,87** | **<0,001** |  |  |
|  | Interest in sex | 0,28 | 0,068 | 0,24 | 0,128 |
| F, ASIA A<br>(*n* = 8) | Sexual satisfaction | **0,80** | **<0,001** |  |  |
|  | Interest in sex | 0,00 | 1,000 | 0,00 | 1,000 |
| F, ASIA B/C/D<br>(*n* = 6) | Sexual satisfaction | 0,34 | 0,516 |  |  |
|  | Interest in sex | 0,00 | 1,000 | -0,21 | 0,688 |

*Adnotation.* Abbreviations: M – male, F – female.

The results showed that the higher the level of sexual activity, the higher the level of sexual satisfaction in the group of men (regardless of the ASIA classification) as well as in the group of women with complete spinal cord injury (strong correlation: in men with AIS A, rho=0,73; in men with AIS B/C/D, rho=0,87; in women with AIS A, rho = 0.80; p < 0.001 for all groups). This relationship was not observed among women with incomplete SCI (i.e., injuries classified as AIS B, C, or D). The data also indicated that sexual interest was not related to the level of sexual activity or sexual satisfaction.

##### 4.2.1.1 Differences in sexual activity, interest, and satisfaction depending on the classification of spinal cord injury, taking gender into account

Using the Mann-Whitney U test, it was examined whether the severity and extent of spinal cord injury differentiate patients in terms of the levels of general sexual quality of life domains. The results are presented in Table 22.

**Table 22.**
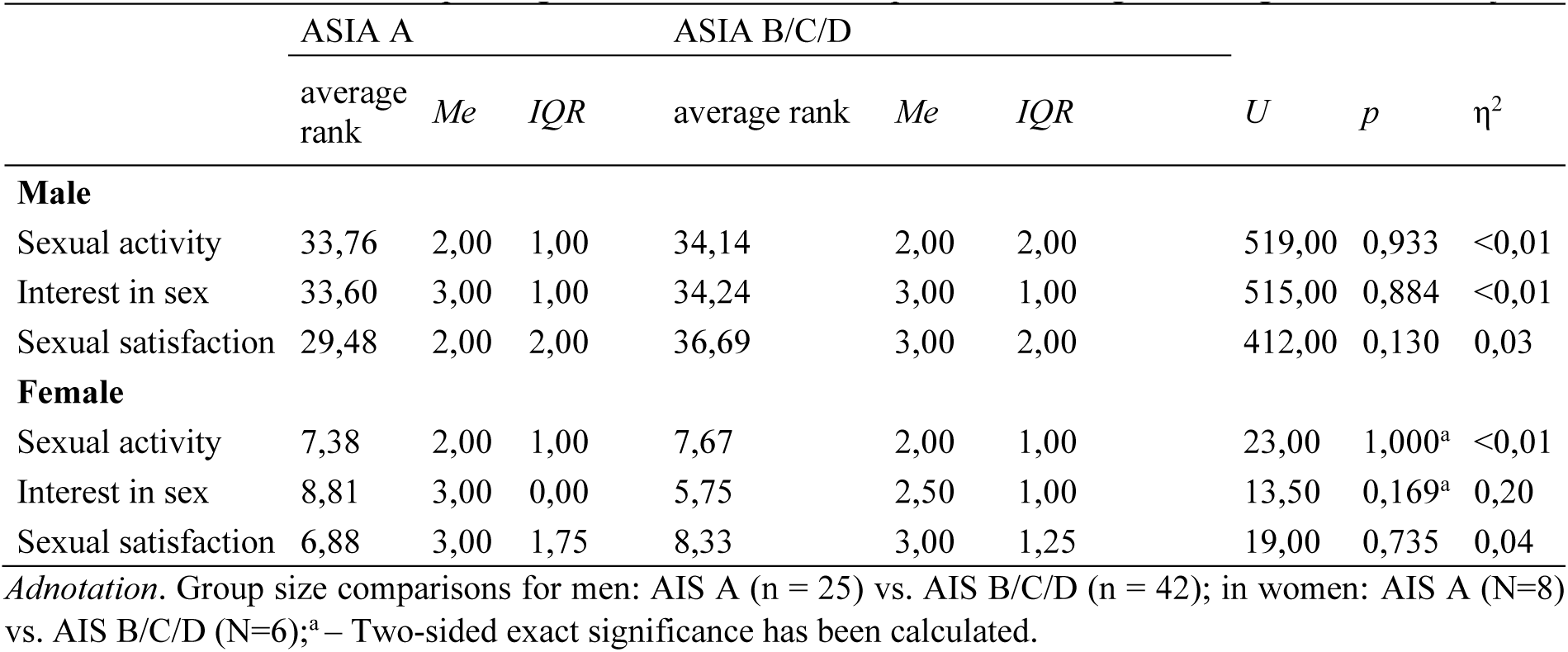
Results of comparative analysis using the Mann Whitney U test for changes in sexual activity, interest in sex and sexual satisfaction depending on the classification of spinal cord damage and the gender of the subjects.

The results showed that the severity and extent of injury (AIS A vs. AIS B/C/D) did not differentiate patients in terms of changes in the three domains of sexual quality of life. This effect was not observed in either gender.

##### 4.2.1.1 The relationship between changes in sexual activity, sexual interest, and sexual satisfaction and age and time since injury, by gender and spinal cord injury classification

In order to address the next research question concerning the relationship between age, time since spinal cord injury, and the domains of sexual life, a correlation analysis of these variables was conducted, taking into account gender and AIS injury classification. For this purpose, Spearman’s rho correlation test was used. The results are presented in Table 23.

**Table 23.**
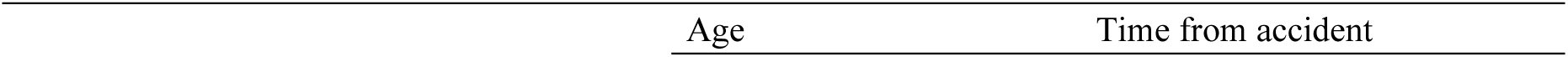
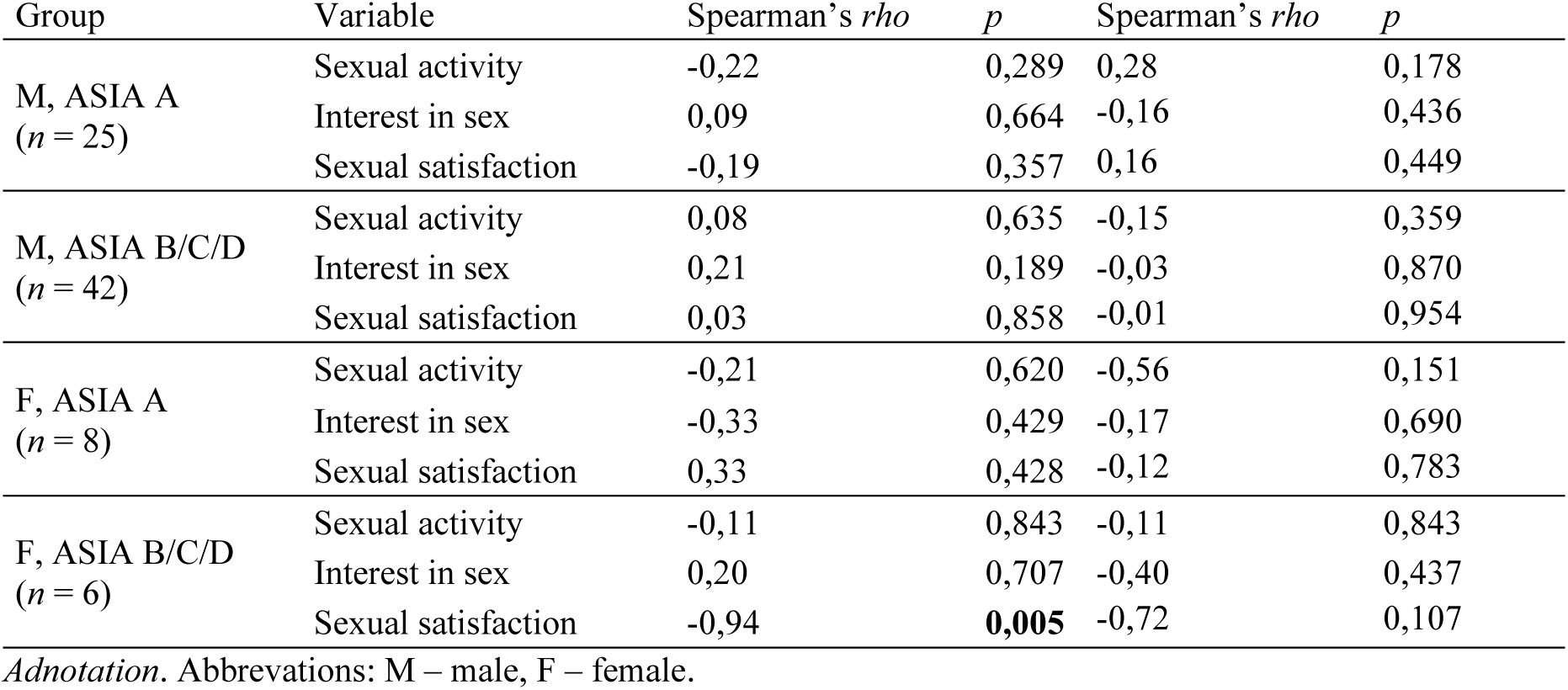
Spearman’s rho correlations between age and time since injury and changes in sexual activity, interest in sex, and satisfaction by gender and SCI classification.

| Age | Time from accident |
| --- | --- |

| Group | Variable | Spearman's $\rho$ | $p$ | Spearman's $\rho$ | $p$ |
| --- | --- | --- | --- | --- | --- |
| M, ASIA A<br>($n = 25$) | Sexual activity | -0,22 | 0,289 | 0,28 | 0,178 |
|  | Interest in sex | 0,09 | 0,664 | -0,16 | 0,436 |
|  | Sexual satisfaction | -0,19 | 0,357 | 0,16 | 0,449 |
| M, ASIA B/C/D<br>($n = 42$) | Sexual activity | 0,08 | 0,635 | -0,15 | 0,359 |
|  | Interest in sex | 0,21 | 0,189 | -0,03 | 0,870 |
|  | Sexual satisfaction | 0,03 | 0,858 | -0,01 | 0,954 |
| F, ASIA A<br>($n = 8$) | Sexual activity | -0,21 | 0,620 | -0,56 | 0,151 |
|  | Interest in sex | -0,33 | 0,429 | -0,17 | 0,690 |
|  | Sexual satisfaction | 0,33 | 0,428 | -0,12 | 0,783 |
| F, ASIA B/C/D<br>($n = 6$) | Sexual activity | -0,11 | 0,843 | -0,11 | 0,843 |
|  | Interest in sex | 0,20 | 0,707 | -0,40 | 0,437 |
|  | Sexual satisfaction | -0,94 | **0,005** | -0,72 | 0,107 |

*Adnotation.* Abbreviations: M – male, F – female.

The results of the analysis showed that sexual satisfaction decreased with age among women with spinal cord injury classified as AIS B, C, or D (a strong negative correlation, rho = -0.94, p = 0.005). Neither age nor time since injury were associated with changes in sexual activity or interest in sex, regardless of gender or the degree of impairment in motor, reflex, and sensory function (AIS classification).

#### 4.2.3 The relationship between primary symptoms of sexual dysfunction after SCI and sexual activity, sexual interest, and sexual satisfaction by gender and spinal cord injury classification

To examine the relationships between primary symptoms of sexual dysfunction and the levels of the three domains of sexual quality of life, Spearman’s rho test was used. The results are presented in Table 24.

**Table 24.**
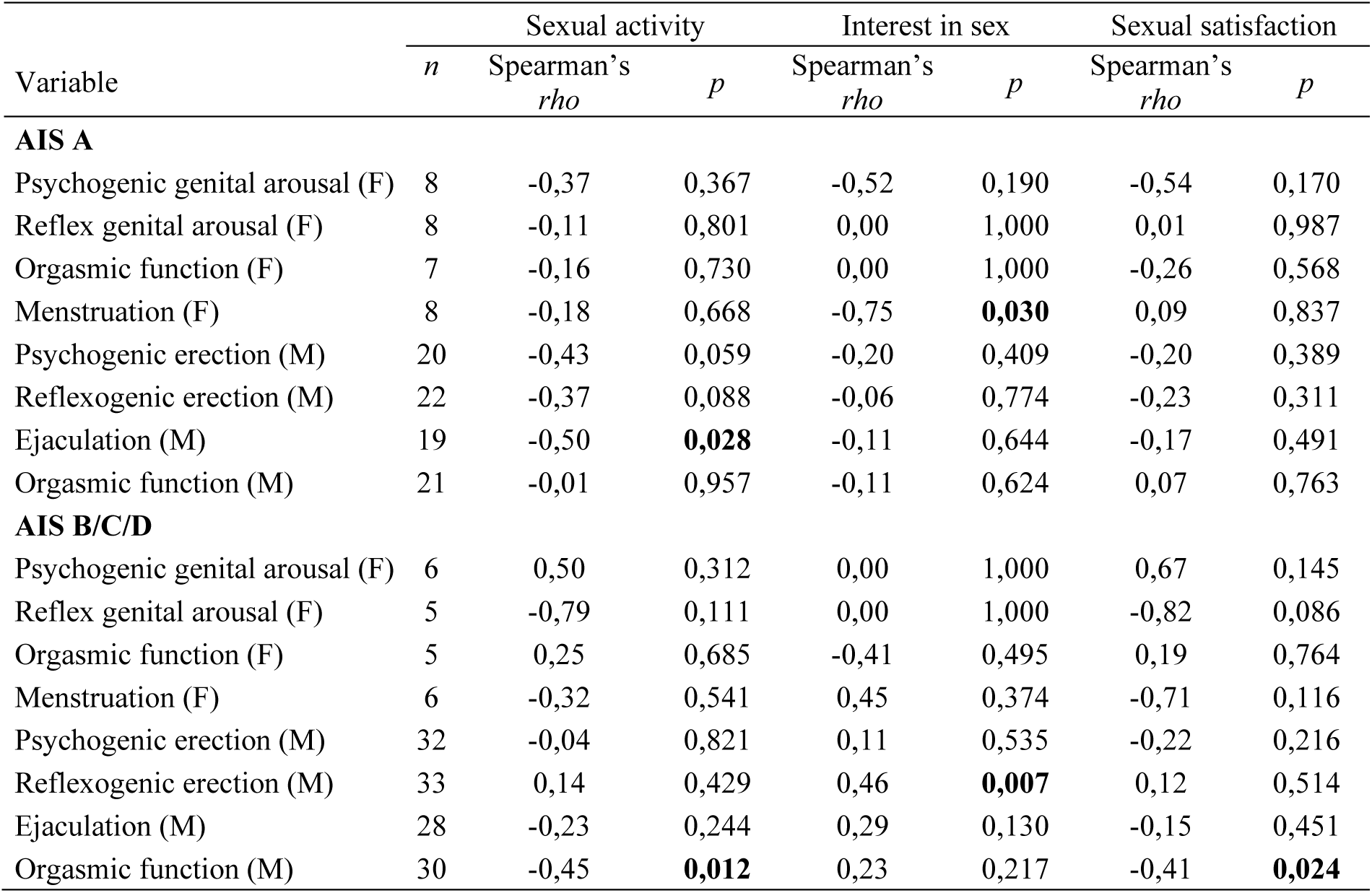
Results of Spearman’s rho correlation analysis between primary symptoms of sexual dysfunction after SCI and sexual activity, interest in sex and sexual satisfaction.

| Variable | *n* | Sexual activity |  | Interest in sex |  | Sexual satisfaction |  |
| --- | --- | --- | --- | --- | --- | --- | --- |
|  |  | Spearman's<br>*rho* | *p* | Spearman's<br>*rho* | *p* | Spearman's<br>*rho* | *p* |
| **AIS A** |  |  |  |  |  |  |  |
| Psychogenic genital arousal (F) | 8 | -0,37 | 0,367 | -0,52 | 0,190 | -0,54 | 0,170 |
| Reflex genital arousal (F) | 8 | -0,11 | 0,801 | 0,00 | 1,000 | 0,01 | 0,987 |
| Orgasmic function (F) | 7 | -0,16 | 0,730 | 0,00 | 1,000 | -0,26 | 0,568 |
| Menstruation (F) | 8 | -0,18 | 0,668 | -0,75 | **0,030** | 0,09 | 0,837 |
| Psychogenic erection (M) | 20 | -0,43 | 0,059 | -0,20 | 0,409 | -0,20 | 0,389 |
| Reflexogenic erection (M) | 22 | -0,37 | 0,088 | -0,06 | 0,774 | -0,23 | 0,311 |
| Ejaculation (M) | 19 | -0,50 | **0,028** | -0,11 | 0,644 | -0,17 | 0,491 |
| Orgasmic function (M) | 21 | -0,01 | 0,957 | -0,11 | 0,624 | 0,07 | 0,763 |
| **AIS B/C/D** |  |  |  |  |  |  |  |
| Psychogenic genital arousal (F) | 6 | 0,50 | 0,312 | 0,00 | 1,000 | 0,67 | 0,145 |
| Reflex genital arousal (F) | 5 | -0,79 | 0,111 | 0,00 | 1,000 | -0,82 | 0,086 |
| Orgasmic function (F) | 5 | 0,25 | 0,685 | -0,41 | 0,495 | 0,19 | 0,764 |
| Menstruation (F) | 6 | -0,32 | 0,541 | 0,45 | 0,374 | -0,71 | 0,116 |
| Psychogenic erection (M) | 32 | -0,04 | 0,821 | 0,11 | 0,535 | -0,22 | 0,216 |
| Reflexogenic erection (M) | 33 | 0,14 | 0,429 | 0,46 | **0,007** | 0,12 | 0,514 |
| Ejaculation (M) | 28 | -0,23 | 0,244 | 0,29 | 0,130 | -0,15 | 0,451 |
| Orgasmic function (M) | 30 | -0,45 | **0,012** | 0,23 | 0,217 | -0,41 | **0,024** |

The conducted analyses showed that the greater the severity of ejaculation problems in men (AIS A group), the lower their sexual activity. The relationship between these variables is negative and strong (rho = - 0.50, p = 0.028).

With regard to men classified as AIS B, C, or D, the collected data indicate that the more difficulties they experienced with the ability to reach orgasm, the lower their sexual satisfaction (moderate correlation, rho = -0.41, p = 0.024), and the lower their sexual activity (rho = -0.45, p = 0.012). It was also observed that the greater the problems they experienced with reflex erections, the higher their level of sexual interest (this relationship was positive and of moderate strength, rho = 0.46, p = 0.007).

In the group of women classified as AIS A, the analyses revealed a significant relationship between menstrual regularity and sexual interest. The correlation was negative and strong (rho = -0.75, p = 0.030).

#### 4.2.4 The Relationship Between Secondary Symptoms of Sexual Dysfunction After SCI and Sexual Activity, Interest, and Satisfaction, by Gender and Classification of Spinal Cord Injury

In order to determine the relationships between secondary symptoms of sexual dysfunction after SCI and the domains of sexual quality of life, Spearman’s rho test was used. The results are presented in Table 25.

**Table 25.**
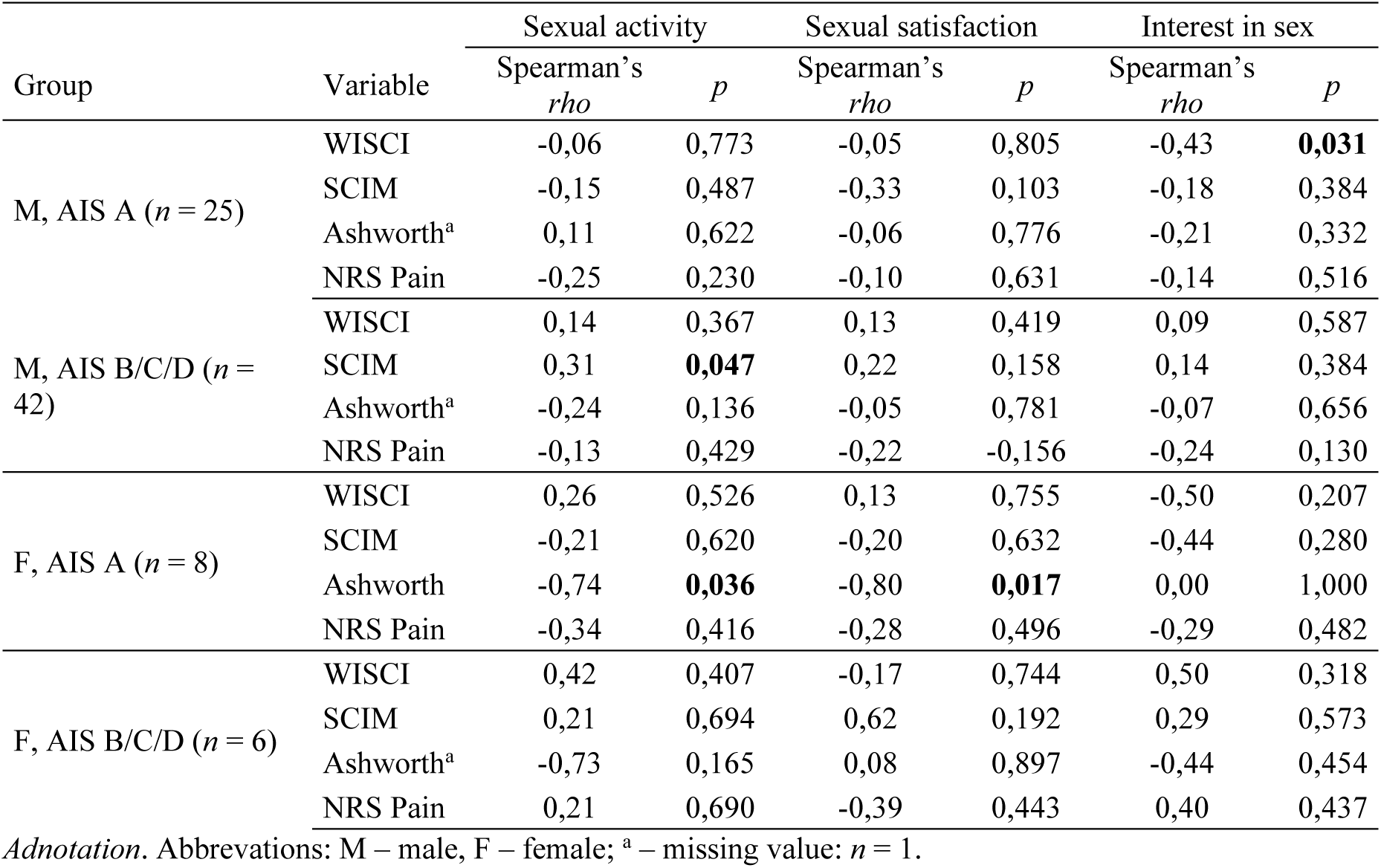
Results of Spearman’s rho correlation analysis between secondary symptoms of sexual dysfunction after SCI and sexual activity, interest in sex and satisfaction.

The data obtained showed that the higher the functional ability (SCIM-III) among men with an AIS B, C, or D classification, the smaller the decline in their sexual activity after SCI (moderate correlation, rho = 0.31, p = 0.047).

In the group of men with an AIS A classification, it was found that the greater their independence in the area of mobility and walking (WISCI II), the smaller the changes in sexual interest (moderate correlation, rho = - 0.43, p = 0.031).

In the group of women with an AIS A classification, it was found that the higher the level of spasticity, the lower their sexual activity and satisfaction after SCI (strong correlations: rho = -0.74, p = 0.036 for sexual activity, and rho = -0.80, p = 0.017 for sexual satisfaction).

The remaining relationships between the examined variables were not statistically significant.

#### 4.2.5. The relationship between anxiety, depression, and sexual attractiveness and the quality of sexual life depending on gender and spinal cord injury classification

To address the research question concerning the relationship between tertiary symptoms related to sexual problems after SCI and the domains of sexual quality of life, the Spearman’s rho test was used. The results are presented in Table 26.

**Table 26.** Spearman’s rho correlations between psychosocial symptoms and changes in sexual activity and sexual satisfaction by gender and SCI classification.

| Group | Variable | Sexual activity |  | Sexual satisfaction |  | Interest in sex |  |
| --- | --- | --- | --- | --- | --- | --- | --- |
|  |  | Spearman's<br>*rho* | *p* | Spearman's<br>*rho* | *p* | Spearman's<br>*rho* | *p* |
| M, AIS A<br>(*n* = 25) | STAI | -0,06 | 0,759 | 0,02 | 0,939 | 0,24 | 0,243 |
|  | KPD | -0,03 | 0,888 | 0,11 | 0,601 | -0,08 | 0,710 |
|  | Sexual<br>attractiveness | 0,68 | **<0,001** | 0,70 | **<0,001** | 0,26 | 0,214 |
| M, AIS<br>B/C/D<br>(*n* = 42) | STAI | -0,03 | 0,839 | -0,13 | 0,425 | -0,38 | **0,012** |
|  | KPD | -0,20 | 0,195 | -0,37 | **0,017** | -0,47 | **0,002** |
|  | Sexual<br>attractiveness | 0,75 | **<0,001** | 0,82 | **<0,001** | 0,48 | **0,001** |
| K, AIS A<br>(*n* = 8) | STAI | -0,79 | **0,021** | -0,65 | 0,081 | -0,33 | 0,423 |
|  | KPD | -0,53 | 0,172 | -0,09 | 0,835 | 0,11 | 0,797 |
|  | Sexual<br>attractiveness | 0,37 | 0,367 | 0,18 | 0,672 | 0,00 | 1,000 |
| K, AIS<br>B/C/D<br>(*n* = 6) | STAI | -0,84 | **0,036** | 0,00 | 1,000 | -0,30 | 0,568 |
|  | KPD | -0,62 | 0,188 | 0,49 | 0,320 | -0,29 | 0,573 |
|  | Sexual<br>attractiveness | 0,50 | 0,312 | 0,67 | 0,145 | 0,00 | 1,000 |

*Adnotation.* Abbreviations: M – male, F – female.

The conducted analyses showed that an increase in anxiety among women with AIS A and AIS B, C, D spinal cord injury classifications was associated with decreased sexual activity (a strong negative correlation: rho = -0.79, *p* = 0.021 and rho = -0.84, *p* = 0.036, respectively). Moreover, in the group of women with AIS A injuries, higher levels of anxiety were linked to lower perceived sexual satisfaction (a strong negative correlation, *p* = 0.081). No such associations were observed in the group of men.

In the group of men with spinal cord injuries classified as AIS B, C, or D, it was found that higher levels of depression were associated with lower perceived sexual satisfaction (a moderate negative correlation, rho = -0.37, *p* = 0.017). Furthermore, it was shown that higher levels of both anxiety and depression were linked to lower levels of sexual interest (moderate negative correlations: for anxiety, rho = -0.38, *p* = 0.012; for depression, rho = -0.47, *p* = 0.002). Among all male participants, a strong positive correlation was observed between perceived sexual attractiveness and each of the three domains of sexual quality of life: sexual activity (rho = 0.75), sexual satisfaction (rho = 0.82), and sexual interest (rho = 0.48), with *p* ≤ 0.001 for all three domains.

#### 4.2.6. Differences in the levels of sexual quality of life domains between individuals with spinal cord injury and individuals from the general population (without SCI)

To address the research question concerning the differences in the general domains of sexual quality of life between individuals with SCI and those from the general population without spinal cord injury, descriptive statistics were first calculated along with a normality test for age, time since injury, and sexual quality of life domains, with results stratified by gender. The Shapiro-Wilk test was used for this purpose. The results are presented in Tables 27–29.

**Table 27.**
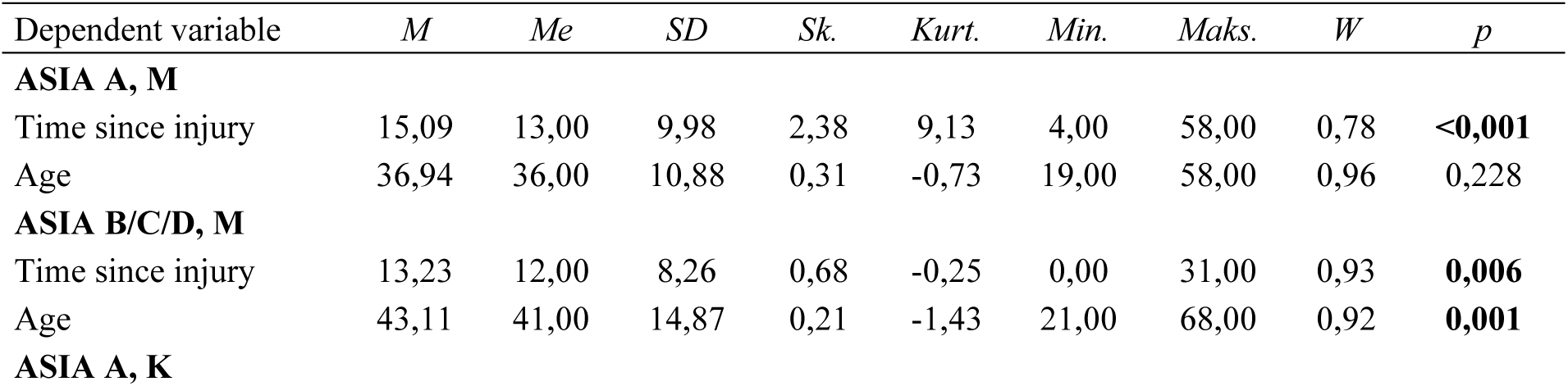
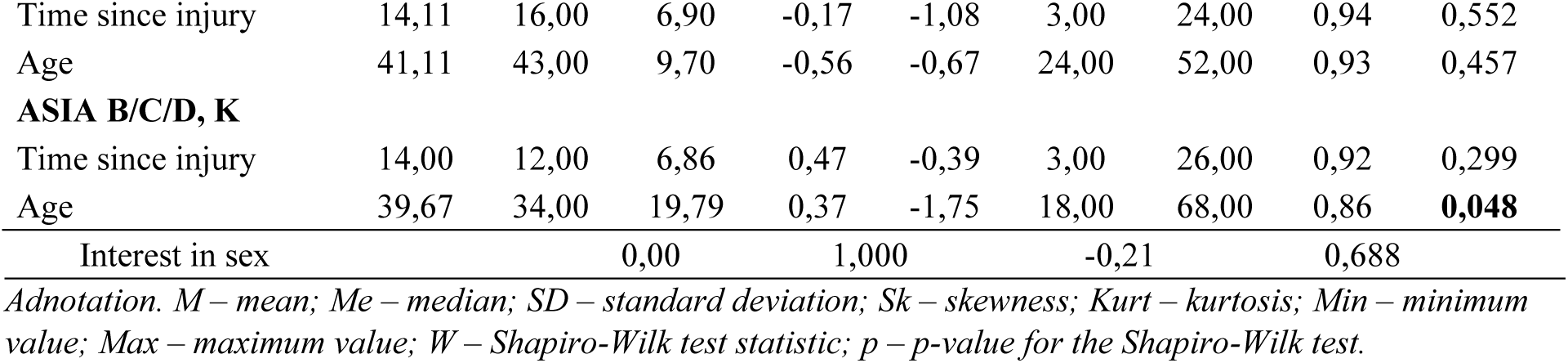
Descriptive statistics and Shapiro–Wilk test results for age and time since injury by gender and level of disability.

**Table 28.** Descriptive statistics and Shapiro–Wilk test results for domains of sexual quality of life by gender and presence of disability.

| Dependent variable | *M* | *Me* | *SD* | *Sk.* | *Kurt.* | *Min.* | *Maks.* | *W* | *p* |
| --- | --- | --- | --- | --- | --- | --- | --- | --- | --- |
| **Women** |  |  |  |  |  |  |  |  |  |
| Sexual activity | 2,51 | 2,00 | 1,05 | 0,28 | -0,54 | 1,00 | 5,00 | 0,90 | **<0,001** |
| Sexual interest | 3,11 | 3,00 | 0,92 | 0,26 | -0,21 | 1,00 | 5,00 | 0,89 | **<0,001** |
| Sexual satisfaction | 2,79 | 3,00 | 1,19 | 0,09 | -0,89 | 1,00 | 5,00 | 0,91 | **<0,001** |
| **Men** |  |  |  |  |  |  |  |  |  |
| Sexual activity | 3,00 | 3,00 | 1,13 | -0,06 | -0,53 | 1,00 | 5,00 | 0,91 | **<0,001** |
| Sexual interest | 3,26 | 3,00 | 1,15 | -0,17 | -0,70 | 1,00 | 5,00 | 0,91 | **<0,001** |
| Sexual satisfaction | 3,40 | 3,00 | 1,04 | -0,08 | -0,57 | 1,00 | 5,00 | 0,90 | **<0,001** |
| **Individuals with disabilities (ASIA A, B, C, D)** |  |  |  |  |  |  |  |  |  |
| Sexual activity | 2,15 | 2,00 | 0,73 | -0,23 | -1,06 | 1,00 | 3,00 | 0,80 | **<0,001** |
| Sexual interest | 2,74 | 3,00 | 0,69 | 0,14 | 1,00 | 1,00 | 5,00 | 0,81 | **<0,001** |
| Sexual interest | 2,73 | 3,00 | 0,65 | -0,22 | 0,12 | 1,00 | 4,00 | 0,80 | **<0,001** |
| Sexual satisfaction | 2,58 | 3,00 | 1,11 | 0,22 | -0,76 | 1,00 | 5,00 | 0,91 | **<0,001** |
| **Healthy individuals** |  |  |  |  |  |  |  |  |  |
| Sexual activity | 3,22 | 3,00 | 1,14 | -0,45 | -0,41 | 1,00 | 5,00 | 0,89 | **<0,001** |
| Sexual interest | 3,56 | 4,00 | 1,11 | -0,59 | -0,29 | 1,00 | 5,00 | 0,89 | **<0,001** |
| Sexual satisfaction | 3,46 | 3,00 | 1,07 | -0,27 | -0,42 | 1,00 | 5,00 | 0,90 | **<0,001** |

**Table 29.** Results of the two-way MANOVA for domains of sexual quality of life by gender and between able-bodied individuals and individuals with disabilities (SCI).

| Gender | with SCI and without SCI | *n* | Sexual activity |  | Sexual interest |  | Sexual satisfaction |  |
| --- | --- | --- | --- | --- | --- | --- | --- | --- |
|  |  |  | *M* | *SD* | *M* | *SD* | *M* | *SD* |
| Total | with SCI | 81 | 2,15 | 0,73 | 2,73 | 0,65 | 2,58 | 1,11 |
|  | without SCI | 90 | 3,22 | 1,14 | 3,56 | 1,11 | 3,46 | 1,07 |

| Main effect<br>SCI | | | $F(1, 167) = 33,40;$<br>$p < 0,001;$<br>$\eta^2_p = 0,17$ | | $F(1, 167) = 26,71;$<br>$p < 0,001;$<br>$\eta^2_p = 0,14$ | | $F(1, 167) = 12,27;$<br>$p < 0,001;$<br>$\eta^2_p = 0,07$ | |
| --- | --- | --- | --- | --- | --- | --- | --- | --- |
| Mem | Total | 101 | 2,51 | 1,05 | 3,10 | 0,90 | 2,79 | 1,19 |
| Women |  | 70 | 3,00 | 1,13 | 3,26 | 1,15 | 3,40 | 1,04 |
| Main effect<br>gender | | | $F(1, 167) = 0,02;$<br>$p = 0,886;$<br>$\eta^2_p < 0,01$ | | $F(1, 167) = 1,50;$<br>$p = 0,223;$<br>$\eta^2_p = 0,01$ | | $F(1, 167) = 2,48;$<br>$p = 0,117;$<br>$\eta^2_p = 0,02$ | |
| Mężczyźni | With SCI | 67 | 2,12 | 0,75 | 2,72 | 0,67 | 2,49 | 1,15 |
|  | without SCI | 34 | 3,29 | 1,12 | 3,85 | 0,82 | 3,38 | 1,07 |
| Kobiety | With SCI | 14 | 2,29 | 0,61 | 2,79 | 0,58 | 3,00 | 0,78 |
|  | without SCI | 56 | 3,18 | 1,16 | 3,38 | 1,23 | 3,50 | 1,08 |
| Interaction effect | | | $F(1, 167) = 0,63;$<br>$p = 0,428;$<br>$\eta^2_p < 0,01;$ | | $F(1, 167) = 2,69;$<br>$p = 0,103;$<br>$\eta^2_p = 0,02$ | | $F(1, 167) = 1,00;$<br>$p = 0,327;$<br>$\eta^2_p = 0,01$ | |

For the variables of age and time since injury, the results in some cases indicated distributions resembling a normal distribution (p > 0.05); however, in other cases, the results were statistically significant, and in the group of men with ASIA A-level disabilities, the distribution of time since injury showed considerable skewness. The analyses aimed to examine relationships involving ordinal variables, which justified the use of a non-parametric test – Spearman’s rho.

In the case of the domains of sexual quality of life, despite significant results of the Shapiro–Wilk test, skewness and kurtosis did not exceed |2| in any instance. Therefore, it was assumed that the distributions of sexual quality of life, when divided by gender and disability status, were approximately normal [127]. For this reason, a parametric test – a two-way MANOVA – was used to examine the effects of gender and health status on sexual quality of life.

Next, an analysis was conducted to examine differences in the domains of sexual quality of life between individuals with SCI and able-bodied individuals (i.e., without SCI), taking gender into account. For this purpose, a two-way MANOVA was used. Prior to analysis, the variable *sexual interest* was transformed using the winsorizing method in the group of individuals with disabilities (where values of 5 were adjusted to 4). The results are presented in Table 19.

The analysis results showed that the main effect of sex was not statistically significant for sexual activity, sexual interest, or sexual satisfaction. This indicates that sex does not differentiate the quality of sexual life.

The main effect of disability was found to be significant for sexual activity: F(1, 167) = 33.40; p < 0.001; η²ₚ = 0.17; for sexual interest: F(1, 167) = 26.71; p < 0.001; η²ₚ = 0.14; and for sexual satisfaction: F(1, 167) = 12.27; p < 0.001; η²ₚ = 0.07. According to the partial eta-squared values, the strongest effect was observed for sexual activity, followed by sexual interest (both strong effects), and the weakest for sexual satisfaction (a moderate effect). The analysis of means showed that the quality of sexual life was higher among individuals without disabilities compared to those with disabilities. The interaction effect of sex and disability was not statistically significant for any of the domains of sexual quality of life-sexual activity, sexual interest, or sexual satisfaction.

## 5 Discussion

### 5.1. Three Levels of Sexual Dysfunction in the Study Population

#### 5.1.1. Primary and Secondary Levels of Sexual Dysfunction Following Spinal Cord Injury in the Study Group

The data obtained indicate that 87.7% of the men surveyed (i.e., 87.5% of those with spinal cord injuries classified as AIS A and 87.8% as AIS B, C, or D) and 83.4% of women (77.8% in the AIS A group and 88.9% in the AIS B, C, or D group) reported experiencing sexual problems related to spinal cord injury. The present findings support estimates from other researchers suggesting that up to 88% of women with SCI experience at least one type of sexual dysfunction, compared to 37% of women in the general population [65]. Studies conducted on male populations similarly indicate that the majority face such difficulties [17,19,65]. One of the most recent studies estimated that virtually all participants reported adverse changes in their overall sexual functioning, including physiological, psychological, and relational aspects [8].

#### 5.2.3 Primary and Secondary Levels of Sexual Problems Related to Spinal Cord Injury

The study described in this work demonstrated that SCI impairs the ability to achieve both psychogenic and reflex arousal of the genital organs in both women and men.

Regarding psychogenic arousal, the greatest difficulties were reported by participants with injuries classified as AIS B, C, or D-namely, 77.8 % of women and 65.8 % of men. All women in this group described their psychogenic arousal as reduced or altered; similarly, 46.3 % of men rated their psychogenic erection as reduced or altered, and 19.5 % reported it as absent. Marked problems were also reported by men with AIS A injuries (65.6 %). By contrast, most women in the AIS A group retained the ability to achieve psychogenic genital arousal. When examined by neurological level, the severest psychogenic-arousal problems occurred in participants-both women and men-with lumbar-level injuries. Among these, 83.3 % of women described their psychogenic arousal as reduced or altered. 77.78 % of men in this subgroup reported difficulties, with 61.11 % rating their psychogenic erection as reduced or altered and 16.67 % as absent. Most women and men-regardless of AIS classification-also experienced difficulties with reflex genital arousal. The highest prevalence was again observed in the AIS B, C, D cohort: 44.4 % of women and 58.5 % of men noted diminished reflex arousal, while 22.2 % and 19.5 %, respectively, reported complete loss of this function. These findings align with earlier studies showing that approximately 67.7 % to 80 % of men with SCI experience erectile difficulties [8, 19]. Robust statistics for both forms of sexual arousal-particularly in women-remain scarce. One available dataset reports that 23.2 % of women with SCI experience insufficient vaginal lubrication, causing discomfort or pain during intercourse [132], a figure comparable to that found in our analysis.

Ejaculatory dysfunction is frequently observed among individuals with SCI. The majority of participants in the current study reported difficulties related to ejaculation. As many as 81% of men with complete SCI experienced ejaculatory problems, with 62.5% of them reporting a complete absence of ejaculation. Men with injuries classified as AIS B, C, or D also reported substantial difficulties: 17% described their ejaculation as reduced, while 48.8% stated it was absent. These findings are consistent with those of previous studies. Miranda et al. found that 89.4% of male participants reported issues with ejaculation. Similarly, Yarkony’s research showed that only 10% of individuals with complete SCI and 32% with incomplete SCI retained the ability to ejaculate [19]. Comparable results were presented by Chehensse et al. in a review and meta-analysis, where 11.8% of men with complete SCI and 33.2% of those with incomplete SCI retained ejaculatory function [133,134].

The present study did not find a correlation between the level of spinal cord injury and the ability to ejaculate. These results are inconsistent with previous findings suggesting that damage to the sympathetic center at the T11– T12 level affects the initiation and progression of the first phase of ejaculation, while lesions at the S2–S4 level are associated with disturbances in the second phase [15]. A potential explanation for this discrepancy lies in the methodology of the current study, in which participants were grouped solely based on the level of injury, without taking into account the extent of the lesion. This grouping approach was adopted due to the significant imbalance and small sample sizes within subgroups when using a more detailed classification.

The results of the study indicated that the majority of participants experienced difficulties or a complete inability to reach orgasm, namely 72.2% of women (including 77.7% in the AIS A group and 66.7% in the AIS B, C, D group) and 77% of men (respectively 75.1% and 78%). The greatest difficulties in this area were reported by women with incomplete spinal cord injuries, 55.6% of whom reported a complete loss of the ability to achieve orgasm. Among men, those with complete spinal cord injuries more frequently and more intensely reported orgasmic dysfunction, with 56.3% indicating the complete inability to reach orgasm. Taking into account the level of spinal cord injury, the most severe orgasm-related problems were observed in both men and women with thoracic and lumbar spinal cord injuries. Among men with thoracic-level injuries, 24.2% reported reduced or altered orgasmic function, and 60.6% reported a complete loss of this function. In the lumbar injury group, 41.2% of men reported a reduced or altered orgasmic response, while 50% indicated a complete absence of orgasm. These findings confirm that both the extent and location of the spinal cord injury play a crucial role in post-SCI sexual functioning.

The obtained results are consistent with the findings of other researchers. Some studies report that approximately 50% of individuals with SCI experience difficulties in achieving orgasm [11,27]. Other studies indicate even higher percentages, such as Di Bell et al. – 74.5%, and Sramkova et al. – 73.4% [15,25].

The results concerning orgasm are inconclusive. This may be due to several factors, such as difficulties experienced by respondents in identifying and describing the sensation and experience of orgasm, possible distortions of results caused by medications taken, and general health status. Moreover, the time required to reach orgasm is significantly prolonged in individuals with SCI, which may result in engaging in sexual intercourse without achieving full sexual satisfaction.

The majority of women participating in the study reported having a regular menstrual cycle. These findings are consistent with those of other studies [29–30].

The present study examined the relationships between time since injury, age, and primary sexual dysfunctions following SCI. The analysis showed that the longer the time since injury, the more dysfunctions occurred in the area of reflex sexual arousal of the female genital organs. The same association was found for age-i.e. the older the women, the more dysfunctions were reported in this domain (the average age of the female participants was 36). This decline may be partially explained by natural ageing processes. Other studies have demonstrated a link between age and a decline in female sexual functioning beginning between the ages of 20 and 30, and intensifying around menopause [135–137]. However, attributing this decline solely to ageing processes may be insufficient. It is important to emphasize that female sexual dysfunctions following SCI are more complex and less well-studied than those in men. They cannot be considered solely in terms of neurological, vascular, or hormonal mechanisms, without taking into account psychological, interpersonal, and social factors [138].

The collected data indicate that, among men, increasing age is associated with a growing difficulty in achieving reflex erection. The decline in the ability to achieve reflex erection may be explained by the general decrease in sexual functioning observed with age, which is also seen in the general population. Furthermore, among men with injuries classified as AIS B/C/D, the longer the time since the injury, the fewer dysfunctions were reported regarding the ability to achieve psychogenic erection and its quality. Some authors suggest that in approximately 60% of patients with complete and incomplete SCI, erectile function improves within 6 months, in 80% within a year, and in 5% after two years post-injury [139]. This phenomenon can be partially explained by recovery and regeneration following spinal shock, as well as by spontaneous mechanisms of neuroplasticity and remyelination [140].

All study participants experienced neurological consequences of spinal cord injury. In terms of functional independence in self-care, respiration, sphincter control, and mobility, they achieved comparable results regardless of the analyzed group. The best functioning was observed among men with incomplete spinal cord injuries (AIS B, C, D). Functional assessment of walking and mobility independence showed that both women and men with incomplete injuries (AIS B, C, D) were significantly more physically capable and independent compared to individuals with complete injuries (AIS A).

In terms of perceived pain within the studied population, the average level was moderate. Among women with an injury classified as AIS A, the mean score was 3.44, while for those classified as AIS B, C, or D, it was 3.25. Similarly, in the male group, the average pain level was 3.49 for AIS A and 3.02 for AIS B/C/D (on a scale of 0 to 10). It is worth noting that while the range between the lowest and highest scores among women was between 1 and 7, this range was wider among men: 1–9 in the group with incomplete injury and 1–10 in the group with complete spinal cord injury. The results obtained in this study are lower than those reported in other research. For example, in a cohort study of individuals after SCI conducted by Müller et al., the mean pain score was 6 (±2) points. That study involved individuals with chronic spinal cord injury [141]. Other findings (concerning neuropathic pain after SCI) were similar, with an average of 4.52 points (4.90 in the female sample and 4.37 among men) [142].

In the conducted study, the secondary level of sexual dysfunction (SD) included issues related to cardiological disorders and sphincter function. The participants did not present with metabolic system disorders associated with SCI. Data related to endocrine system disorders and the impact of medication on sexual functioning were not analyzed.

#### 5.1.2. The relationship between psychosocial (Tertiary) symptoms after SCI and their potential impact on sexuality

The majority of participants did not score above the clinical threshold for anxiety or depression. However, 6.2% exceeded the cutoff point for a clinical diagnosis of depression. This value is similar to the results found in the general European population [143]. It is worth noting that the result obtained in this study is lower than those reported by other researchers, who indicated rates ranging between 19–26%. Other studies show that 20.9% of people with SCI experienced mild depression, 17.9% moderate, and 10.4% severe depression [144–145]. The relatively low rate observed in this study may be explained by the specific characteristics of the sample. Participants enrolled voluntarily, which suggests that they were actively seeking help and were motivated to undertake long-term, demanding institutional rehabilitation. Most participants were in a committed relationship at the time of the study (i.e., 58% of men and 52.4% of women). It is also worth mentioning that research conducted among people with spinal cord injuries has shown that individuals in stable relationships report higher levels of sexual satisfaction [14].

The average level of subjective sexual attractiveness among the participants ranged from 2.2 to 2.67 points on a 5-point scale and showed no significant variation. Men rated their own attractiveness slightly lower. A study using a semi-structured interview revealed that individuals with spinal cord injury often struggle with reduced self-confidence in the sexual domain, which affects both sexual activity and satisfaction [84]. Other studies have shown that a partner’s satisfaction, relationship quality, mood, and sense and degree of independence were more important predictors of sexual satisfaction than the mere preservation of genital function among both women and men with SCI [14].

### 5.2. The relationship between factors influencing sexual problems after SCI and *patients’ sexual interest, activity, and satisfaction*

In the present study, the domains of general sexual quality of life were defined as: subjective level of sexual interest, sexual activity, and sexual satisfaction. The sexual activity of the participants was positively correlated with sexual satisfaction-that is, the higher their sexual activity, the greater their satisfaction from these acts. These results are consistent with those obtained by other researchers [89]. The average level of sexual interest among the studied men was 2.72 (AIS A) and 2.74 (AIS B, C, D), and among women, 3.00 (AIS A) and 2.50 (AIS B, C, D), on a self-reported scale from 1 to 5. The results indicate relatively low sexual interest among the participants. Sexual interest after SCI differed significantly from that declared by able-bodied individuals (i.e., without SCI), which is not consistent with findings from other researchers. A. Reitz et al. showed that 53% of respondents rated their sexual interest after SCI as very high or high-higher than before the injury-and 44% of respondents reported a similar level [97].

The average sexual activity of the studied men (assessed on a self-reported scale from 1 to 5) was 2.12 (regardless of AIS classification). In the population of individuals without SCI, the corresponding value was 3.22, and this difference was statistically significant. Data collected during interviews suggest that the most common reason for this decline is the lack of opportunities for intimate contact due to the long-term rehabilitation process carried out in centers without the presence of a partner and the lack of adequate privacy for patients. The data indicating low activity among individuals with SCI are consistent with the findings of other researchers (including Otero-Vilaverde et al., Gomes et al., Dahlberg et al., Reitz et al.) [79–80, 94, 97].

In terms of sexual satisfaction, the average value, estimated on a five-point scale, was 2.20 (AIS A) and 2.67 (AIS B, C, D) in the group of men, and 2.88 (AIS A) and 3.17 (AIS B, C, D) in the group of women. These results are lower than those obtained in the general population, as confirmed by other analyses. The study by Gomes C.M. et al. (2017) indicated that the percentage of men with SCI who were satisfied with their sex life was 21.3%, whereas in the general male population this figure was as high as 94.2% [80]. Among women, the respective percentages were 51% and 62% [96]. A multivariate analysis by Sale et al. (2012) showed that 31% of individuals with SCI (both women and men) were satisfied with their sexual activity [100].

Time since injury was not associated with the factors of overall sexual quality of life. However, other researchers have shown that with time and adjustment to the disability, sexual satisfaction among individuals tends to increase again [73,146]. All participants in the described study were at an early stage post-SCI (i.e. within two years of the injury) and shared a similar experience of prolonged hospitalization followed by intensive, long-term rehabilitation outside their place of residence, under conditions that limited intimacy and closeness with a partner.

In the group of male participants, no relationship was observed between age and the overall domains of sexual quality of life. Among women with an incomplete spinal cord injury, a relationship was observed between age and sexual satisfaction, suggesting that the older the women were, the lower their satisfaction in the intimate sphere. This result differs from findings reported by other researchers [79]. The References emphasizes that women may maintain sexual satisfaction after SCI, but this depends on many factors, such as: the possibility of intimacy and privacy, access to sexual education, the degree of tolerance and openness, avoidance of stigmatization, the quality and stability of the partner relationship, general mental health, self-esteem, etc. [14,16,93]. The majority of women participating in the study were in a stable relationship (52.4%), but the quality of the partner relationship was not analyzed. The group of women did not differ significantly from the group of men in terms of psychological and somatic problems. Most of the women (81%) had a higher education. Possible explanations for the above results include: the small size of the female sample (N=21) and generational differences. The average age of the women in the study was 41 years, and the group was not generationally homogeneous. Access to sexual education and attitudes toward sexuality have changed in Poland over the past 30 years, which may have influenced the readiness for adaptation and experimentation among older women in the sample. However, these aspects require further empirical investigation.

#### 5.2.1. Dynamics of sexual quality of life domains in the studied population

The results obtained indicated that sexual activity among individuals with SCI is positively associated with sexual satisfaction in all male participants, as well as among women with complete spinal cord injury. However, the study did not reveal any relationship between sexual interest and levels of sexual activity or satisfaction. This may be due to the varied ways in which “sexual interest” is understood and the different factors underlying it. High sexual desire may result from unmet sexual needs, may be a personality trait, or may relate to situational context. These factors can influence sexual activity and the level of sexual interest-either high or low- and, depending on the adequacy of one’s sexual activity, may or may not be experienced as satisfying. It is worth emphasizing that the study participants were in the early stage following SCI, which, on the one hand, focused their attention on somatic health and physical autonomy. On the other hand, some participants reported experiencing considerable sexual frustration due to the deprivation of this need. Sexual interest was generally low and showed no correlation with the low levels of sexual activity and satisfaction. This could be attributed to the redirection of attention and engagement toward other areas of daily functioning and more immediate needs. In some cases, although sexual interest was present, it was not associated with sexual activity, as fantasies and desires could not be fulfilled-an issue which may have, in turn, negatively influenced sexual satisfaction. All domains of sexual quality of life were rated low by the participants. Institutionalized rehabilitation-prolonged, carried out away from home, and without the presence of a partner-had an impact on sexual activity and the process of adjusting to a new sexual reality after injury, which likely contributed to the low levels of sexual satisfaction reported by the participants.

In the described study, quantitative data were not collected for the purpose of analyzing the quality of romantic relationships, beliefs about male and female roles, or the sexual potential of individuals with disabilities. These beliefs were considered individually and were treated as qualitative data.

#### 5.2.2. Primary and secondary levels of sexual dysfunction following SCI and their impact on sexual interest, activity, and satisfaction

The results of the analyses suggest that primary and secondary problems following SCI affect the overall quality of sexual life.

Reflex erection problems were associated with higher sexual interest among men with incomplete spinal cord injury. The data collected support patients’ narratives of experiencing sexual frustration due to the inability or limited ability to engage in sexual intercourse caused by insufficient or absent erection. However, it remains unclear why this issue only applies to men with incomplete spinal cord injuries and only to reflex erection. No such correlation was observed in individuals with injuries classified as AIS A, nor with psychogenic erection. The data analysis also revealed that the greater the orgasm-related dysfunction reported by patients, the lower their sexual activity and satisfaction. Ejaculatory problems (altered/reduced or absent ejaculation) were associated with lower sexual activity as well. These correlations were also found only among patients classified as AIS B, C, or D. These results may be partly explained by the disparity in group sizes (N AIS A = 35 vs. N AIS B, C, D = 54).

Among women, problems with reflex genital arousal were associated with lower sexual satisfaction, although this correlation appeared only at a trend level and only among women with incomplete spinal cord injury. The study also indicated that, in the group of women classified as ASIA A, greater severity of menstruation-related problems was linked to decreased sexual interest.

Significant correlations were also observed in the area of secondary symptoms following SCI. Among men with incomplete spinal cord injury, higher levels of functional independence (in terms of mobility and self-care in daily functioning) were associated with greater sexual activity. This may suggest that increased physical self-sufficiency enables patients to engage more frequently in sexual contact or behavior. The results also indicated that maintaining better physical function and independence in walking and mobility was linked to increased sexual interest among men with complete spinal cord injury.

Among women, it was observed that greater problems with muscle spasticity were associated with lower levels of sexual activity and satisfaction. Issues related to excessive muscle tone may be linked to pain and, more importantly, to difficulties in achieving appropriate body positioning during sexual intercourse. Experiencing such difficulties may lead to decreased sexual activity and prevent women from fully enjoying the experience.

Based on the above data, it can be concluded that primary and secondary issues following SCI affect the overall quality of sexual life; however, this impact is neither straightforward nor unidirectional. It can be assumed that these problems may serve as predictors of reduced sexual health, but which variable-and to what extent-it influences an individual’s sexual interest, activity, or satisfaction will depend on the interplay of various factors and personal predispositions.

#### 5.2.3. Psychosocial factors and their relationship with sexual interest, activity, and satisfaction

Among the psychosocial factors influencing sexual problems after SCI, the study included low mood, depression, anxiety, and the subjective perception of sexual attractiveness. The obtained results indicated that higher levels of anxiety were associated with decreased sexual activity in women with incomplete spinal cord injury. This relationship was not observed among women classified as AIS A. In the group of men with complete spinal cord injury, anxiety, low mood, and depression negatively impacted both sexual interest and satisfaction. These associations were not found in the group of men with incomplete injuries. These findings are consistent with those of other researchers concerning the link between depression, anxiety, and sexuality in individuals with SCI [147,14]. The absence of such associations in certain subgroups may be partly explained by the generally low levels of psychological distress observed. Moreover, the studied groups might have differed in other factors influencing the identified domains of sexual quality of life, such as relationship quality, partner’s sexual satisfaction, or stereotypes about sexuality-which were not covered in this study [14,16,27].

The level of perceived sexual attractiveness also plays an important role in sexual interest, activity, and satisfaction - but only among men. A higher self-assessment of sexual attractiveness was associated with increased sexual activity and satisfaction in all men, regardless of the classification of spinal cord injury. Moreover, a higher level of perceived attractiveness was linked to greater sexual interest, but only in men with incomplete spinal cord injury. These findings are consistent with those of other researchers who emphasize the influence of perceived sexual attractiveness, as well as self-esteem and a sense of agency, on sexual functioning after spinal cord injury [14,16,81].

The psychosocial factors analysed in the study were associated with the domains of overall sexual quality of life in the participants; however, as with primary and secondary symptoms, their impact on sexual quality of life was not straightforward.

### 5.3. Implications

The aforementioned primary, secondary, and tertiary dysfunctions following SCI that affect patients’ sexuality determine the scope, direction, and objectives of therapeutic and medical interventions. Human sexuality is multifactorial (biopsychosocial model); therefore, it requires an interdisciplinary, coordinated approach which, when introduced as early as possible, may prevent the consolidation of medical problems and the accumulation of psychological difficulties (e.g., development or reinforcement of maladaptive beliefs, lowered self-esteem, deterioration of intimate relationships, withdrawal from partnerships, etc.). The conducted study indicates that 56.6% of respondents had never received any counselling regarding sexual life and had not been provided with any medical or psychological support in this area, despite the vast majority expressing interest in such support. Patients rarely receive targeted assistance in the area of sexual functioning and, if they do, it usually occurs at a relatively late stage after SCI. This situation may be explained by insufficient training of healthcare and support personnel to address sexuality-related issues, embarrassment or fear of discussing sexual topics (as sex remains a taboo subject for many), and prejudices that invalidate the sexual needs of people with disabilities. Organisational challenges in medical facilities-such as insufficient staffing or lack of time-also play a significant role [83, 148].

Based on the data described above, a proprietary therapeutic intervention protocol was developed, tailored to the needs of patients with SCI in the area of sexual health. In light of the positive evaluations provided by SCI patients who participated in treatment and rehabilitation activities carried out according to this protocol, it is advisable to recommend its implementation within comprehensive rehabilitation programs dedicated to this patient group (Annex 3).

## Limitations

Although the research project aimed to address the issue of sexuality among individuals with spinal cord injury (SCI) as comprehensively as possible, it did not include quantitative data on several important aspects: the assessment of relationship quality, beliefs regarding male and female roles, the potential of people with disabilities, the main premises regarding sexual activity among individuals with disabilities, and the values they uphold. These issues are of considerable interest and should be taken into account in future analyses.

Another limitation was the research procedure itself. During the study, the psychologist was not a blinded member of the research team and read the questionnaire items to participants, marking the answers on their behalf due to the participants’ physical disabilities. This could have affected the quality of the collected data, particularly due to the influence of social desirability bias. In future studies, it would be advisable to support the psychologist’s work with information technology and modern computer software to enable participants to complete the questionnaires independently. It would also be beneficial if the assessor were unaware of the purpose of the research program.

This study was part of a scientific research project aimed at assessing the effectiveness of Robot-Assisted Gait Training (RAGT) and comparing it with conventional rehabilitation methods using a Dynamic Parapodium (DPT). The initial hypotheses formulated in this study concerned the relationship between the rehabilitation modality (RAGT vs DPT) and changes across three levels of sexual dysfunction, as well as changes within three domains of sexual quality of life. However, the study did not account for the possibility that patients in the control group would access robotic rehabilitation in other rehabilitation centers. Although the comparative analysis of sexual functioning in individuals with spinal cord injury undergoing treatment with Lokomat and/or exoskeletons is an interesting research area, this goal could not be achieved. For ethical reasons, it was not possible to require patients to refrain from attempting RAGT-based therapy during the course of the study.

## Author Contributions

Conceptualization, A.W.-S. and B.T.; methodology, A.W.-S., B.T. and B.K.; software, A.W.-S.; formal analysis, A.W.-S.; investigation, A.W.-S., B.T. and B.K.; data curation, A.W.-S., B.T. and B.K.; writing, A.W.-S., B.T. and A.B; draft preparation, A.W.-S., B.T. and A.B.; writing—review and editing, B.T. and A.B.; visualization, A.W.-S.; supervision, B.T. and A.B.; project administration, B.T. and B.K. All authors have read and agreed to the published version of the manuscript.

## Institutional Review Board Statement

The study was conducted in accordance with the Declaration of Helsinki and approved by the Ethics Committee of the District Medical Chamber in Szczecin (Poland) (Nr OIL-Sz/MF/KB/452/05/07/2018; Nr OIL-SZ/MF/KB/450/UKP/10/2018). The study was not preregistered before commencement.

## Informed Consent Statement

Informed consent was obtained from all subjects involved in the study.

## Data Availability Statement

Data are contained within the article.

## Appendix 1

As part of the conducted research, a Polish adaptation and validation were carried out for the International Spinal Cord Injury Male Sexual Function Basic Data Set (Version 2.0) and the International Spinal Cord Injury Female Sexual Function Basic Data Set developed by the International Spinal Cord Society (ISCoS) [1–2].

In the first step, the tools were translated by two independent translators. Based on these translations, uniform versions of the two questionnaires were created and subjected to analysis to ensure consistency and content accuracy.

In the Polish translation, the items take the form of questions, unlike in the English version, where the items are sentence fragments. This results from linguistic specificities-formulating the items as questions is more comprehensible and natural for Polish-speaking respondents.

In the next step, the study was conducted using the translated questionnaires. A test-retest procedure was carried out with an interval of 7 to 10 days.

The study was conducted on a group of 24 women and 42 men with spinal cord injury (up to two years post-injury). During the research, demographic data and information regarding the type of injury were also collected.

The obtained results were subjected to statistical analysis. To test the reliability and validity of the tool, analyses were performed using IBM SPSS Statistics, version 29. A frequency analysis was conducted for individual responses within the tool. To examine absolute stability, the two measurements were compared using the Wilcoxon signed-rank test. To assess validity, dependency and difference analyses were performed in relation to other variables associated with spinal cord injury (including the extent and severity of the injury according to the AIS classification, and the level of injury), as well as sociodemographic variables (gender in relation to orgasm-related dysfunctions, and age). For this purpose, the Mann-Whitney *U* test and Spearman’s rho correlation analysis were applied. The level of statistical significance was set at α = 0.05. Due to the small sample size, p-values ranging between 0.05 and 0.1 were considered significant at the level of statistical tendency.

### Characteristics of the study group

Initially, the study included 24 women and 42 men. However, due to a lack of openness in discussing issues related to sexuality-which resulted in missing data in the dataset-some participants were excluded from the analysis. The characteristics of the initial sample are presented in Table 1, while the characteristics of the final sample, after excluding cases with missing data for the entire tool, are presented in Table 2.

**Tabele 1.** Characteristics of the Study Group Before Exclusion of Participants.

| Variables | Women (*n* = 24) | Men (*n* = 42) |
| --- | --- | --- |
| **Age** |  |  |
| *M* (*SD*) | 34,96 (9,94) | 34,14 (9,57) |
| *Me* (*Min.-Maks.*) | 33,00 (18-60) | 32,00 (20-55) |
| **ASIA** |  |  |
| ASIA A | 12 (50%) | 20 (47,6%) |
| ASIA B/C/D | 12 (50%) | 22 (52,4%) |
| **Spinal segment injury** |  |  |
| Spasticity | 11 (45,8%) | 26 (61,9%) |
| Flaccidity | 13 (54,2%) | 16 (38,1%) |
| **Interest in discussing sexuality** |  |  |
| Yes | 11 (45,8%) | 29 (69,0%) |
| Yes, but only to receive information | 7 (29,2%) | 7 (16,7%) |
| No | 6 (25,0%) | 6 (14,3%) |

**Tabele 2.**
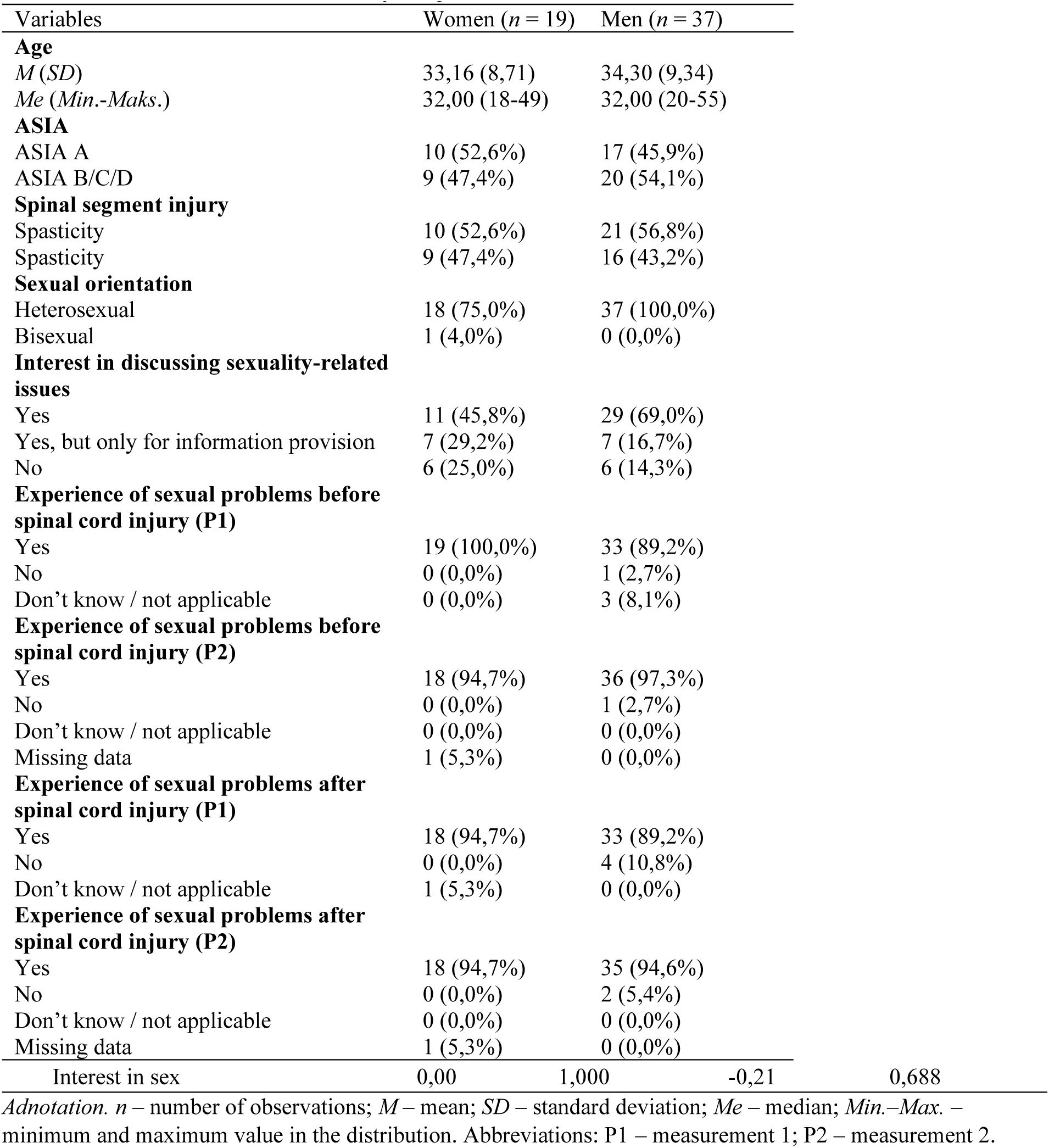
Characteristics of the final study sample.

### Test-retest reliability of the tool

To verify the reliability of the tool in terms of test-retest stability, a comparison of sexual functioning symptoms reported by participants was conducted. Due to the distinct nature of symptoms (apart from orgasm), analyses were carried out separately for the female and male samples. Tables 3 and 4 and Figures 1–4 present the distribution of responses. Responses such as “I don’t know/not applicable” were excluded from the analysis, resulting in an ordinal variable. To compare both measurements, the Wilcoxon signed-rank test was used. The analyses were first performed for the group of women (Table 3), followed by the group of men (Table 4).

**Table 3.** Analysis of the distribution of sexual functioning in the female group for measurements 1 and 2, including results of the Wilcoxon signed-rank test.

| Including results of the Wilcoxon signed-rank test. |  |  |  |  |  |  |  |
| --- | --- | --- | --- | --- | --- | --- | --- |
| Variables | P1 | | P2 | | Z | p | $\eta^2$ |
|  | N (%) | Me (IQR) | N (%) | Me (IQR) |  |  |  |
| **Psychogenic genital arousal** |  |  |  |  |  |  |  |
| Normal | 8 (42,1%) |  | 7 (36,8%) |  |  |  |  |
| Altered/reduced | 7 (36,8%) |  | 6 (31,6%) |  |  |  |  |
| Absent | 4 (21,1%) | 2,00 (1,00) | 5 (26,3%) | 2,00 (2,00) | -1,00 | 0,317 | 0,03 |
| Don't know/not applicable | 0 (0,0%) |  | 0 (0,0%) |  |  |  |  |
| Missing data | 0 (0,0%) |  | 1 (5,3%) |  |  |  |  |
| **Reflex genital arousal** |  |  |  |  |  |  |  |
| Normal | 2 (10,5%) |  | 2 (10,5%) |  |  |  |  |
| Altered/reduced | 12 (63,2%) |  | 12 (63,2%) |  |  |  |  |
| Absent | 5 (26,3%) | 2,00 (1,00) | 5 (26,3%) | 2,00 (1,00) | <0,01 | 1,000 | <0,01 |
| Don't know/not applicable | 0 (0,0%) |  | 0 (0,0%) |  |  |  |  |
| Missing data | 0 (0,0%) |  | 0 (0,0%) |  |  |  |  |
| **Menstruation** |  |  |  |  |  |  |  |
| Normal | 18 (94,7%) |  | 18 (94,7%) |  |  |  |  |
| Altered/reduced | 0 (0,0%) |  | 0 (0,0%) |  |  |  |  |
| Absent | 1 (5,3%) | 1,00 (0,00) | 1 (5,3%) | 1,00 (0,00) | <0,01 | 1,000 | <0,01 |
| Don't know/not applicable | 0 (0,0%) |  | 0 (0,0%) |  |  |  |  |
| Missing data | 0 (0,0%) |  | 0 (0,0%) |  |  |  |  |
| **Orgasm** |  |  |  |  |  |  |  |
| Normal | 2 (10,5%) |  | 1 (5,3%) |  |  |  |  |
| Altered/reduced | 5 (26,3%) |  | 5 (26,3%) |  |  |  |  |
| Absent | 8 (42,1%) | 3,00 (1,00) | 10 (52,6%) | 3,00 (1,00) | <0,01 | 1,000 | <0,01 |
| Don't know/not applicable | 4 (21,1%) |  | 2 (10,5%) |  |  |  |  |
| Missing data | 0 (0,0%) |  | 1 (5,3%) |  |  |  |  |

*Annotation.* N – number of observations; Me – median; IQR – interquartile range; Z – test statistic value; p – statistical significance; $\eta^2$ – effect size indicator.

**Table 4.** Analysis of the distribution of sexual functioning in the male group for measurements 1 and 2, along with Wilcoxon signed-rank test results.

| Mann-Whitney U signed rank test results: |  |  |  |  |  |  |  |
| --- | --- | --- | --- | --- | --- | --- | --- |
| Variable | P1 |  | P2 |  | Z | p | η<sup>2</sup> |
|  | N (%) | Me (IQR) | N (%) | Me (IQR) |  |  |  |
| **Psychogenic erection** |  |  |  |  |  |  |  |
| Normal | 6 (16,2%) |  | 8 (21,6%) |  |  |  |  |
| Altered/reduced | 22 (59,5%) |  | 20 (54,1%) |  |  |  |  |
| Absent | 8 (21,6%) | 2,00 (0,00) | 9 (24,3%) | 2,00 (0,50) | -1,41 | 0,157 | 0,03 |
| Don't know/not applicable | 1 (2,7%) |  | 0 (0,0%) |  |  |  |  |
| Missing data | 0 (0,0%) |  | 0 (0,0%) |  |  |  |  |
| **Reflex erection** |  |  |  |  |  |  |  |
| Normal | 6 (16,2%) |  | 8 (21,6%) |  |  |  |  |
| Altered/reduced | 22 (59,5%) |  | 20 (54,1%) |  |  |  |  |
| Absent | 9 (24,3%) | 2,00 (0,50) | 9 (24,3%) | 2,00 (0,50) | -0,82 | 0,414 | 0,01 |
| Don't know/not applicable | 0 (0,0%) |  | 0 (0,0%) |  |  |  |  |
| Missing data | 0 (0,0%) |  | 0 (0,0%) |  |  |  |  |
| **Ejaculation** |  |  |  |  |  |  |  |
| Normal | 3 (8,1%) |  | 3 (8,1%) |  |  |  |  |
| Altered/reduced | 11 (29,7%) |  | 10 (27,0%) |  |  |  |  |
| Absent | 22 (59,5%) | 3,00 (1,00) | 23 (62,2%) | 3,00 (1,00) | -1,00 | 0,317 | 0,01 |
| Don't know/not applicable | 1 (2,7%) |  | 0 (0,0%) |  |  |  |  |
| Missing data | 0 (0,0%) |  | 0 (0,0%) |  |  |  |  |
| **Orgasm** |  |  |  |  |  |  |  |
| Normal | 3 (8,1%) |  | 1 (2,7%) |  |  |  |  |
| Altered/reduced | 9 (24,3%) |  | 12 (32,4%) |  |  |  |  |
| Absent | 21 (56,8%) | 3,00 (1,00) | 21 (56,8%) | 3,00 (1,00) | -1,00 | 0,317 | 0,02 |
| Don't know/not applicable | 4 (10,8%) |  | 3 (8,1%) |  |  |  |  |
| Missing data | 0 (0,0%) |  | 0 (0,0%) |  |  |  |  |

**Figure 1.**
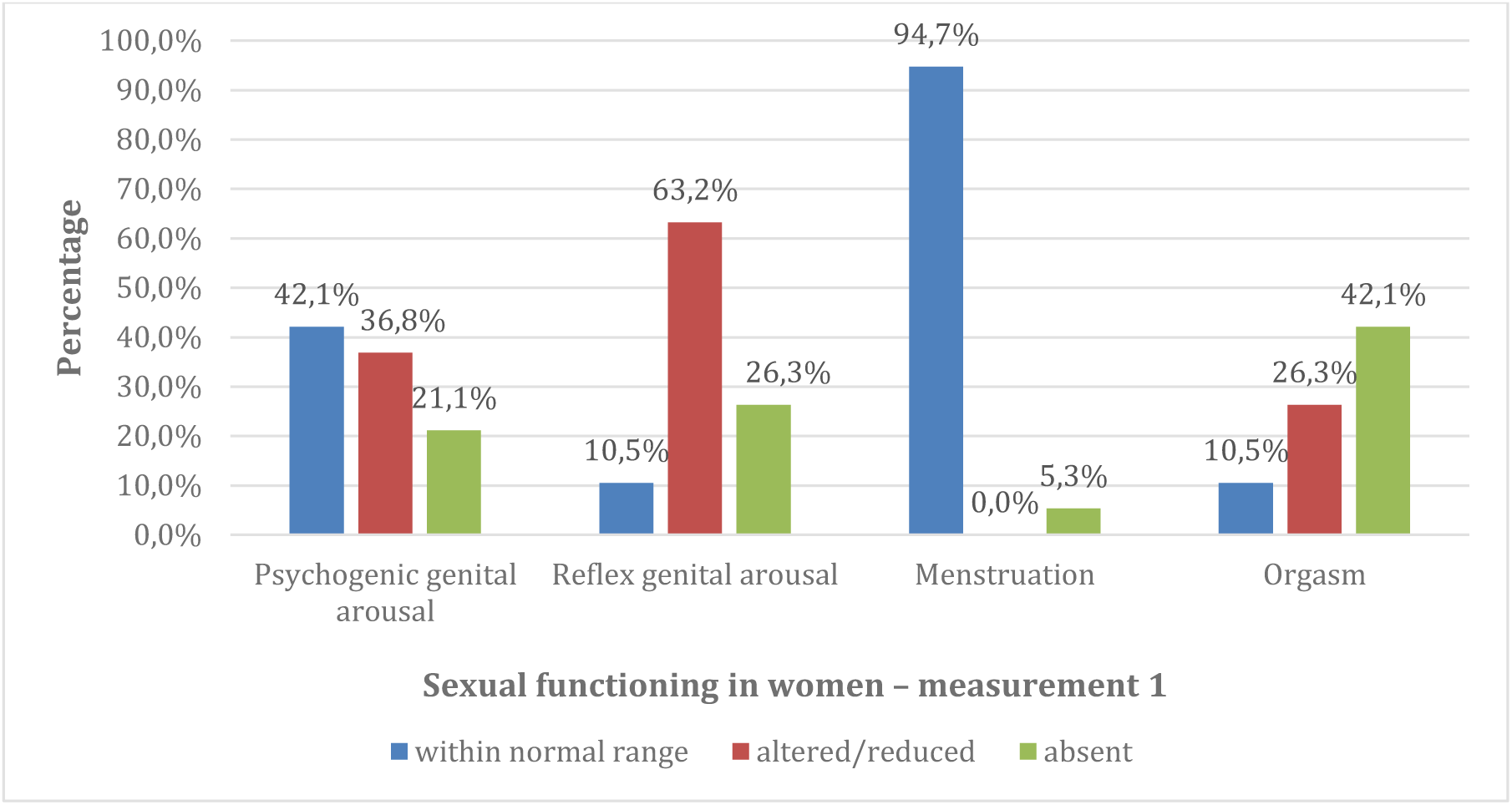
Percentage distribution of responses regarding sexual functioning disorders among women in the first assessment.

**Figure 2.**
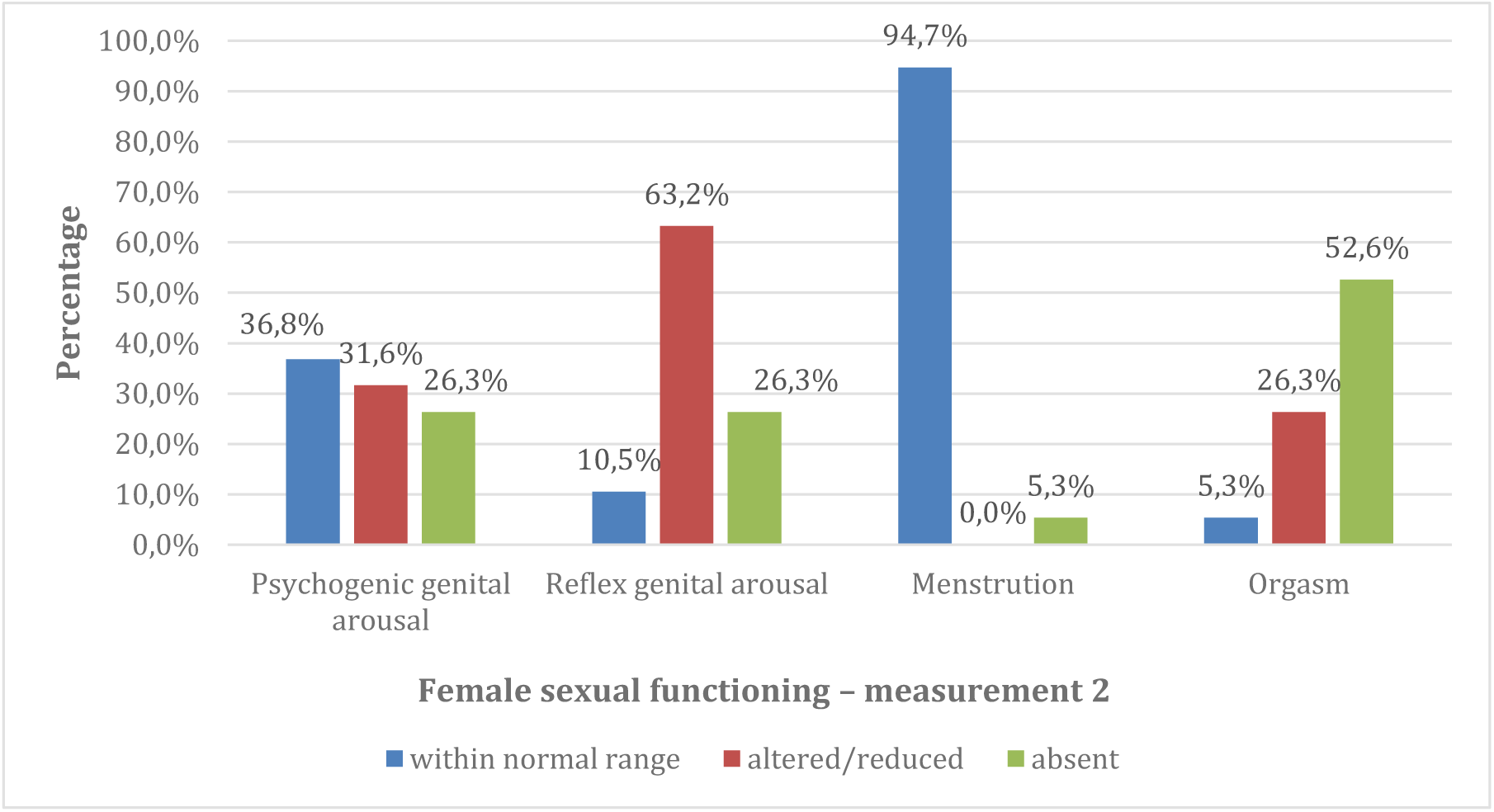
Percentage distribution of responses regarding sexual functioning disorders among women in the second assessment.

**Figure 3.**
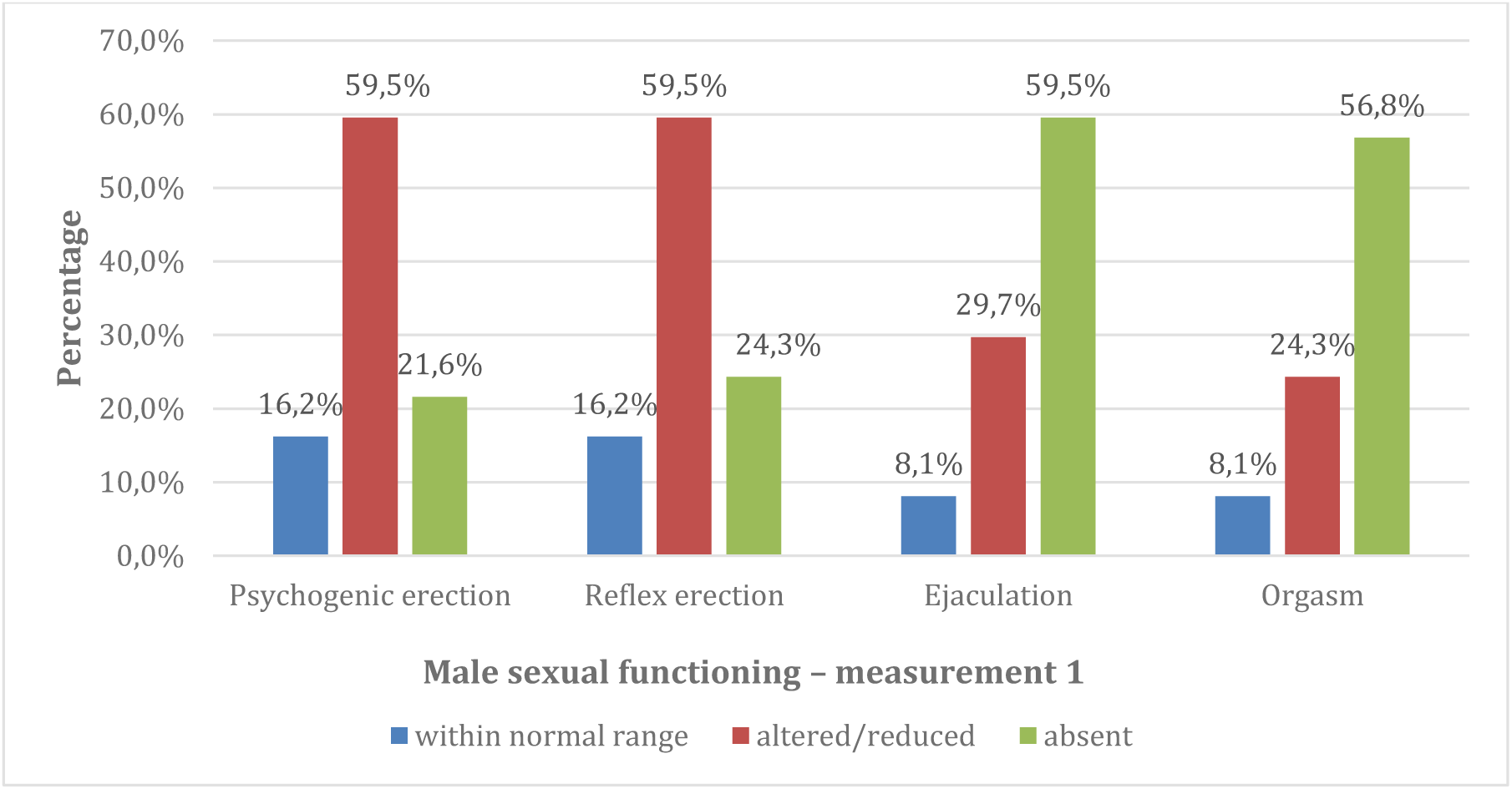
Percentage distribution of responses to questions about sexual dysfunctions among men in the first measurement

**Figure 4.**
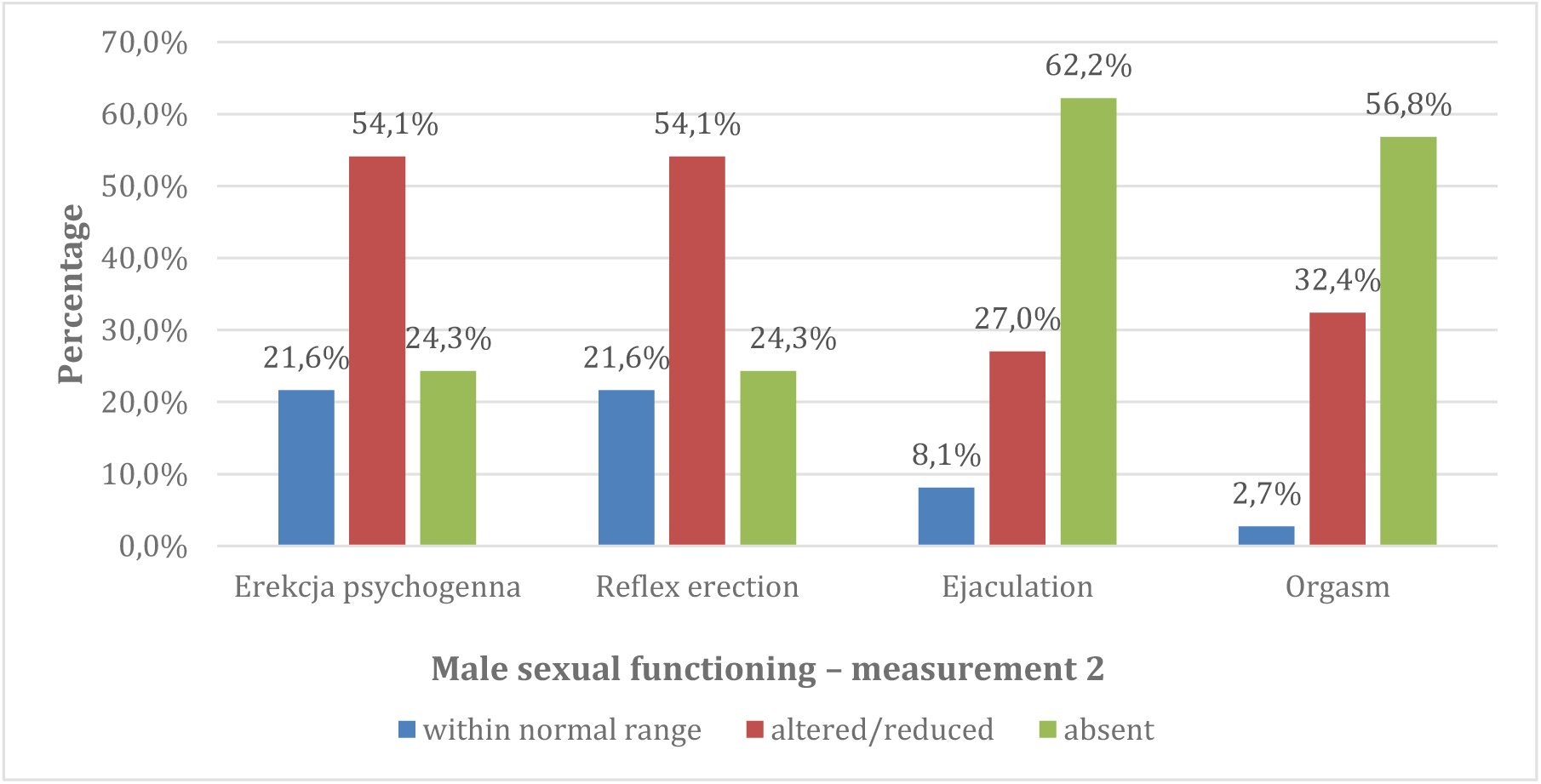
Percentage distribution of responses to questions about sexual dysfunctions among men in the second measurement

No significant differences were found between the measurements, indicating their temporal stability. The severity of symptoms did not differ significantly between the two measurements.

### Validity of the tool

In the next stage of statistical analysis, the relationships and differences between additional variables related to spinal cord injury and sociodemographic variables were examined to assess the convergent and discriminant validity of the tool. In the first step, groups divided according to the ASIA classification – A vs. B/C/D – were compared in terms of the severity of sexual functioning disorders using the Mann–Whitney U test (Table 5).

**Table 5.** Mann–Whitney U test results for symptoms related to sexual functioning in female and male groups according to spinal cord injury classification.

| Dependent variable | AIS A<br>($n_k = 10$)<br>($n_m = 17$) | | | AIS B/C/D<br>($n_k = 9$)<br>($n_m = 20$) | | | Z | p | $\eta^2$ |
| --- | --- | --- | --- | --- | --- | --- | --- | --- | --- |
|  | mean rank | Me | IQR | mean rank | Me | IQR |  |  |  |
| **Women** |  |  |  |  |  |  |  |  |  |
| Psychogenic genital arousal (P1) | 10,85 | 2,00 | 1,25 | 9,06 | 1,00 | 1,50 | -0,74 | 0,471 | 0,03 |
| Psychogenic genital arousal (P2) | 9,65 | 2,00 | 1,25 | 9,31 | 1,50 | 2,00 | -0,14 | 0,826 | <0,01 |
| Reflex genital arousal (P1) | 10,20 | 2,00 | 0,25 | 9,78 | 2,00 | 1,50 | -0,19 | 1,000 | <0,01 |
| Reflex genital arousal (P2) | 10,20 | 2,00 | 0,25 | 9,78 | 2,00 | 1,50 | -0,19 | 1,000 | <0,01 |
| Menstruation (P1) | 10,45 | 1,00 | 0,00 | 9,50 | 1,00 | 0,00 | -0,95 | 1,000 | 0,05 |
| Menstruation (P2) | 10,45 | 1,00 | 0,00 | 9,50 | 1,00 | 0,00 | -0,95 | 1,000 | 0,05 |
| Orgasm (P1) | 8,61 | 3,00 | 1,00 | 7,08 | 2,50 | 2,00 | -0,72 | 0,483 | 0,04 |
| Orgasm (P2) | 8,50 | 3,00 | 1,00 | 8,50 | 3,00 | 1,25 | 0,00 | 1,000 | <0,01 |
| **Men** |  |  |  |  |  |  |  |  |  |
| Psychogenic erection (P1) | 18,56 | 2,00 | 0,00 | 18,45 | 2,00 | 0,75 | -0,04 | 0,971 | <0,01 |
| Psychogenic erection (P2) | 18,62 | 2,00 | 1,00 | 19,33 | 2,00 | 0,75 | -0,22 | 0,827 | <0,01 |
| Reflex erection (P1) | 16,03 | 2,00 | 0,50 | 21,53 | 2,00 | 1,00 | -1,75 | 0,080 | 0,09 |
| Reflex erection (P2) | 16,91 | 2,00 | 0,50 | 20,78 | 2,00 | 1,00 | -1,20 | 0,232 | 0,04 |
| Ejaculation (P1) | 20,94 | 3,00 | 0,75 | 16,55 | 2,50 | 1,00 | -1,44 | 0,150 | 0,06 |
| Ejaculation (P2) | 21,50 | 3,00 | 0,00 | 16,10 | 2,50 | 1,00 | -1,80 | 0,071 | 0,09 |
| Orgasm (P1) | 18,60 | 3,00 | 1,00 | 15,67 | 3,00 | 1,00 | -1,02 | 0,307 | 0,03 |
| Orgasm (P2) | 19,60 | 3,00 | 1,00 | 15,84 | 3,00 | 1,00 | -1,29 | 0,198 | 0,05 |

*Annotation.* n – number of observations; Me – median; IQR – interquartile range; Z – test statistic value; p – statistical significance; $\eta^2$ – effect size. In the group of women, exact two-tailed significance was reported. In both groups, there were missing data for some comparisons: the lowest sample size for the AIS A group among women was 9, and for ASIA B/C/D – 6; the lowest sample size for the AIS A group among men was 15, and for AIS B/C/D – 18.

The analysis revealed no statistically significant differences within the group of women. Women in the AIS A group did not differ from those in the AIS B/C/D group in terms of sexual functioning. Likewise, no statistically significant differences at the *p* < 0.05 level were observed in the male group. However, results significant at the level of statistical tendency (0.05 < *p* < 0.1) were noted for reflexive erection disorders in the first measurement and ejaculation in the second measurement (both effects were moderate). Greater severity of reflexive erection-related symptoms was observed in men from the AIS B/C/D group compared to AIS A. Conversely, the opposite relationship was found for ejaculation – a higher level of ejaculation-related dysfunction was recorded in men from the AIS A group than in AIS B/C/D. Other differences were neither statistically significant nor significant at the level of statistical tendency.

Next, it was examined whether the participants differed in terms of symptoms related to sexual functioning depending on whether their condition indicated spasticity or flaccidity. Once again, the Mann–Whitney U test was conducted (Table 6).

**Table 6.** Results of the Mann–Whitney U test for sexual functioning symptoms in female and male participants depending on the type of neurological impairment.

| Dependent variable | Spasticity<br>($n_k = 10$)<br>($n_m = 21$) | | | Flaccidity<br>($n_k = 9$)<br>($n_m = 16$) | | | $Z$ | $p$ | $\eta^2$ | |
| --- | --- | --- | --- | --- | --- | --- | --- | --- | --- | --- |
| | mean rank | $Me$ | $IQR$ | mean rank | $Me$ | $IQR$ | | | | |
| **Women** |  |  |  |  |  |  |  |  |  |  |
| Psychogenic genital arousal (P1) | 9,55 | 2,00 | 1,00 | 10,50 | 2,00 | 2,00 | -0,39 | 0,772 | <0,01 |  |
| Psychogenic genital arousal (P2) | 9,00 | 2,00 | 1,25 | 10,13 | 2,00 | 2,00 | -0,47 | 0,774 | 0,01 |  |
| Reflex genital arousal (P1) | 8,50 | 2,00 | 0,00 | 11,67 | 3,00 | 1,50 | -1,43 | 0,123 | 0,11 |  |
| Reflex genital arousal (P2) | 8,50 | 2,00 | 0,00 | 11,67 | 3,00 | 1,50 | -1,43 | 0,123 | 0,11 |  |
| Menstruation (P1) | 10,45 | 1,00 | 0,00 | 9,50 | 1,00 | 0,00 | -0,95 | 1,000 | 0,05 |  |
| Menstruation (P2) | 10,45 | 1,00 | 0,00 | 9,50 | 1,00 | 0,00 | -0,95 | 1,000 | 0,05 |  |
| Orgasm (P1) | 7,44 | 2,00 | 1,00 | 8,64 | 3,00 | 2,00 | -0,58 | 0,685 | 0,02 |  |
| Orgasm (P2) | 7,33 | 2,00 | 1,00 | 10,00 | 3,00 | 0,00 | -1,30 | 0,250 | 0,11 |  |
| **Men** |  |  |  |  |  |  |  |  |  |  |
| Psychogenic erection (P1) | 16,98 | 2,00 | 0,00 | 20,63 | 2,00 | 1,00 | -1,18 | 0,238 | 0,04 |  |
| Psychogenic erection (P2) | 16,57 | 2,00 | 1,00 | 22,19 | 2,00 | 1,00 | -1,73 | 0,084 | 0,08 |  |
| Reflex erection (P1) | 17,79 | 2,00 | 0,00 | 20,59 | 2,00 | 1,00 | -0,89 | 0,374 | 0,02 |  |
| Reflex erection (P2) | 17,93 | 2,00 | 0,50 | 20,41 | 2,00 | 1,00 | -0,76 | 0,446 | 0,02 |  |
| Ejaculation (P1) | 16,55 | 2,50 | 1,00 | 20,94 | 3,00 | 0,75 | -1,44 | 0,150 | 0,06 |  |
| Ejaculation (P2) | 16,93 | 3,00 | 1,00 | 20,47 | 3,00 | 0,75 | -1,18 | 0,237 | 0,04 |  |
| Orgasm (P1) | 15,24 | 3,00 | 1,00 | 18,88 | 3,00 | 0,75 | -1,27 | 0,203 | 0,05 |  |
| Orgasm (P2) | 17,05 | 3,00 | 1,00 | 18,07 | 3,00 | 1,00 | -0,35 | 0,728 | <0,01 |  |

*Annotation.* In the female group, exact two-tailed significance was reported. In both groups, data were missing for some comparisons: the lowest sample size for the spasticity group among women was 8, and for the flaccidity group – 7; the lowest sample size for the spasticity group among men was 17, and for the flaccidity group – 15.

The results did not reveal any significant differences between the compared groups regarding sexual functioning among women. In the male group, only one result reached the level of statistical trend significance for psychogenic erection measured in the second assessment. Eta squared indicated a moderate effect size, and based on the analysis of mean ranks -with caution -it can be assumed that participants with flaccidity exhibited greater severity of psychogenic erection dysfunction symptoms than those with spasticity. The p-values for the remaining comparisons exceeded 0.1.

The relationship between sociodemographic variables and sexual functioning was also examined. Initially, the association between participants’ age and their symptoms related to sexual life was tested.

Due to the ordinal measurement of symptoms, a correlation analysis using Spearman’s rho coefficient was selected (Table 7).

**Table 7.** Results of Spearman’s rho correlation analysis between age and symptoms related to sexual functioning in women and men.

| Variable | Age |  |  |
| --- | --- | --- | --- |
|  | *n* | *Spearman's rho* | *p* |
| **Women** |  |  |  |
| Psychogenic genital arousal (P1) | 19 | 0,27 | 0,259 |
| Psychogenic genital arousal (P2) | 18 | 0,16 | 0,529 |
| Reflexive genital arousal (P1) | 19 | 0,12 | 0,619 |
| Reflexive genital arousal (P2) | 19 | 0,12 | 0,619 |
| Menstruation (P1) | 19 | -0,02 | 0,930 |
| Menstruation (P2) | 19 | -0,02 | 0,930 |
| Orgasm (P1) | 15 | 0,08 | 0,776 |
| Orgasm (P2) | 16 | -0,23 | 0,395 |
| **Men** |  |  |  |
| Psychogenic erection (P1) | 36 | -0,10 | 0,575 |
| Psychogenic erection (P2) | 37 | 0,05 | 0,766 |
| Reflexive erection (P1) | 37 | 0,30 | 0,072 |
| Reflexive erection (P2) | 37 | 0,18 | 0,275 |
| Ejaculation (P1) | 36 | 0,10 | 0,576 |
| Ejaculation (P2) | 36 | 0,10 | 0,563 |
| Orgasm (P1) | 33 | 0,14 | 0,440 |
| Orgasm (P2) | 34 | 0,21 | 0,226 |

The results of the analysis did not reveal any statistically significant effects – age is not associated with sexual functioning in either women or men. A relationship approaching statistical significance was found between age and reflexive erection problems in the first measurement. The correlation coefficient indicates that the relationship should be interpreted as moderate and positive, meaning that as age increases, the severity of reflexive erection dysfunction also increases.

Additionally, gender differences in orgasm-related symptoms were examined, as orgasm was the only symptom common to both sexes. For this purpose, the Mann–Whitney U test was conducted. The results are presented in Table 8.

**Table 8.** Results of the Mann–Whitney U test for orgasm-related symptoms by gender.

| Variable | Women<br>($n_{p1} = 15$)/($n_{p2} = 16$) | | | Men<br>($n_{p1} = 33$)/($n_{p2} = 34$) | | | Z | P | $\eta^2$ |
| --- | --- | --- | --- | --- | --- | --- | --- | --- | --- |
|  | mean rank | Me | IQR | mean rank | Me | IQR |  |  |  |
| Orgasm (P1) | 22,70 | 3,00 | 1,00 | 25,32 | 3,00 | 1,00 | -0,69 | 0,489 | 0,01 |
| Orgasm (P2) | 25,41 | 3,00 | 1,00 | 25,54 | 3,00 | 1,00 | -0,04 | 0,971 | <0,01 |

The results were not statistically significant. This indicates that women did not differ from men in terms of sexual functioning related to orgasm.

### Conclusions

It was assumed that the tool demonstrates reliability, as confirmed by its temporal stability. Its validity was only partially confirmed-primarily discriminant validity. The absence of expected differences may result from the low sample size in the compared groups. Nevertheless, the properties of the tool were deemed sufficient for use in research. Further work on the validity of the Polish version of the tool is recommended.

## Appendix No. 2

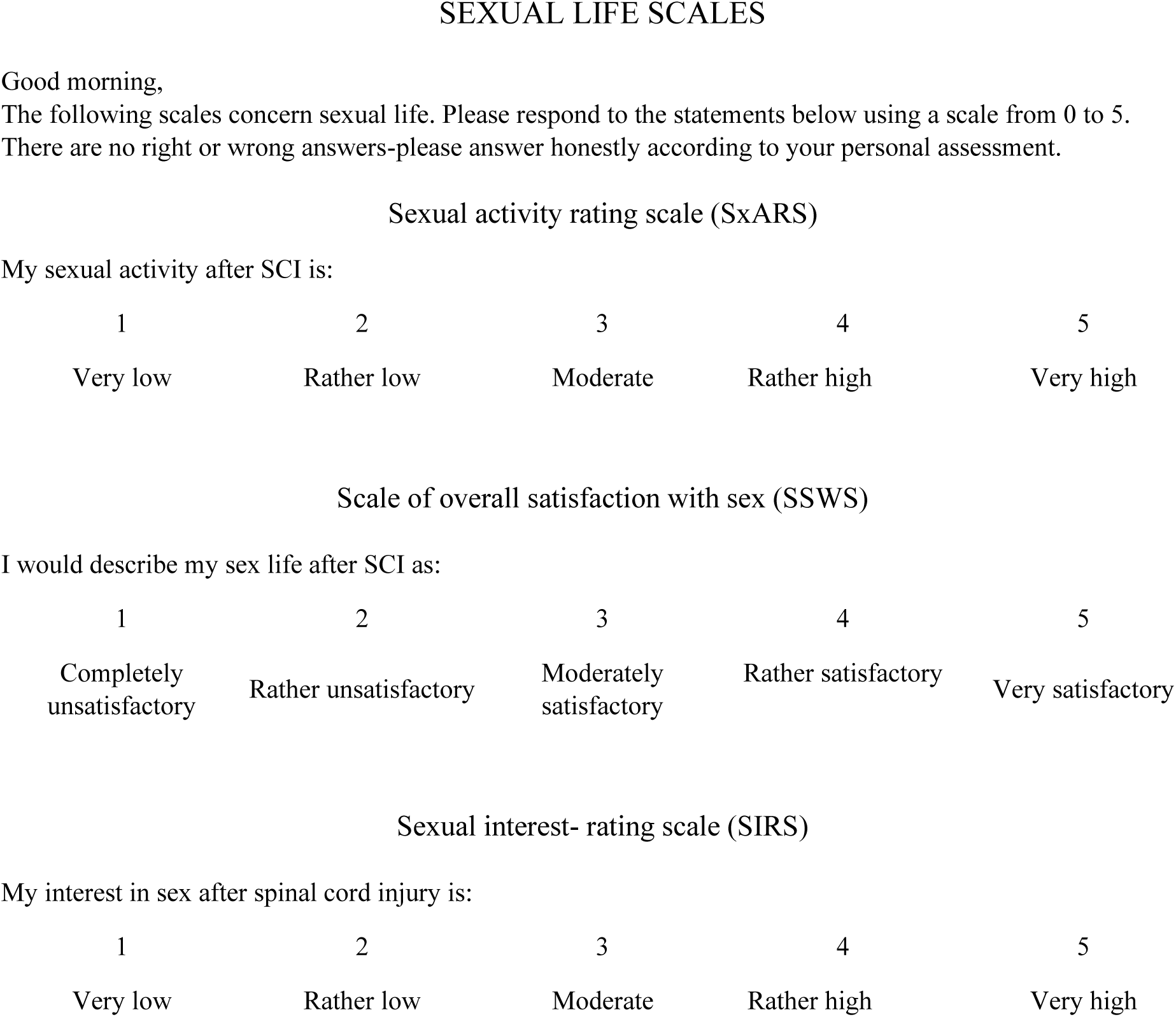

## Appendix No. 3

The intervention involved the patients themselves or, whenever possible, also their partners.

In the first step, a medical interview and examinations were conducted. Then, screening questionnaire assessments and a psychological interview were carried out, focusing on evaluating the level of openness to conversation and motivation to work on sexuality-related matters; satisfaction with intimate life; understanding and approach to sexual difficulties that emerged after SCI; main fears and concerns related to intimate life after SCI; level of body acceptance and self-esteem in the context of physical and sexual attractiveness; issues related to partnership (relationship status, partner dynamics, experience of being in an intimate relationship); perception of oneself as a sexual partner (beliefs); internalized prejudices regarding the possibilities of treatment and sexual potential after SCI and in the context of the patient’s specific problems. The time allocated for the interview was not strictly fixed, as it depended on the openness and sensitivity of the patient or the couple. Some participants needed more time and the establishment of a certain level of rapport with the psychologist-therapist.

After the completion of the questionnaire assessments and the interview, the intervention involved a meeting with a physician, a rehabilitation specialist, and a representative of the Active Rehabilitation Foundation (FAR). The meetings with the FAR representative and the attending physician focused on providing information regarding first-, second-, and third-line treatment options, as well as suggesting pharmacological treatment or equipment most suitable for the individual patient.

First-line treatment for increasing libido in women includes the use of flibanserin (unfortunately expensive and difficult to obtain in Poland). As supportive therapy for insufficient vaginal lubrication, patients were advised to use commonly available lubricants or hyaluronic acid-based vaginal gels. In cases of low serum testosterone levels potentially contributing to difficulties with libido and sexual arousal, oral testosterone supplementation was recommended, along with the implementation of a healthy lifestyle-achieving a normal BMI, following a high-protein, low-carbohydrate diet, and engaging in aerobic exercise at appropriate intensity and frequency [1].

In cases of sexual arousal disorders in men, the first-line treatment involved the use of phosphodiesterase type 5 (PDE5) inhibitors, such as sildenafil (Viagra), vardenafil (Levitra), or tadalafil (Cialis).

In cases where the effects of these medications were unsatisfactory, second-line interventions were used, such as intracavernosal injections of prostaglandin E1, papaverine, or phentolamine, as well as devices including vacuum erection devices (used alone or with a rubber or silicone ring placed at the base of the penis). In cases of additional difficulties with ejaculation, penile vibratory stimulators (PVS) were recommended, combined with the use of midodrine or electroejaculation via a rectal probe (EEJ) [2–3]. If these treatments proved ineffective, surgical options involving penile prostheses were available, although none of our patients were offered such solutions. Likewise, the use of devices such as vacuum pumps and constriction rings was not a common or preferred method of treatment [3–5].

The physiotherapist recommended exercises aimed at improving sexual function, which were incorporated into the patient’s six-week rehabilitation program.

The psychological intervention consisted of sixty-minute individual sessions (or sessions with partners) and two-day group workshops. (Due to low post-pandemic attendance, the workshops were suspended in July 2021.) After conducting an in-depth interview, targeted psychoeducation was provided concerning sexual difficulties and available treatment methods. This included learning about one’s own body and its reactions during autoerotic activity, and where possible, together with a sexual partner. The educational content covered the following topics:

– exploring erogenous zones located beyond the genitals, such as nipples, neck, armpits, lips, ears, etc.;
– discovering the most pleasurable types of stimulation, deepening and associating these sensations with observing a partner during foreplay, arousal triggered by erotic fantasies, and memories of previously experienced emotions;
– caresses and associating them with pleasant mental sensations and stimulation of other senses, including genital stimulation-even in cases of people lacking sensation in those areas;
– becoming familiar with one’s own body and initially assessing difficulties with arousal and the potential for orgasm, as well as monitoring progress over time;
– supplementing the information provided by other professionals, addressing misconceptions, beliefs, and fears regarding pharmacological treatments using medical devices, aids, and lubricants;
– strategies for preparing for intimacy, such as timing the intake of erection- or arousal-enhancing medications, emptying the bowels beforehand, positioning the catheter in a comfortable and discreet way, and ensuring proper hygiene;
– factors that improve the quality of intimate encounters, such as expanding and diversifying foreplay and the atmosphere to make it more engaging for the specific couple; seeking comfortable and suitable sexual positions; learning about the body and mapping erogenous zones; prolonging or intensifying stimulation of these areas; trying out different forms of caressing and sexual acts; and engaging additional senses (smell, taste, sight, and hearing).

The therapeutic intervention focused primarily on cognitive and behavioral elements and included the following:

– work in the area of emotions, aimed at developing the ability to name and gain insight into one’s emotional states and their associated physiological responses. This was particularly helpful for patients with low acceptance of their bodies after SCI. Cognitive-behavioral techniques and mindfulness-based approaches were used in this area.
– addressing cognitive distortions, mainly related to the perception of one’s own sexuality after SCI, attractiveness, and body image (appearance, limitations). Body image and self-acceptance, as well as dysfunctional beliefs and coping strategies, are common predictors of low self-esteem, mood disturbances, and even depression. This component employed cognitive-behavioral techniques, mindfulness practices, and coping skills training based on DBT, emphasizing a non-judgmental attitude and the practice of gratitude.
– analysis of personal values related to partnership and sexuality, along with the modification and flexibilization of dysfunctional beliefs about oneself as a sexual and life partner. CBT techniques were primarily used here.
– communication and assertiveness training, aimed at developing the ability to communicate one’s own needs clearly and openly, and to discuss mutual needs with one’s partner.

The two forms of intervention-psychoeducation and the implementation of therapeutic techniques-were interwoven depending on the patient’s readiness to discuss specific issues and engage in the intervention process. For example, in the case of a patient struggling with body acceptance after SCI, therapeutic intervention was prioritized over psychoeducation concerning intimate encounters.

Most patients participated in all of the above-mentioned therapeutic interventions; however, psychoeducation related to partner relationships was provided to a limited extent due to the patients attending the sessions without their partners.

The majority of project participants (N=52, including 26 women and 26 men) were satisfied with the sexuality-related interventions introduced in the project. A total of 88.46% of respondents reported being either *very satisfied or satisfied* with their participation in the workshops, which followed a common framework while also allowing for individualization and adjustment of the discussed content to meet the specific needs of each participant.

The remaining 11.54% of participants *had no opinion*. No negative evaluations were recorded, and none of the respondents expressed dissatisfaction with the intervention. Qualitative data were also collected in the form of patient feedback, which enabled improvements to the implemented interventions. Participants most frequently pointed out the lack of detailed explanations, discussions, and demonstrations concerning devices used to improve erection and ejaculation. Efforts were made to enhance this component of the support provided in the area of sexual education for patients after spinal cord injury (SCI).

